# The somatic and germline mutational landscape of HPV-negative oral cancer patients with a history of chewing tobacco and betel nut use

**DOI:** 10.64898/2026.01.20.26344267

**Authors:** Hafeeda Kunhabdulla, Manju Kashyap, Mohammed S Mustak, Rohan Thomas Mathew, Arun Khattri, H.T. Marc Timmers, Sheikh Nizamuddin, Riaz Abdulla

## Abstract

Head and neck cancer (HNC) is highly prevalent in South-Asia, driven by additional region-specific exposures such as chewing tobacco and betel nut. Despite therapeutic advances, five-year survival rate remains around 50-60%, underscoring urgent need to identify novel therapeutic targets and improve disease-free survival. This study was designed to identify both somatic and germline drivers contributing to HNC pathogenesis. Through whole-exome sequencing of 103 patients, we detected mutations in known HNC drivers (TP53, CDKN2A, NOTCH1) as well as novel hotspots in several genes, including TRIM48, MAP3K19, and CDC20. A recurrent hotspot mutation (p.A187T) in POLQ gene was identified in patients with high tumor mutation burden and was absent in both TCGA and ICGC cohorts. Among known hotspots, the MYC p.T73A mutation was highly prevalent, occurring in over 50% of patients. As MYC is considered an “undruggable” target, alternative strategies targeting upstream regulators such as BRD4 with specific inhibitors may hold promise for South-Asian HNSCC patients harboring the p.T73A mutation. Copy-number variation analysis further revealed EGFR amplification and TP73 deletion in the majority of patients, highlighting additional layers of genomic dysregulation. Comparative genomic analyses showed no recurrent mutations in epigenetic regulators (ARID2, EP300, KMT2B/MLL2, KMT2D/MLL4, NSD1, and TET1). We report p.S456L germline variant in SDHA consistently among South-Asian cohorts. Patients with p.S456L mutation were younger than those without it, reflecting typical epidemiological signature of a genetic variant that increases susceptibility. Systematic molecular characterization of recurrent mutations is required to elucidate mechanism of action of these variants and to find actionable therapeutic targets.

## Introduction

Head and neck cancer is the seventh most common cancer globally, with approximately 0.9 million new cases reported each year [1]. Most of these cases (41%) have tumors located in the lip and oral cavity. Other frequently affected sites include the larynx (20%), nasopharynx (13%), oropharynx (11%), hypopharynx (9%), and salivary glands (6%). The high incidence of lip and oral cavity cancer is mirrored in the South Asian populations with an age-standardized rate (ASR) of 14.1 per 100,000. To systematically document this, National initiatives such as the *National Oncology Cancer Registry (NOCR)* have been instrumental in documenting incidence and mortality patterns across the India, while specialized genomic databases like *GENVOC* provide valuable molecular insights into Indian oral cancer cohorts [2]. Together, these resources highlight the disproportionately high burden of head and neck cancers in India.

Compared to patients from other regions—where the major risk factors include smoking, alcohol consumption, and HPV infection (especially HPV-16)—South Asian populations also commonly use chewing tobacco and betel nuts, which serve as additional risk factors alongside smoking and alcohol use [3, 4]. HPV-related oropharyngeal cancers are relatively rare in South Asia, whereas their incidence is rising in North America and Europe [5]. Importantly, despite advances in treatment, the five-year survival rate remains around 50–60%. This underscores not only the need for early detection and prevention strategies but also the urgent requirement to identify novel therapeutic targets and biomarkers to improve disease-free survival among patients [6].

Large cohort based studies, such as The Cancer Genome Atlas (TCGA), have been instrumental in advancing the understanding of the biology of various cancers, including head and neck cancer, and in guiding the development of therapeutic strategies [7]. Further, studies on gingivo-buccal OSCC-GB on South Asian population identified recurrently mutated genes specific to this subtype, including *USP9X*, *MLL4* (*KMT2B*), *ARID2*, *UNC13C*, *TRPM3*, along with canonical drivers such as *TP53*, *FAT1*, *CASP8*, and *NOTCH1* and also reported frequent amplifications of *DROSHA* and *YAP1*, and homozygous deletions of *DDX3X* [8]. Recent studies from our laboratory on serum autoantibody profiling in OSCC has identified *NUBP2* as a potential diagnostic marker [9], while integrative multi-omics approaches have uncovered novel biomarkers and therapeutic targets for HNSCC [10]. These findings reveal subtype-specific mutational signatures and novel candidate drivers in South Asian oral cancers[8]. Despite this improvement, head and neck cancer patients from South Asia remain underrepresented in previous research, suggesting that novel driver genes may yet be identified in this population. Given the distinct risk factors in this region—such as areca nut and tobacco chewing—as well as differences in tumor localization, it is possible that unique driver genes contribute to cancer development. Mutations also tend to cluster in specific anatomical regions of the head and neck. For instance, recurrent mutations in the tumor suppressor genes *CTNNA2* and *CTNNA3* have been identified, with a notable prevalence in laryngeal carcinomas [11]. Since the oral cavity comprises subsites with distinct biological features the genetic drivers of cancer may also vary across different anatomical regions.

Many studies have identified pathogenic germline variants in adult cancers [12]. Pan-cancer analyses, such as those from The Cancer Genome Atlas (TCGA), the Collaborative Oncological Gene-environment Study (COGS), and Pediatric Cancer Genome Project (PCGP) have reported germline variants in tumor suppressor genes [12–14]. However, genetic risk factors identified in one population may not generalize to others due to founder effects and restricted gene flow [15]. This may be particularly relevant for South Asian sub-continent, where some risk factors observed in Caucasian populations are absent, while certain genetic factors appear to be unique to this region [15–17]. Given these considerations, we characterized both somatic and germline variants in head and neck cancer patients enrolled from a tertiary care center and hospitals in South India.

## Material and methods

### Sample details

The study included multiple cohorts of patients with head and neck squamous cell carcinoma (HNSCC). Cohort A comprised tumor samples collected in the southern region of India (2021–2023). These included formalin-fixed paraffin-embedded (FFPE) tissues from the Departments of Oral Pathology and Oncopathology, as well as fresh biopsy specimens from the Department of Surgical Oncology, Yenepoya (deemed to be University), Mangalore. The study was approved by the Yenepoya Ethics Committee (YEC-1/2023/131), and written informed consent was obtained from all patients in accordance with the declaration of Helsinki and ICMR (Indian Council of Medical Research) guidelines.

Additional patient datasets were included to increase the statistical power for detecting recurrently mutated genes. Patient-derived raw sequencing datasets (FASTQ) from SRP325222 and ERP015832 were obtained from the Sequence Read Archive (SRA) and were designated as Cohort B and Cohort C, respectively, in the current study. For comparison of the mutational landscape across patients from different geographical regions, pre-annotated somatic variant data in MAF format were obtained from cBioPortal for the TCGA-HNSCC dataset and were designated as Cohort D. Additional patient datasets were also obtained from the ICGC consortium, which includes HNSCC patients from South Asia. An overview of all cohorts is provided in Table 1.

**Table 1.**
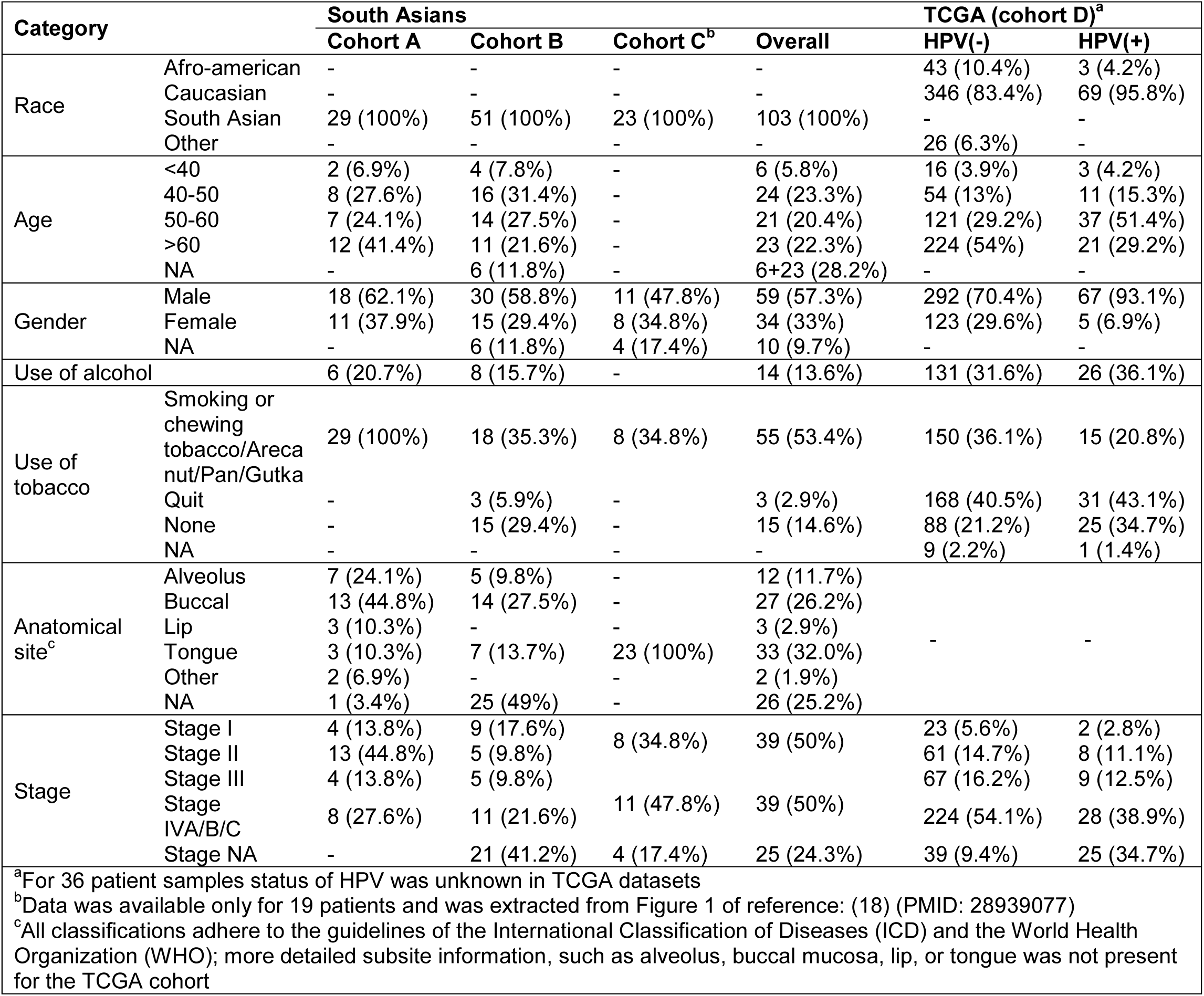
Summary of clinical and demographic characteristics of the patientsin each cohort. The table summarizes the distribution of key clinical and demographic features of the study cohort including race, age, gender, lifestyle habits (smoking, alcohol consumption), anatomical site and tumor stage.

### DNA isolation and library preparation

DNA extraction from FFPE tissues and fresh biopsied tissues were performed using the QIAamp DNA Investigator Kit (QIAGEN, Germany).The quantity of extracted DNA was confirmed using a Nanodrop 1000, with the average of two individual readings. The quality of the quantified DNA was assessed either on a 2% agarose gel or with DNA ScreenTape. For agarose gel analysis, 2 µl of DNA was mixed with 1 µl of 2x Loading Dye (Invitrogen,USA) and subjected to electrophoresis at 80 volts for 60 minutes. TekBio DNA Ladder was loaded alongside the samples. Samples with visible bands on the gel were selected for library preparation. Alternatively, when DNA ScreenTape was used, samples with a DIN value ≥ 2 were used for library preparation. Library preparation was carried out using the Twist Exome 2.0 Kit. The final libraries were quantified using a Qubit 4.0 fluorometer with the DNA HS Assay Kit, following the manufacturer’s protocol. To determine the insert size of the libraries, samples were analyzed on the TapeStation 4150 (Agilent, USA) using High Sensitivity D1000 ScreenTapes, following the manufacturer’s guidelines. The libraries were sequenced on the Illumina NovaSeq 6000 platform using a 150-nucleotide paired-end design, achieving an average coverage depth of 80–100x.

### Quality control and alignment of the reads

Paired-end reads were initially subjected to quality control filtering using fastp (v0.20.0) with default parameters to remove low-quality reads and bases. The filtered reads were then aligned to the human reference genome (hg38) using bowtie (bwa; v0.7.17-r1188). Aligned reads were further processed following GATK best practices, beginning with the removal of PCR and optical duplicates using Picard (v2.18.14), followed by base quality score recalibration with GATK (v4.0.11.0). The raw data downloaded from the ICGC portal were aligned to human reference genome (hg19).

### HPV detection

To detect HPV in the both cancer and normal tissues, HPVdetector (v1.0) was used [18]. In brief, this tool uses BWA (or Bowtie) to align sequencing reads against HPV genomes and reports any reads that successfully align. It also identifies the specific HPV type present in the raw data based on the alignment results.

### Somatic variant

Somatic variant calling was performed using the GATK-Docker pipeline (v1.0), which uses Mutect2 from GATK (v4.1.4.1). Matched normal adjacent tissues were included as controls when available. Variant calling was conducted separately for each cohort, generating a VCF file for each sample. VCF files were subsequently converted to MAF format using vcf2maf (v1.6) (https://github.com/mskcc/vcf2maf), and the resulting MAF files were merged for all samples and further analyzed using maftools (v2.22.10).The maftools was used for summarizing variants and for generating graphs. Variants were annotated using annotation database “ensembl”(hg38; v111). To identify genes with a significantly higher rate of somatic mutations, two different tools, Mutsig2cv and dNdScv (v0.0.1.0), were employed. For Mutsig2cv, datasets from all samples in Cohorts A, B, and C were merged into a single MAF file, and genomic coordinates were converted from hg38 to hg19 using CrossMap (v0.7.0), becauseMutsig2cv was designed to run on the hg19 reference genome. Genes with a q-value < 0.05 were selected for further analysis. Similarly, dNdScv was run on the same datasets (variants with hg19 genomic coordinates), and only genes with a qglobal_cv< 0.05 were retained for subsequent analysis.To identify *de novo* hotspot mutations, an R script ‘hotspot_algo’ (v0.6) was used which was applied to merged dataset of Cohort A, B and C. Genes with q value < 0.05 were considered as significant. On the other hand, to identify known hotspot mutations in somaticfreq (v0.1.0) was used. Known cancer hotspots were downloaded from ‘Cancer Hotspots’ database (v2)[19].Pathway analysis was performed using web tool “string-db” (https://string-db.org/).

### Germline variant

Germline variants were identified using the HaplotypeCaller function of GATK (version 4.5.0.0), followed by joint genotyping across samples. Variant quality score recalibration (VQSR) was subsequently performed using multiple training datasets, including variants from the 1000 Genomes Project (phase 3, version 4; prior = 15), high-confidence single nucleotide polymorphisms (SNPs) from the 1000 Genomes Project (phase 1; prior = 10), and variants from the Omni 2.5 genotyping array (prior = 12). The annotations utilized for SNP recalibration included QD, MQ, MQRankSum, ReadPosRankSum, FS, SOR, DP, and InbreedingCoeff. For indel recalibration, Mills and 1000G gold standard indels were employed with a prior of 12, using the same set of annotations except MQ. Variants that passed the recalibration filters were retained for downstream analysis using bcftools (v1.10.2). Variant annotation was performed using the Ensembl database (hg38; v111) via the Variant Effect Predictor (VEP) toolkit (v111.0). Additional variant filtering was conducted with bcftools, applying the filter ‘F_MISSING > 0.50’ to exclude variants missing in more than 50% of subjects and ‘MAF >= 0.05’ to exclude rare variants. Furthermore, the’filter_vep’ script from the VEP toolkit was used to select functional variants, specifically those categorized as missense or classified as ‘high impact’ (e.g., stop gain, stop loss), and rare in the general population (frequency < 10⁻⁴). The’isec’ function of bcftools was used to identify overlapping variants across Cohorts A, B, and C. To find the relatedness among samples, vcftools (v0.1.16) was used. To calculate the confidence intervals for patient proportions, the binom package (v1.1-1.1) in R was used, employing the “Wilson Score Interval method for binomial proportions”. Linkage disequilibrium (LD) scores were calculated using Haploview (v4.2).

### Copy number variation (CNV)

Somatic copy number variants were identified using cnv_facets (v0.16.1)[19]. Matched normal adjacent tissues were used as controls when available. To call CNVs in samples without adjacent normal tissues, we first randomly selected five adjacent tissues and merged alignment files using ‘merge’ function of samtools and then used this combined normal dataset against cancer tissues to find CNVs. For downstream analysis, GISTIC2.0 was used to identify significantly amplified or deleted regions. The output files from cnv_facets were converted into GISTIC2.0-compatible input formats using custom scripts. Briefly, Seg.CN values for GISTIC2.0 were derived from the by subtracting median log-ratio (logR) of total read depth in tumor relative to that in the normal with dipLogR values. CNV regions with a q-value less than 0.05 were considered statistically significant.

## Results

### Samples and clinical data

In total 33 cases (cohortA) from Southern part of India were selected for molecular characterization of the HNSCC cancer tissue samples (**Supplementary table 1**). Normal adjacent tissue samples were available for 24 cases. In this cohort, we also included 4 cases for which only normal adjacent tissues were available. These additional normal tissues were included to find germline variants. We also included HNSCC patients from South Asia from the two different publically available datasets (cohort B and C)[20, 21]. These two datasets are consists of 51 and 23 patient’s tissue samples, dissected from the different regions of the head and neck. For comparison, we used TCGA-HNSCC cohort (cohort D) [7, 8]. Sample details are summarized in **table 1**.

HPV-positive patient’s frequency is low in Cohort A, B and C, compared to the cohort D. None of the samples in Cohorts A and C showed evidence of HPV infection, while only four samples in Cohort B were HPV-positive (**Supplementary table 1**). Three of these were infected with HPV-16, and one sample was infected with HPV-21. In one of the HPV-16 positive samples, reads from both HPV-16 and HPV-71 were identified. The majority of patients (∼86%) in the TCGA (cohort C) were Caucasian populations. None of the patients in the cohort D were assigned to South Asian population. On average, patients in cohorts A and B developed HNSCC at a younger age (mean∼56 years) compared to Caucasian patients in the cohort D(∼61 years).The proportion of male patients was approximately twice as high as that of females in cohorts A and B. In the cohort D, the percentage of Caucasian males was about 2.5 times higher than that of female subjects. After excluding HPV-positive patients from the cohort D, the male-to-female ratio among Caucasians remained similar, with males being approximately 2.4 times more prevalent than females. These consistent patterns across cohorts suggest that males may have a higher predisposition to developing HNSCC. Moreover, all patients of the cohort C were having tongue cancer, while in cohort A and B, most of the patients were having cancer at the Buccal area. Among the patients for whom annotation was available, the prevalence was ∼45% in Cohort A and 48.3% in Cohort B, respectively (**Table 1**). Besides this, patients from the cohort D were more skewed toward advanced disease, with approximately 54% of patients diagnosed at stage IV among HPV-negative cases. Furthermore, chewing tobacco or areca nut is more common among cohort A, B and C’s patients.

### Landscape of mutation

Average depth of sequencing for cohort A, B and C is 42.3±15.0, 23.9±12.2 and 22.3±13.8 reads per bases respectively. The median number of the somatic variants per patients was 1050 and median tumor mutations burden per mega base pairs was 21 (0.88 – 655.9 mutations/MB) collectively in cohorts-A, B and C. Of these, 6 patients present in cohort B had>14,000 mutations (**Supplementary table 2**). These patients had a defect in a DNA repair gene, which could explain the higher number of mutations and is discussed in later section. As expected, the total number of mutations was significantly higher in FFPE tissues (2105±1576.5 mutations) than in fresh tissues (140.9±132.24 mutations) (**Supplementary Table 2**), which aligns with the known tendency for FFPE samples to accumulate more mutations.As expected, transition frequency (∼68%) was higher compared to transversion (∼32%). The C>T (∼51%), T>C (∼13%) and C>A (∼10%) mutations were higher in frequency (**Supplementary table 3**). Of which C>T is most prevalent in FFPE tissues while C>A is hallmark of tobacco-associated cancer tissues [22]. T>C is more commonly originate due to oxidative damage done by smoking or by defecting DNA repair pathways [23]. It is similar to cohort D where C>T and C>A were higher in frequency, except that C>G is also prevalent in tumor tissues in HPV (-) HNSCC cancer tissues in cohort D (**Supplementary table 3**).

In the adjacent normal tissue samples, we identified 0.3-12.8 million germline variants per patients, after filtering only those variants which were successfully genotyped in at least 50% of the subjects for the further analysis. We also called germline variants in cohort B and C and filtered variants genotyped in >= 50% subjects, and identified ∼2.5 million germline variants per sample. The number of germline variants called per sample depends on sequencing depth; with deeper-coverage samples generally exhibited higher variant counts. The transition frequency (∼72%) was slightly higher in germline variants compared to somatic variants (**Supplementary table 3**). In total, C>T and A>G variants were the most frequent, each accounting for ∼35% of all mutations. Higher frequency of A>G could be due to less evolutionary pressure because A>G often results in synonymous variants due to codon wobble.

### Somatic mutations

To characterize the somatic mutation landscape in HNSCC patients, we integrated Cohorts A, B, and C to enhance statistical power for detecting driver genes. Subsequent comparison with TCGA datasets (cohort D) aimed to uncover novel mutations and evaluate shared mutational patterns.

We identified 41 and 27 significantly mutated genes in two different statistical frameworks: mutSig2CV and dNdscv (**Figure 1A, Supplementary table 4 and 5**). In brief, MutSig2CV identifies recurrently mutated genes by modeling background mutation rates and covariates like expression of genes and DNA replication rate, while dNdScv quantifies selection pressure via dN/dS ratios to detect driver genes [24, 25]. As expected, we identified enrichment of gene-sets associated with head and neck cancer (WP4674, FDR = 1.4×10^-3^; DOID: 5520, FDR = 2.6×10^-4^) (**Supplementary table 6**).

**Figure 1.**
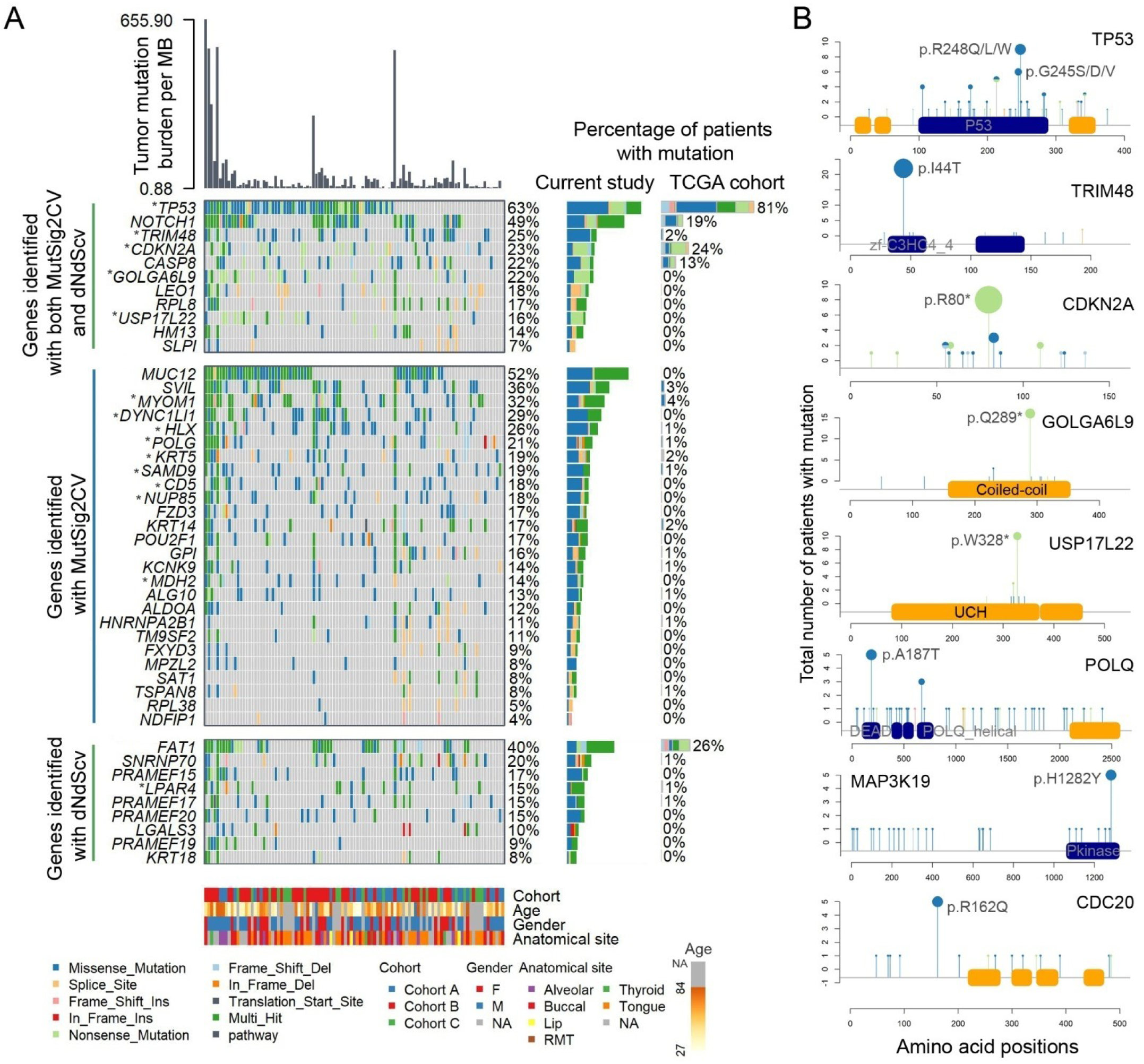
A. Oncoplots depicting the somatic mutation landscape across patient cohorts. Each column represents an individual patient, and each row corresponds to a gene. Colors denote mutation types (e.g., missense, nonsense, frameshift, splice site). The top bar plot shows the overall mutation burden per patient per megabase-pairs (MB). The side bar plot illustrates the proportion of mutations detected per gene, collectively for Cohorts A, B, and C, as well as for Cohort D (TCGA) (HPV-negative patients).Recurrently mutated genes with hotspot mutations were highlighted with asterisk sign (“*”). **B.** Lollipop plot of the selected set of genes to show hotspot mutations. Each panel depicts the somatic mutation distribution for a recurrently mutated gene with hotspot mutation/s. Protein domains, family and motifs are represented along the x-axis with annotated amino acid positions. Domains are in blue color while motifs and families are in the green color rectangle boxes. Circles indicate individual mutation sites, with colors corresponding to mutation types (*e.g.*, frameshift deletions, nonsense mutations, missense mutations). Different types of mutation at the same site were represented as a pie chart instead of filled circle. The y-axis shows the number of patients carrying each mutation. Frequently recurring hotspot mutation/s is labeled in each panel.

In total, eleven genes (*TP53*, *CDKN2A*, *CASP8*, *HM13*, *RPL8*, *SLPI*, *TRIM48*, *NOTCH1*, *GOLGA6L20*/*L9*, *USP17L22/USP17* and *LEO1*) were common in both framework and mutated in >5% of patients. This list overlaps with cohort D where *TP53*, *CDKN2A*, *CASP8* and *NOTCH1* were significantly mutated genes [7]. Like TCGA (cohort D), *TP53* is highly mutated genes in our cohorts (A, B and C) with ∼63% patients with mutation in *TP53*. The ∼56% of the total mutations present in *CDKN2A* were truncating mutations in Indian cohort (A, B and C) similar to cohort D (**Supplementary table7**).While the overall distribution of variant types (*e.g.* truncating or missense mutations) did not differ (p value ∼ 1) between our cohort and TCGA for *NOTCH1* gene, but number of the mutations were significantly high in Indian cohort, occurring in ∼49% of patients. The ≥ 5% patients of our cohort (A, B and C) were having mutation at the same genomic location in *TP53*, *CDKN2A*, *TRIM48*, *GOLGA6L20/L9* and *USP17L22/USP17* (**Figure 1B**) making it hotspot mutations. Of these, TRIM48 is novel with p.I44T mutation in zinc-finger (zf-C3HC4_2) domain (**Figure 1B**). This mutation was found in ∼21% of the patients of our cohort (A, B and C) (**Table 2**). This indicates that p.144T may be a driver mutation in tobacco-chewing patients or those with clinical characteristics similar to the patients of cohort A, B and C as p.144T was not found in cohort D. Another novel gene is *GOLGA6L20/L9* with hotspot mutation “p.Q289*” (**Figure 1B**). This hotspot mutation introduces truncation in the GOLGA6L20/L9 protein and present in 14 (∼14%) patients. *USP17L22*/*USP17* was highly significant and mutated in ∼16% of the patients. Most of the mutations were concentrated within 316^th^ and 342^th^ amino-acids in Peptidase_C19 protein-domain of USP17L22/USP17. Of which p.W328* truncating mutation was present frequently and found in ∼10% of total patients (**Figure 1B**).

**Table 2.**
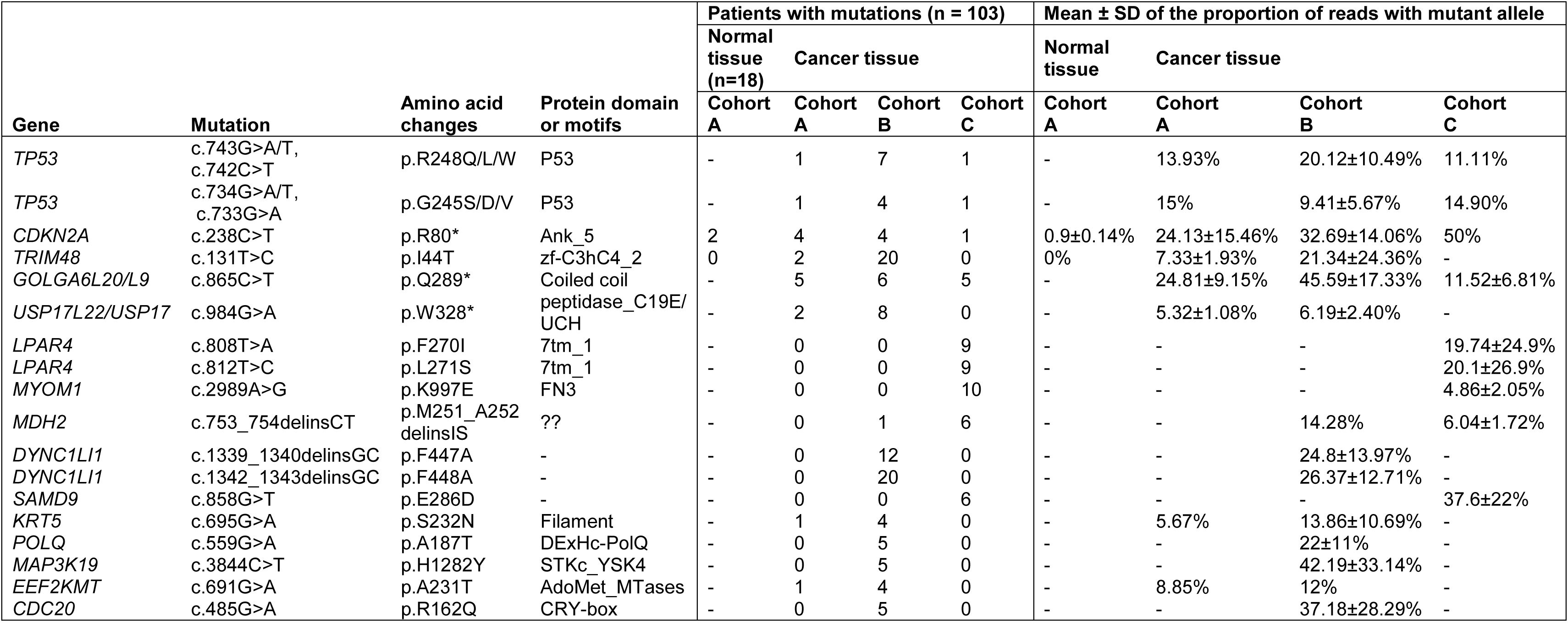
Summary of hotspot mutations in recurrently mutated genes in normal and cancer tissues of 103 patients across different cohorts.The associated gene, chromosomal position (hg38), nucleotide and amino acid changes, protein domain, the number of patients harboring the mutation, and the mean ± standard deviation of the proportion of reads carrying the mutation are listed for each mutation.

Using statistical framework of dNdScv, we identified additional genes (**Supplementary table 4 and 5**). The *FAT1* gene which is second significantly highly mutated gene in TCGA cohort (cohort D) was identified in ∼40% of total South Asian patients (Cohort A, B and C) exclusively using statistical framework of dNdScv. It indicates that like TP53, *FAT1* gene is common driver gene in the South Asian cohorts compared to Caucasian cohort which is having different exposure. Another significantly mutates gene is *LPAR4* which has most of the mutations concentrated at 270^th^ and at 271^th^ amino-acid (p.F270I and p.L271S). Both hotspot mutations were present in the∼9% of the patients in our cohort. This mutation was absent in the cohort D. On the other hand using statistical framework of mutSig2CV, additional 31 significantly and recurrently mutated genes were identified. Of which, mutations at the same genomic location were present in at least∼5% of total patients in*MYOM1*, *MDH2*, *CD5*, *DYNC1LI1*, *NUP85*, *SAMD9*, *HLX* and*KRT5* genes (**Supplementary figure 1**). It includes p.K997E (present in FN3 protein of MYOM1), p.M251I-A252S (MHD glyoxysomal mitochondrial domain of MDH2), p.H461C (C-terminal region of CD5), p.F447A-F448A (C-terminal region of DYNC1LI1), p.D555N (Nucleopor_Nup85 domain of NUP85), p.E286D (N-terminal region of SAMD9), p.A235V (mid-region of HLX) and p.S232N (Filament domain of KRT5) mutations which were present in ∼10%, ∼7%, ∼10, ∼19%, ∼7%, ∼6%, ∼15% and ∼5% of total patients. None of these mutations were reported in the TCGA cohort (cohort D).

Some genes that were significantly and recurrently mutated in the TCGA cohort (cohort D) and present in at least ≥5% of the patients were not identified in South Asian cohorts (A, B and C). For instance, *KMT2D*, which is mutated in approximately 16% of TCGA patients (cohort D) and predominantly harbors truncating mutations (∼66%; 43 of 65 patients harboring *KMT2D* mutations), was mutated in ∼45% of patients in the our cohort, but harbors truncating mutations only in ∼30% patients (14 out of 46 patients harboring *KMT2D* mutations). The statistical framework of the both MutSig2CV and dNdScv indicated that KMT2D in South Asian cohort are not recurrently mutated and the observed mutation spectrum are likely attributable to random chance..*AJUBA* had a high proportion of truncating mutations (∼75% of total mutations) in TCGA but was not mutated in South Asian cohort. *PIK3CA* displayed hotspot mutations in TCGA—p.E542K (11 patients), p.E545K (∼4%; 16 patients), and p.H1047R (∼3%; 12 patients)—whereas in South Asian cohort, only one patient had Glu542Lys and two had Glu545Lys mutations. *NSD1*, which was predominantly truncating (∼75%; 38 of 51 patients) in TCGA (cohort D), was not significantly mutated in present cohort. Similarly, *FBXW7* showed a contrast in truncation frequency (∼38%; 11 out of 29 patients in TCGA *vs.* ∼13%; 1 out of 8 patients in South Asian cohort). *HRAS* had hotspot mutations at p.G12 (∼46%; 13 out of 28 patients) and p.G13 (∼39%; 11 out of 28 patients) positions in TCGA (cohort D) but none of patients in present cohort had mutations in this gene. *NFE2L2*, though significantly mutated in TCGA (cohort D) (without clear hotspot or truncating mutation bias), was not significantly mutated. Lastly, *TGFBR2* was significantly mutated in TCGA with a higher frequency of truncating mutations (∼33%; 7 out of 21 patients), but again, it was not significantly mutated. This suggests that the TCGA cohort may include a specific subset of patients with distinct underlying biological mechanisms that are not represented in the South Asian cohort. However, it should be noted that except for KMT2D, subset of patients with mutation in these genes were relatively rare in the TCGA cohort, and our sample size may not be sufficiently powered to reliably detect such low-frequency variants.

### Hotspot somatic mutations

Mutations originate in non-germline tissues do not pass from one generation to the next. We anticipated that hotspot mutations in South Asian cohorts may differ from those observed in Caucasian dominated cohorts like TCGA. To investigate this, we employed two complementary approaches: (1) identifying *de novo* hotspot mutations, and (2) analyzing the frequency spectrum of previously reported hotspot mutations.

After filtering *de novo* hotspot mutations that were statistically significant and present in at least 5 patients (∼5% patients of South Asian cohort), we identified 44 genes (**Supplementary table7**). TP53 was one of them with two frequent mutations: p.R248Q/L/W and p.G245S/D/V, present in 9 and 6 patients, respectively (**Table 2**). The hotspot mutation which was present in most of the patients in South Asian cohort was in DYNC1LI1. The hotspot mutations p.F448A (20 patients:∼19%) and p.F447A (12 patients: ∼12%) were present at the C-terminal region of protein (**Figure 1B**). DYNC1LI1is involved in chromosomal segregation. It has been reported that F447 and F448 mutations present in the helix-1 of LIC1/DYNC1LI1 and present in evolutionary conserved regions. These mutations can abrogate binding of DYNC1LI1 with several adaptor proteins [26, 27]. The other important gene with hotspot mutation was POLQ. Out of 6 patients of cohort B which were having highly tumor mutation burden, 5 patients were having same hotspot mutation “p.A187T” in POLQ (**Table 2**). The POLQ is involved in DNA repair pathways and its defect leads to the high mutation burden patients as expected. The p.A187T present in the DExHc-PolQ domain. The hotspot mutation “p.Q289*” present in GOLGA6L20/L9 was also identified. This mutation was present in 14 (∼14%) patients. Another hotspot mutation “p.H1282Y” was present in the C-terminal of MAP3K19 in the protein-kinase domain (STKc_YSK4). The hotspot mutation “p.A231T” was present in the AdoMet_MTases protein domain of EEF2KMT and might be functionally significant in the HNSCC cancer. The “p.R162Q” is present in the CRY-box of CDC20 and was identified in the 5 patients (**Supplementary table7)**. Hotspot mutations p.R248 and p.G245 of TP53 were present in the TCGA datasets (cohort D) in ∼3% and ∼4% patients. On the other hand none of the hotspot mutations were present in LIC1/DYNC1LI1, POLQ, GOLGA6L20/L9, MAP3K19 and CDC20 in TCGA cohort.

In the next step, we analyzed the frequency spectrum of known hotspot mutations. Using same filter *i.e.* presence of same mutation in at least in 5 patients and additional filter *i.e.* at least 10% of the reads should have hotspot mutations, we identified 11 genes (**Supplementary table 8**). Top three genes with hotspot mutations were: CTLA4 (p.T17A in 59 patients, ∼57%), HNF1A (p.I27L in 65 patients, ∼63%) and *MYC* (p.T73P in 53 patients, ∼52%) (**Table 3** and **Supplementary table 8**). The CTLA4 is an immune checkpoint regulator and HNF1A is a transcription factor involved in metabolic regulation. The p.T73P hotspot mutation in MYC was present in higher frequency in South Asian cohort compared to TCGA cohort. In the TCGA-cohort, MYC is mutated in 14% subject, but none of them were having p.T73P hotspot mutations. Between 11 to 26% of the total reads mapping to the genomic location of p.T73P were carrying mutant allele. The F-box protein FBW7 phosphorylates MYC at p.T73 which results in the degradation of MYC protein [28]. Constitutively presence of active in MYC might be the reason of tumor cell proliferation. The other known hotspot mutations, p.A1185D (∼18% of patients) and p.R80* (∼8% of patients) were present in INPPL1 and CDKN2A (**Table 2**, **Table 3** and **Supplementary table 8**). These mutations were present in ≥ 10% of total reads (p.A1185D: ∼10%-25% and p.R80*: ∼16%-56%). The INPPL1 is involved in PI3K signaling pathways, but functional significance of p.A1185D is unknown. On the other hand, p.R80* results in the premature truncation of CDKN2A and thus, loses binding with CDKs [29]. It leads to uncontrolled cell proliferation. The other known hotspot mutations in CDKN2A *e.g.* p.R58*, p.S12*, p.W110*, p.H83Y and p.P11H were also present in 9 (∼9%) patients. It is important to note that most of the observed known hotspot mutations in CDKN2A were truncating mutations. Similar to CDKN2A, many different known hotspot mutations of TP53 were found in the 59 (∼57%) patients in the South Asian cohort, but only one hotspot mutation “p.R248Q/W” was present in 8 patients (**Supplementary table 8**). Several known hotspot mutations of PIK3CA were also observed in 12patients (∼10%), but none of mutations were present in more than 2 patients. The other hotspot mutations were also present in >= 5 patients having at least 10% of reads with mutant alleles: p.T96S (∼9% patients, GNAQ), p.G1057D (∼44%, IRS2), p.V12G (∼15%, KNSTRN) and p.P479_A480dup (∼18%, LATS2).

**Table 3.** Summary of known hotspot mutations in normal and cancer tissues of 103 patients across different cohorts.The associated gene, chromosomal position (hg38), nucleotide and amino acid changes, protein domain, the number of patients harboring the mutation, and the mean ± standard deviation of the proportion of reads carrying the mutation are listed for each mutation.

| Gene | Mutation | Amino acid changes | Protein domain or motifs | Patients with mutations (n = 103) | | | | Mean ± SD of the proportion of reads with mutant allele | | | |
| --- | --- | --- | --- | --- | --- | --- | --- | --- | --- | --- | --- |
|  |  |  |  | Normal tissue (n=18) | Cancer tissue |  |  | Normal tissue | Cancer tissue |  |  |
|  |  |  |  | Cohort A | Cohort A | Cohort B | Cohort C | Cohort A | Cohort A | Cohort B | Cohort C |
| TP53 | c.743G>A/T, c.742C>T | p.R248Q/L/W | P53 | 0 | 1 | 7 | 1 | - | 13.93% | 20.12±10.49% | 11.11% |
| TP53 | c.734G>A/T, c.733G>A | p.G245S/D/V | P53 | 0 | 1 | 4 | 1 | - | 15% | 9.41±5.67% | 14.90% |
| CDKN2A | c.238C>T | p.R80* | Ank_5 | 2 | 4 | 4 | 1 | 0.9±0.14% | 24.13±15.46% | 32.69±14.06% | 50% |
| CTLA4 | c.49A>G | p.T17A | - | 16 | 15 | 27 | 17 | 55.26±18.75% | 52.9±14.11% | 56.89±22.96% | 57.39±20.71% |
| HNF1A | c.305A>C | p.I27L | Hnf-p1 | 15 | 15 | 34 | 16 | 73.05±28.37% | 73.33±25.91% | 60.15±27.57% | 64.4±31.58% |
| MYC | c.580A>C/G | p.T73P | Myc_N | 0 | 0 | 32 | 21 | - | - | 14.98±2.44% | 17.89±6.68% |
| INPPL1 | c.3772A>C | p.A1185D | - | 0 | 0 | 4 | 15 | - | - | 18.13±10.49% | 15.69±3.76% |
| GNAQ | c.862T>A | p.T96S | G_alpha | 2 | 4 | 5 | 0 | 11.75±1.77% | 11.28±1.33% | 18.7±12.05% | - |
| IRS2 | c.3700C>T | p.G1057D | - | 11 | 13 | 27 | 5 | 61.26±17.89% | 54.83±16.76% | 62.07±20.76% | 67.5±24.51% |
| KNSTRN | c.149T>G | p.V12G | - | 0 | 0 | 0 | 15 | - | - | - | 16.11±4.09% |
| LATS2 | c.1881->GGGGCG | p.P479_A480dup | - | 15 | 18 | 0 | 0 | 29.29±19.09% | 26.24±15.21% | - | - |

To investigate how the mutation spectrum at the known hotspot regions may have evolved, we compared the frequency of reads containing mutant alleles in both cancer tissues and in matched adjacent normal tissue samples. This analysis was feasible in Cohort A due to the availability of corresponding normal tissue samples, providing a significant advantage over other cohorts (B and C). Here, we consider only those known hotspot variants which were present in at least 5% of the patients. For most known hotspot mutations, the proportion of cancer tissues harboring a known hotspot mutation was comparable to that observed in normal adjacent tissues (**Supplementary Table 8** and **Table 3**). Same was true for the proportion of mutation carrying reads (**Table 3**). This balanced distribution suggests that these mutations (*i.e.*p.T17A, p.I27L, p.T96S, p.G1057D and p.P479_A480dup) may not play a significant role in cancer development. On the other hand, the proportion of reads carrying p.R80* (CDKN2A) was increased from ∼1% in normal tissues to ∼24% in cancer tissues which suggest that p.R80* might be important in cancer development and is oncogenic driver mutation. A similar role may also apply to variants p.R248Q/L/W (TP53), p.G245S/D/V (TP53), p.T73P (MYC), p.A1185D (INPPL1) and p.V12G (KNSTRN). However, some of these missense mutations result in the substitution of amino acids with others from the same class. For example, valine (V) and glycine (G) both have hydrophobic side chains. Such substitutions may not significantly alter the protein’s function. But, why these mutations enriched in tumor tissues are subject to further study. Since, several hotspot mutations were present in the TP53 and PIK3CA; we explored all hotspot mutations collectively, and found that reads carrying hotspot mutations were increased in cancer tissue samples.

### Copy-number variations

Copy number variations (CNV) analysis was performed in two separate steps. In the first step, CNVs were identified in tumor tissues from cohort A by comparing them with their corresponding adjacent normal tissues. In the second step, five adjacent normal tissues were randomly selected, and copy number variations in cohorts B and C were called against this combined set of normal tissues as a reference.

In total, 3, 34, and 46 genomic regions were amplified, while 9, 41, and 18 genomic regions showed copy number loss in cohorts A, B, and C, respectively (**Figure 2 and Supplementary table 9**; manhattan plot for the copy number gain of few selected samples are show in **Supplementary figure 2**). For cohorts B and C, we only considered those CNVs that overlapped with cohort A. The 7p11.2 (q-value = 3.5×10^-5^; 8 out of 24 patients) and 9p13.3 (q-value = 3×10^-2^; 5 out of 24 patients) was amplified only in cohort A, while 11q13.2 was amplified in cohort A (8 out of 24 patients; q-value = 1×10^-6^) and in cohort B (23 out of 51 patients; q-value = 3.7×10^-11^). The 7p11.2 region contains several genes. Among these, the *EGFR* appears to be particularly important. Amplification of the *EGFR* gene has been frequently reported in HPV-negative head and neck squamous cell carcinoma (HNSCC), including in patients from the TCGA cohort (cohort D) [7]. In addition, a recurrent copy number gain at 11q13.2 has also been documented in several studies, often occurring alongside a deletion at 11q22[7]. However, in our analysis, this pattern was not observed. Notably, previous studies reporting amplification at 11q13.3 have identified concurrent amplification of the FADD gene. In contrast, our data did not show amplification of the genomic region encompassing FADD in significant number of samples. Only one sample was having copy number gain of region containing FADD gene. Instead of 11q13.3, we observed amplification of 11q13.2 cytoband, encompassing *ALDH3B1*, *CHKA*, *NDUFS8*, *TCIRG1*, and *UNC93B1*, which may represent alternative drivers in our cohort. The 11q13.3 gain, which includes the *CCND* gene, was detected only in cohort C.

**Figure 2.**
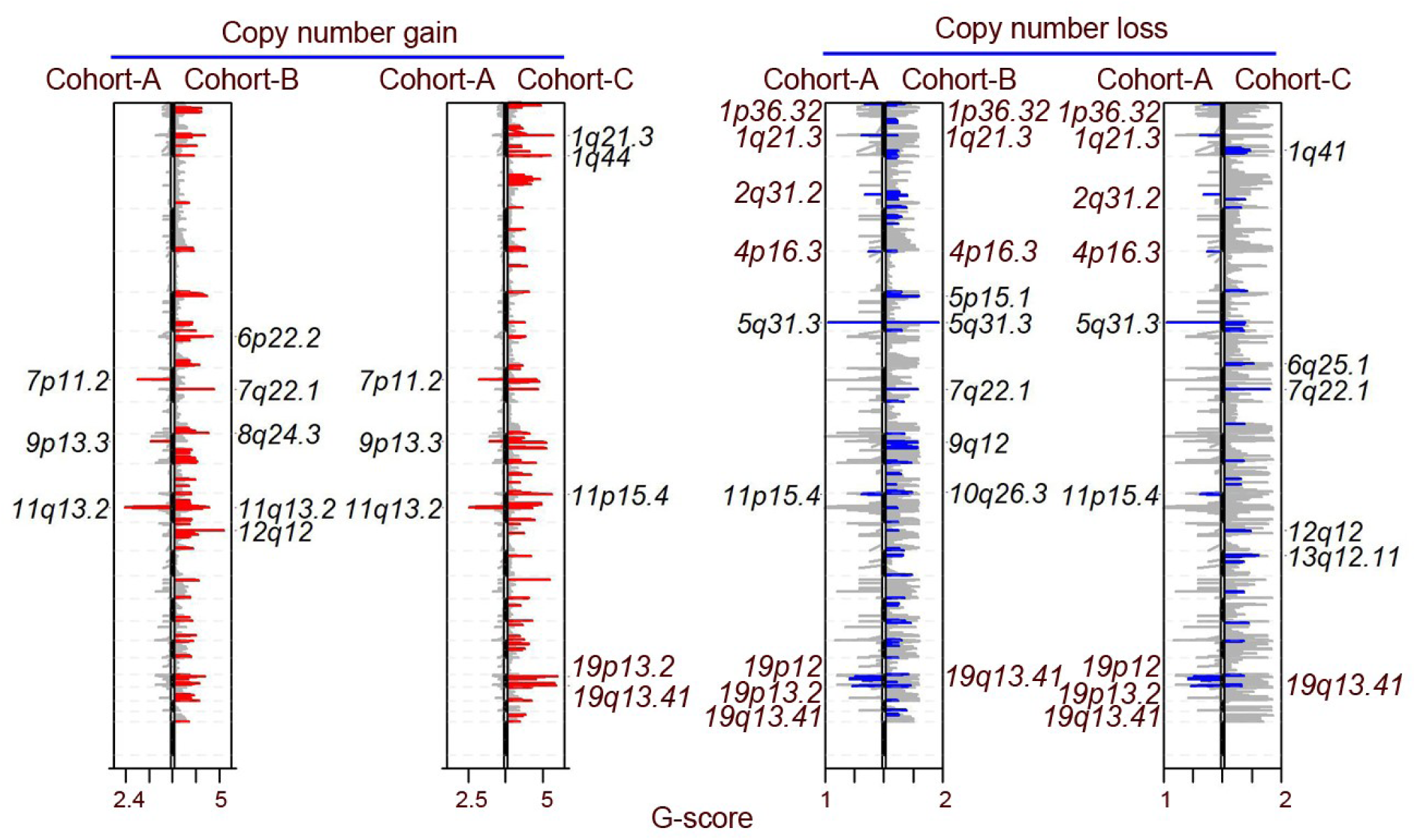
Copy number variations (CNVs) identified in patients from Cohorts A, B, and C. Panels show the GISTIC plot illustrating both copy number gains and losses. In each subpanel, CNVs in Cohorts B and C are illustrated and compared with Cohort A, for which CNVs were determined relative to matched normal adjacent tissues.

Among the nine genomic regions with copy number loss identified in cohort A, one region—19q13.41—exhibits copy number loss across all three cohorts. This CNV was observed in 8 out of 24 patients in cohort A (q-value = 1.26×10^-5^), 26 out of 51 patients in cohort B (q-value = 1.4×10^-10^), and 1 out of 23 patients in cohort C (q-value = 3×10^-2^). Most of the genes within this region encode Zinc Finger proteins (**Supplementary table 9**). Besides this, four regions with copy number loss overlapped between cohorts A and B. Of which, 1p36.32 (cohort A: 3 out of 24 patient, q-value = 3.3×10^-2^; cohort B: 19 out of 51 patients, q-value = 1.27×10^-6^) appears to be particularly important as TP73 present in this cytoband. The other three regions with copy number loss were 1q21.3 (cohort A: 3 out of 24 patient, q-value = 4.1×10^-3^; cohort B: 11 out of 51 patients, q-value = 1.19×10^-2^), 4p16.3 (cohort A: 3 out of 24 patient, q-value = 4.8×10^-2^; cohort B: 17 out of 51 patients, q-value = 2.1×10^-2^) and 5q31.3 (cohort A: 9 out of 24 patient, q-value = 6.04×10^-13^; cohort B: 39 out of 51 patients, q-value = 4.33×10^-44^) (**Figure 2** and **Supplementary table 9**). Among these regions, the copy number loss at 5q31.3 was the highly significant. The high frequency and strong statistical significance suggest that this region may play a critical role in tumor development or progression. The overlapping segment of 5q31.3 affected in both cohorts contains a cluster of protocadherin beta (PCDHB) genes (**Supplementary table 9**).

### Germline mutations

To investigate the genetic risk factors for HNSCC in South Asian cohort, we began by analyzing cohort A, which contains exome sequencing data from normal tissue samples. First, we assessed the relatedness among the samples using the unadjusted Ajk statistic and found thatnone of the samples were related to each other. We then identified genetic variants with a frequency of at least 5% that were not missing in more than 50% of the samples. It is equivalent to those variants present in at least ∼2 out of 28 normal adjacent tissues. It results in 9.5 million variants. This set of variants includes putative non-functional variants also. Therefore, in the next step, we filtered those variants which were putative function (*e.g.* missense, stop loss/gain, start loss, inframe/frameshift, splice site donor/acceptor variants) and were having frequency ≤ 10^-3^ in gnomAD and 1000 genome project samples for further analysis. To find true positive putative functional variants, we selected only those variants which were common among cohort A, B and C. During this filtering, variants present in mitochondrial chromosome were removed. We also manually removed variants that were flagged as unsupported in the dbSNP database or located in the local repetitive regions.

In total, seven germline single nucleotide variants (SNVs) were identified which were present in heterozygous state in all patients. All variants were present in the protein-domain: (1) p.V446A and p.S456L were present in FAD/NAD-binding domain of the SDHA (succinate dehydrogenase complex flavoprotein subunit A) protein, (2) p.Q525R and p.G532S were present in the Cadherin_repeats domain of PCDHB3, and (3) p.M691V, p.A692V and p.S693R were present in the EGF_CA domain of MEP1A (**Table 4**). The genomic region (chr5:236504 and 236534) having enough read depth in 19 patients from Cohort A, with 7 (∼37%, 95% CI: 19–59%) patients harboring the p.S456L variant in heterozygous form. In Cohort B, screening of 51 samples revealed 12 patients (∼24%, 95% CI: 14-37%) carrying heterozygous variant. Besides this, p.S456L was also detected in 5 patients in heterozygous condition (∼22%, 95% CI: 10-42%) in Cohort C. The p.V446A variant was observed at a lower frequency compared to p.S456L (**Table 4**), and it did not result in a change in amino acid properties (both “V” and “A” amino-acids are hydrophobic). It is possible that the increased presence of p.V446A in patients is due to hitchhiking, as all individuals carrying p.V446A also carried p.S456L, suggesting strong linkage disequilibrium between these two variants(D’ = 1 and r2 = 0.47; **Table 4**) (**Supplementary figure 3**). Similar to the variants observed in SDHA, p.Q525R and p.G532S were also in the the stong linkage disequilibrium and all patients carrying p.G532S also carried p.Q525R. Given the increased frequency of p.G532S could be a result of hitchhiking. All three variants (p.M691V, p.A692V and p.S693R) were also linked with each other and present in the same set of patients. Among them, only p.S721R results in a change in amino acid class, while the other two do not alter amino acid properties. To compare with the TCGA datasets, we extracted 853 germline putative functional variants which were reported elsewhere [12]. In total, 9 variants in SDHA were reported in other cancer types in TCGA patient cohort, but none of the variants were reported in HNSCC and none of them were p.S456L.

**Table 4.** Distribution of germline variants identified in patients from Cohorts A, B, and C, including their chromosomal positions (hg38), reference and alternate alleles, and the corresponding amino acid changes. Linked variants were annotated using COSMIC (COSV) and dbSNP (rsID) databases. The corresponding genes and the number of patients harboring each variant in each cohort are indicated. Linkage disequilibrium metrics are provided to reflect the strength of association between variants.

| Chr | Genomic location (hg38) | Alleles | Amino acid change | Linked variants | Gene | Number of patients with mutations in each cohorts |  |  | Linkage disequilibrium (D'; r<sup>2</sup>; LOD score) |
| --- | --- | --- | --- | --- | --- | --- | --- | --- | --- |
|  |  |  |  |  |  | Cohort A | Cohort B | Cohort C |  |
| 5 | 236504 | T/C | p.V446A | rs201741295,COSV53770276 | SDHA | 7 | 2 | 3 | 1; 0.466; |
| 5 | 236534 | C/T | p.S456L | rs76896145,COSV53768124 | SDHA | 7 | 12 | 5 | 8.49 |
| 5 | 141102223 | A/G | p.Q525R | rs1588306958,COSV50563932 | PCDHB3 | 16 | 8 | 2 | 1; 0.76; |
| 5 | 141102243 | G/A | p.G532S | rs1751970029 | PCDHB3 | 10 | 7 | 1 | 13.22 |
| 6 | 46835536 | A/G | p.M691V | COSV57919076 | MEP1A | 3 | 4 | 1 |  |
| 6 | 46835540 | C/T | p.A692V | rs752666180,COSV57918539 | MEP1A | 3 | 4 | 1 | 1; 1.0; |
| 6 | 46835544 | C/G | p.S693R | - | MEP1A | 3 | 4 | 1 | 11.92<sup>†</sup> |

<sup>†</sup>For *MEP1A* variants (p.M691V, p.A692V, and p.S693R), identical LD values were observed for all pair-wise combinations *i.e.* p.M691V vs. p.A692V, p.M691V vs. p.S693R and p.A692V vs. p.S693R.

Although the observed difference did not reach statistical significance (p value = 0.07), potentially due to the limited sample size, there was a trend suggesting that patients harboring the p.S456Lmutation in SDHA were younger than those carrying the wild-type allele. The mean age of patients with the mutation was 52.43±9.52 years compared to 62.82±13 years in patients without the mutation. The other variants *i.e.* p.Q525R and p.S693R did not show any trend towards age (p value = 0.25 and 0.11, respectively).

## Discussion

The genomic landscape of head and neck squamous cell carcinoma (HNSCC) is marked by striking molecular heterogeneity, driven by divergent etiological factors such as HPV infection, tobacco use, and might be ethnic-specific exposures like betel nut and tobacco chewing[7, 30].

By prioritizing genomic profiling of underrepresented HPV-negative cohorts with betel nut and tobacco chewing exposure, we can uncover ethnicity-specific driver mutations, refine risk stratification, and expand the repertoire of actionable targets for precision oncology. Considering this, we recruited patients from the hospital and performed somatic mutation profiling using massively parallel sequencing. To increase statistical power, we included somatic profiling of the additional patients from the same geographical region. Consistent with previous reports, we observed a predominance of C>A transversions, a known mutational signature associated with tobacco exposure. Additionally, the study cohort had a higher proportion of male patients, aligning with established gender distributions in HNSCC epidemiology [1]. Most samples were obtained from patients in the early stages of the disease, suggesting that the identified variants and driver genes may reflect early molecular events in tumorigenesis. Our cohort differs from the Indian HNSCC patients of ICGC cohort, where approximately 86% of cases are diagnosed at advanced clinical stage (Stage IV). Although both groups share similar ethnicity-specific exposures such as betel nut and tobacco chewing, ∼26% of the ICGC patients were HPV-positive and had a younger average age (mean: 48 years) compared to our South Asian cohort and the TCGA cohort[8]. These distinctions suggest the potential to identify novel driver genes unique to our cohort, while also observing overlap with known oncogenic drivers with patients from the ICGC cohort. This is likely to hold true for comparisons with the TCGA-HNSCC cohort as well, which is characterized by distinct clinical and demographic features.

In the present study, we identified <10% patients with high tumor mutation burden in our South Asian cohort. Tumor mutation burden (TMB) has shown some correlation with immunotherapy response. In this study, we report for the first time a recurrent hotspot mutation (p.A187T) in the *POLQ* gene, identified in our patient cohort having high tumor mutation burden. This variant was absent in both TCGA and ICGC cohorts, but has been reported in few cases with elevated level of TMB in uterine and tubo-ovarian carcinomas [31]. This finding aligns with POLQ’s established role in DNA damage repair, where dysfunction can drive hypermutation [32, 33].

The driver genes identified in the TCGA cohort, including *CDKN2A*, were also identified as driver genes in the patients of our cohorts having many truncating mutations. However, no truncating mutations in *CDKN2A* were found in the Indian HNSCC patients from the ICGC cohort—only copy number losses were detected in ∼10% of cases. This discrepancy may be attributed to the higher proportion of HPV-positive cases in the ICGC cohort, as HPV-positive tumors in Caucasian populations (such as those in the TCGA cohort) also showed no *CDKN2A* mutations or copy number losses. The *CDKN2A* gene, which encodes the critical cell cycle regulator p16/INK4A, has been strongly linked to poor survival outcomes in oral cancers, particularly in HPV-negative contexts where its loss is more prevalent. The recurrent copy number losses and truncating mutations in *CDKN2A* could potentially serve as a biomarker for adverse prognosis, but, further validation studies are needed to confirm this association and establish its clinical utility. In oral squamous cell carcinoma, *CDKN2A* inactivation—whether through deletion, mutation, or epigenetic silencing—is frequently associated with disease progression and treatment resistance, underscoring its role as a key molecular driver in this malignancy [34].

Comparative genomic analyses have identified recurrent mutations in epigenetic regulators—including *ARID2*, *EP300*, *KMT2B/MLL2*, *KMT2D/MLL4*, *NSD1* and *TET1*—in Indian HNSCC patients from the ICGC cohort, while *KMT2D*/*MLL4* and *NSD1* mutations were prevalent in the TCGA cohort. In contrast, our South Asian cohort lacked significantly mutated epigenetic factors. Although *KMT2D* mutations were observed in ∼30% of patients, this gene did not reach statistical significance, suggesting that it may represent stochastic events rather than driver alterations. These findings imply that tumorigenesis in our cohort may not be epigenetically driven. However, further functional studies assessing histone modifications are warranted to validate this hypothesis. An alternative explanation could be the early clinical stage of our cohort’s patients, as epigenetic mutations may arise later in disease progression. Notably, this contrasts with other malignancies such as bladder cancer, where epigenetic dysregulation often occurs early, followed by secondary driver mutations [35]. Whether this paradigm applies to HNSCC—or whether our cohort represents a distinct molecular subset—remains an open question requiring further investigation. EGFR and its downstream effectors (*e.g. HRAS*, *KRAS*) frequently acquire copy number gains or activating mutations in HNSCC (cohort D). However, in South Asian cohort, neither *HRAS* nor *KRAS* emerged as significantly mutated driver genes. However, in the copy number variation analysis, we identified amplification of *EGFR* gene in ∼30% of the patients which was comparable with the proportion of patients with amplification in *EGFR* in TCGA cohort. It suggests that South Asian patients can get benefited from the cetuximab treatment and percentage of the responder could be same as Caucasian patients. Cetuximab targets a binding pocket in the EGFR protein. We have previously identified a mutation, p.G465R, in the *EGFR* gene, which is predicted to prevent cetuximab from binding to EGFR [36]. Therefore, sequencing the EGFR binding pocket prior to cetuximab treatment is highly recommended for patients with EGFR amplification.

Consistent with findings from The Cancer Genome Atlas (TCGA) and the International Cancer Genome Consortium (ICGC) cohorts of Indian head and neck squamous cell carcinoma (HNSCC) patients, our analysis revealed several commonly mutated driver genes, including *TP53*, *CASP8*, and *NOTCH1*. These genes are frequently altered in HNSCC patients, supporting their fundamental role in tumor development and progression. However, we did not detect mutations in a subset of driver genes previously reported in the TCGA cohort, such as *AJUBA*, *PIK3CA*, *FBXW7*, *NFE2L2*, and *TGFBR2[7]*. The absence of these alterations in our cohort may reflect differences in ethnic specific exposure or biological variability. Notably, in the TCGA dataset (Cohort D), these genes were mutated in only a small proportion of patients, suggesting that their detection may require larger sample sizes to achieve sufficient statistical power. Therefore, the limited cohort size in our study could have constrained our ability to identify less frequently mutated driver genes. Further studies involving larger, well-characterized cohorts are necessary to validate these findings and to better understand the landscape of both common and rare driver mutations in HNSCC, particularly within South Asian populations. Furthermore, besides the well-established drivers, we report novel driver genes identified in the current study, including *TRIM48* which has not been previously associated with HNSCC. TRIM48 is an E3 ubiquitin ligase that promotes K48-linked polyubiquitination and degradation of PRMT1—an enzyme that suppresses the ASK1 kinase—thereby enhancing ASK1-mediated apoptotic signaling under oxidative stress[37]. In experimental cancer models, overexpression of TRIM48 increased oxidative stress–induced cell death, while knockdown attenuated ASK1 activation and cell death. This functional profile suggests TRIM48 may play a previously under recognized role in modulating stress-induced apoptosis in tumor cells and constitutes a compelling novel candidate driver in HNSCC patients - particularly in South Asian populations with distinct ethnic-specific exposures.

Among the additional potential driver genes identified in our study are CDC20 and MAP3K19, which harbor the hotspot mutations p.R162Q and p.H1282Y, respectively. The p.R162Q variant in CDC20 is located within the CRY box, a previously characterized degron motif that is critical for proper spindle assembly checkpoint (SAC) function [38]. The CRY box promotes the formation of the mitotic checkpoint complex (MCC) and facilitates its interaction with the anaphase-promoting complex/cyclosome (APC/C). Mutation at this site, such as p.R162Q, may impair checkpoint fidelity and potentially abrogate normal cell cycle control, contributing to genomic instability and tumor progression [38]. Although the functional impact of the p.H1282Y mutation in MAP3K19 remains unclear, MAP3K19 has recently been identified as a novel regulator of MAPK signaling. It activates both ERK and JNK pathways, including via a RAF-independent mechanism. Biochemical assays show that MAP3K19 directly phosphorylates MEK and MKK7, and RNAi screens indicate it is essential for KRAS-mutant cancer cell survival. These findings suggest MAP3K19 as a potential therapeutic target in oncogene-driven tumors [39]. However, it is important to note that in our cohort, we did not detect any activating mutations in KRAS or HRAS, but observed high-level EGFR copy number gains, suggesting alternative mechanisms of MAPK pathway activation in these patients.

Of the known hotspot mutations identified in current study, p.T73A in MYC stands out as particularly interesting, as it was detected in multiple patients within our cohort. This mutation has been previously characterized and is known to enhance MYC protein stability by reducing its ubiquitination [40]. Specifically, phosphorylation of the highly conserved threonine 73 (T73) residue normally promotes ubiquitin-mediated degradation of the MYC protein. Substitution of this residue, as in the T73A mutation, is therefore predicted to impair degradation, resulting in increased MYC protein stability [41]. The MYC oncogene is a central regulator of numerous pathways involved in tumorigenesis, including cellular proliferation, inhibition of cell cycle exit, promotion of angiogenesis, and enhancement of genomic instability [42–45]. Despite its critical role, MYC is currently considered “undruggable.” However, alternative therapeutic strategies have emerged, such as targeting upstream regulators like BRD4 using inhibitors such as JQ1, which has shown potential in suppressing MYC-driven transcriptional programs. Such approaches may hold particular promise for South Asian HNSCC patients harboring the MYC T73A mutation.

Copy number alterations (CNAs) represent a significant form of genomic instability in cancer, frequently impacting oncogenes and tumor suppressor genes. Among the observed copy number gains, amplification of the EGFR gene was identified. This finding contrasts with the Indian HNSCC-ICGC cohort, where EGFR amplification was not reported. One potential explanation is the higher proportion of HPV-positive patients in the ICGC cohort, whereas our South Asian cohort had negligible HPV-positive cases. Notably, EGFR amplification is predominantly reported in HPV-negative HNSCC patients. In fact, none of the CNVs reported in the Indian ICGC cohort were identified in our South Asian cohort [8]. The gain of chromosome region 11q13.3, which harbors the *CCND1* gene, was detected exclusively in cohort C. Since this cohort consists solely of tongue cancer cases, it would be valuable to investigate whether amplification of this gene is specific to tongue cancer and to further explore its functional role in this context. Regarding copy number losses, deletion of 1p36.32, which includes the *TP73* gene, was observed in two cohorts. TP73 is critical for regulating apoptosis, cell cycle arrest, and genomic stability. Unlike *TP53*, which is commonly mutated in cancers, *TP73* is more frequently inactivated via epigenetic silencing or genomic deletions, especially in solid tumors [46]. Loss of heterozygosity (LOH) at 1p36.3, encompassing the *TP73* locus, has been frequently reported in oral squamous cell carcinoma (OSCC) [47]. Significant deletions were also noted at 5q31.3 in cohorts A and B. This region contains a cluster of *PCDHB* genes, members of the cadherin superfamily involved in cell-cell adhesion, signaling, and tissue architecture maintenance. Loss of protocadherin genes has been associated with multiple cancers and is believed to facilitate increased cellular motility, reduced adhesion, and enhanced tumor invasiveness. Thus, the recurrent deletion of 5q31.3 and the involvement of multiple *PCDHB* genes suggest a potentially important role for this gene cluster in the pathogenesis of head and neck squamous cell carcinoma (HNSCC). Although we identified significant proportion of CNAs, it should be noted that since our analysis was based on exome sequencing data, it may not capture the full spectrum of CNVs and thus does not provide a comprehensive catalogue of copy number alterations.

Although rare, germline genetic variants can contribute to cancer susceptibility. To investigate this in the context of head and neck cancer (HNC), we examined putative functional germline variants across three South Asian cohorts (A, B, and C). Notably, the p.S456L variant in the *SDHA* gene was consistently identified across all three cohorts. This variant was also present in ∼15% of patients in the Indian ICGC-HNSCC cohort, encompassing both HPV-positive and HPV-negative cases, suggesting it may be a genetic risk factor for the HNSCC cancer among South Asian populations. SDHA encodes a core subunit of the succinate dehydrogenase (SDH) complex, which functions in both the tricarboxylic acid (TCA) cycle and the mitochondrial electron transport chain. While pathogenic mutations in SDHB, SDHC, and SDHD have been well documented in several cancer types, including paragangliomas and gastrointestinal stromal tumors, SDHA mutations are less commonly reported. To date, the p.S456L variant has only been previously documented in a single patient from a Middle Eastern population. Functional studies suggest that this variant destabilizes the SDHA protein, potentially impairing SDH complex activity [48]. However, in our study, all individuals harboring this variant carried it in a heterozygous state, and no loss of heterozygosity (LOH) was observed in matched tumor samples. This suggests that if the variant contributes to oncogenesis, it may act via a dominant-negative mechanism or require a secondary hit, consistent with the “two-hit” hypothesis in tumor suppressor gene inactivation. While functionally deleterious, the precise role of the p.S456L variant in cancer development remains to be fully elucidated. Besides this, we observed a trend that patients harboring p.S456L variants were diagnosed with cancer at younger age compared to patients carrying wild-type alleles. In our previous study, we reported a higher incidence rate of oral cancer among the younger population in the coastal region of Karnataka [49]. Since the samples in this study (Cohorts A and B) were collected from the same geographical area, the p.S456L variant might contribute to the increased cancer risk observed in this region. This finding opens an opportunity to investigate this variant in oral cancer patients from other geographical regions of South Asia. This is the expected epidemiological signature of a genetic variant that increases disease susceptibility. Additionally, two other germline variants—p.G532S in PCDHB3 and p.S693R in MEP1A—were identified and may also confer increased genetic risk. However, MEP1A expression is largely restricted to intestinal tissues (according to datasets available at https://vastdb.crg.eu and https://www.proteinatlas.org/), raising an unresolved question as to how a germline variant in this gene could mechanistically influence the risk of developing head and neck cancer. This remains an open area of investigation. On the other hand, PCDHB3 express in most of the tissues including tissues in the head and neck cancer.

In summary, this study has identified novel oncogenes distinct from those predominantly observed in Caucasian populations, likely reflecting differences in ethnic background and associated clinical parameters. These newly identified candidate drivers require validation in additional set of independent cohorts to confirm their prevalence and significance. Moreover, detailed functional studies are essential to elucidate their mechanistic roles in tumorigenesis. Evaluating the therapeutic potential of these targets in preclinical and clinical settings will be critical to advance personalized treatment approaches tailored to diverse patient populations.

## Data Availability

All data produced in the present study are available upon reasonable request to the authors

## Acknowledgement

We thank all the patients for providing samples, and all others who contributed to the sample collection process. The H.K. acknowledges the support received through the ICMR-DHR Women Scientist. The study was carried out with the collaboration of the Department of Oral Pathology, Yenepoya Dental College, Oncopathology, Surgical oncology, Yenepoya medical college, German cancer Research center, Germany and Uniklinik-Freiburg, Germany. Institutional support from Yenepoya University and Zulekha Yenepoya Institute of Oncology, is gratefully acknowledged.

## Author contribution

S.N. and M.S.M. conceptualized the work; R.A. provide resources, H.K. and R.A. acquire funding; H.K., R.T.M. and R.A. collected samples for the Cohort A; H.K. performed the experiments; S.N. supervised the study, performed the analysis and wrote paper with H.K.; Project was administered by R.A., S.N. and H.K.; M.K. was involved in visualization; all authors were involved in reviewing and editing

## Funding

This research has been supported by the ICMR-DHR, Government of India.

## Role of the funding source

The funder had no role in study design, data collection and analysis, decision to publish, or preparation of the manuscript.

## Competing Interests

The authors declare no competing financial interests

## Data availability

Raw data is available with corresponding authors and can be requested to access it.

## Supplementary figures

**Supplementary figure 1.** Lollipop plot of the remaining recurrently mutated genes with hotspot mutations.

**Supplementary figure 2.** Manhattan plots of copy number gains in selected samples. The left, middle and right subpanels in B are related to sample #24, #32 and #28, respectively (**Supplementary Table 1**).

**Supplementary figure 3.** Distribution of putative functional and common germline variants among patients in Cohorts A, B, and C. The’+’ symbol indicates the presence of a heterozygous genotype, the’.’ symbol denotes a homozygous wild-type genotype, and the’-’ symbol indicates insufficient read depth to confidently determine the genotype.

## Notes

### Competing Interest Statement

The authors have declared no competing interest.

### Author Declarations

Yenepoya ethics committee-1 kindly gave ethical approval for this work. The ID of the approval is: YEC-1/2023/131.

