## Supplementary table for "The somatic and germline mutational landscape of HPV-negative oral cancer patients with a history of chewing tobacco and betel nut use"

**Supplementary table 1**: Demographic details of the patients. Patient-wise sample details for Cohort D (TCGA) can be obtained from cBioPortal and from PMID: 25631445

| sample # | Cancer tissue ID | Normal tissue ID | Cohort | HPV | Age | Gender |
|---|---|---|---|---|---|---|
| 1 | 7736J | 7736N | Cohort-A | - | 76-81 | F |
| 2 | 8412E | 8412P | Cohort-A | - | 56-61 | M |
| 3 | 9465C | 9465K | Cohort-A | - | 51-56 | F |
| 4 | 9640E | 9640L | Cohort-A | - | 81-86 | M |
| 5 | 9675D | 9675B | Cohort-A | - | 46-51 | M |
| 6 | 9472C | 9472R | Cohort-A | - | 66-71 | M |
| 7 | 5286G | 5286Q | Cohort-A | - | 41-46 | F |
| 8 | NA | 9653K | Cohort-A | - | 71-76 | M |
| 9 | NA | 10175A | Cohort-A | - | 56-61 | F |
| 10 | NA | 10026C | Cohort-A | - | 41-46 | M |
| 11 | 6750C | 6750N | Cohort-A | - | 71-76 | M |
| 12 | 6702E | NA | Cohort-A | - | 71-76 | M |
| 13 | NA | 6627T | Cohort-A | - | 56-61 | F |
| 14 | 6331D | 6331A | Cohort-A | - | 51-56 | F |
| 15 | 5521F | 5521N | Cohort-A | - | 36-41 | M |
| 16 | 5288G | NA | Cohort-A | - | 71-76 | M |
| 17 | 5212E | 5212C | Cohort-A | - | 61-66 | M |
| 18 | 5103G | NA | Cohort-A | - | 51-56 | M |
| 19 | 5058F | NA | Cohort-A | - | 46-51 | F |
| 20 | 4975M | 4975A | Cohort-A | - | 51-56 | F |
| 21 | 4968E | 4968U | Cohort-A | - | 46-51 | M |
| 22 | 4384A | NA | Cohort-A | - | 41-46 | M |
| 23 | 4741E | 4741H | Cohort-A | - | 41-46 | M |
| 24 | 41207403440 | 11NFBYEN/41207403451 | Cohort-A | - | 36-41 | M |
| 25 | 41207403442 | 12NFBYEN/41207403452 | Cohort-A | - | 36-41 | M |
| 26 | 41207403444 | 14NFBYEN/41207403454 | Cohort-A | - | 66-71 | F |
| 27 | 41207403445 | 15NFBYEN/41207403455 | Cohort-A | - | 56-61 | M |
| 28 | 41207403446 | 16NFBYEN/41207403456 | Cohort-A | - | 56-61 | M |
| 29 | 41207403447 | 17NFBYEN/41207403457 | Cohort-A | - | 61-66 | F |
| 30 | 41207403448 | 18NFBYEN/41207403458 | Cohort-A | - | 66-71 | F |
| 31 | 41207403449 | 19NFBYEN/41207403459 | Cohort-A | - | 56-61 | M |
| 32 | 41207403450 | 20NFBYEN/41207403460 | Cohort-A | - | 41-46 | F |
| 33 | 41207403461 | 22NFBYEN/41207403462 | Cohort-A | - | 61-66 | F |
| 34 | SRR14886042 | NA | Cohort-B | - | 71-76 | M |
| 35 | SRR14886043 | NA | Cohort-B | - | 36-41 | M |
| 36 | SRR14886044 | NA | Cohort-B | - | 46-51 | M |
| 37 | SRR14886045 | NA | Cohort-B | HPV-16; HPV-71 | <NA> | NA |
| 38 | SRR14886048 | NA | Cohort-B | - | <NA> | NA |
| 39 | SRR14886049 | NA | Cohort-B | HPV-16 | 41-46 | F |
| 40 | SRR14886050 | NA | Cohort-B | - | 56-61 | M |
| 41 | SRR14886051 | NA | Cohort-B | - | 56-61 | M |
| 42 | SRR14886055 | NA | Cohort-B | - | 46-51 | F |
| 43 | SRR14886059 | NA | Cohort-B | - | 36-41 | M |
| 44 | SRR14886060 | NA | Cohort-B | - | 46-51 | M |
| 45 | SRR14886061 | NA | Cohort-B | HPV-16 | 46-51 | M |
| 46 | SRR14886064 | NA | Cohort-B | - | 46-51 | F |
| 47 | SRR14886066 | NA | Cohort-B | - | 81-86 | M |
| 48 | SRR14886069 | NA | Cohort-B | - | 61-66 | F |
| 49 | SRR14886070 | NA | Cohort-B | - | 66-71 | M |
| 50 | SRR14886071 | NA | Cohort-B | - | 56-61 | F |
| 51 | SRR14886072 | NA | Cohort-B | - | 56-61 | F |
| 52 | SRR14886073 | NA | Cohort-B | - | 46-51 | M |
| 53 | SRR14886074 | NA | Cohort-B | - | 46-51 | M |
| 54 | SRR14886079 | NA | Cohort-B | - | 56-61 | F |
| 55 | SRR14886087 | NA | Cohort-B | - | 51-56 | M |
| 56 | SRR14886040 | NA | Cohort-B | - | 56-61 | M |
| 57 | SRR14886041 | NA | Cohort-B | - | 56-61 | F |
| 58 | SRR14886046 | NA | Cohort-B | - | 71-76 | M |
| 59 | SRR14886047 | NA | Cohort-B | - | 51-56 | F |
| 60 | SRR14886052 | NA | Cohort-B | - | <NA> | NA |
| 61 | SRR14886053 | NA | Cohort-B | - | 46-51 | M |
| 62 | SRR14886054 | NA | Cohort-B | - | <NA> | NA |
| 63 | SRR14886056 | NA | Cohort-B | - | 56-61 | M |
| 64 | SRR14886057 | NA | Cohort-B | - | 76-81 | M |
| 65 | SRR14886058 | NA | Cohort-B | HPV-21 | 36-41 | M |
| 66 | SRR14886062 | NA | Cohort-B | - | 46-51 | F |
| 67 | SRR14886063 | NA | Cohort-B | - | 76-81 | F |
| 68 | SRR14886065 | NA | Cohort-B | - | 41-46 | M |
| 69 | SRR14886067 | NA | Cohort-B | - | 46-51 | F |
| 70 | SRR14886068 | NA | Cohort-B | - | 26-31 | M |
| 71 | SRR14886075 | NA | Cohort-B | - | 51-56 | M |
| 72 | SRR14886076 | NA | Cohort-B | - | 36-41 | M |
| 73 | SRR14886077 | NA | Cohort-B | - | 66-71 | M |
| 74 | SRR14886078 | NA | Cohort-B | - | 46-51 | M |
| 75 | SRR14886080 | NA | Cohort-B | - | 51-56 | M |
| 76 | SRR14886081 | NA | Cohort-B | - | 56-61 | F |
| 77 | SRR14886082 | NA | Cohort-B | - | 56-61 | M |
| 78 | SRR14886083 | NA | Cohort-B | - | <NA> | NA |
| 79 | SRR14886084 | NA | Cohort-B | - | 56-61 | F |
| 80 | SRR14886085 | NA | Cohort-B | - | 41-46 | M |
| 81 | SRR14886086 | NA | Cohort-B | - | 51-56 | F |
| 82 | SRR14886088 | NA | Cohort-B | - | 46-51 | M |
| 83 | SRR14886089 | NA | Cohort-B | - | 41-46 | M |
| 84 | SRR14886090 | NA | Cohort-B | - | <NA> | NA |

| | | | | | | |
|---|---|---|---|---|---|---|
| 85 | ERR1429982 | NA | Cohort-C | - | NA | NA |
| 86 | ERR1429983 | NA | Cohort-C | - | NA | NA |
| 87 | ERR1429984 | NA | Cohort-C | - | NA | NA |
| 88 | ERR1429985 | NA | Cohort-C | - | NA | NA |
| 89 | ERR1429986 | NA | Cohort-C | - | NA | NA |
| 90 | ERR1429987 | NA | Cohort-C | - | NA | NA |
| 91 | ERR1429988 | NA | Cohort-C | - | NA | NA |
| 92 | ERR1429989 | NA | Cohort-C | - | NA | NA |
| 93 | ERR1429990 | NA | Cohort-C | - | NA | NA |
| 94 | ERR1429991 | NA | Cohort-C | - | NA | NA |
| 95 | ERR1429992 | NA | Cohort-C | - | NA | NA |
| 96 | ERR1429993 | NA | Cohort-C | - | NA | NA |
| 97 | ERR1429994 | NA | Cohort-C | - | NA | NA |
| 98 | ERR1429995 | NA | Cohort-C | - | NA | NA |
| 99 | ERR1429996 | NA | Cohort-C | - | NA | NA |
| 100 | ERR1429997 | NA | Cohort-C | - | NA | NA |
| 101 | ERR1429998 | NA | Cohort-C | - | NA | NA |
| 102 | ERR1429999 | NA | Cohort-C | - | NA | NA |
| 103 | ERR1430000 | NA | Cohort-C | - | NA | NA |
| 104 | ERR1430001 | NA | Cohort-C | - | NA | NA |
| 105 | ERR1430002 | NA | Cohort-C | - | NA | NA |
| 106 | ERR1430003 | NA | Cohort-C | - | NA | NA |
| 107 | ERR1430004 | NA | Cohort-C | - | NA | NA |
| 108 | ERR1430005 | NA | Cohort-C | - | NA | NA |

**Supplementary table 2**: Tumor mutation burden among patients

| Cohort | Tumor_Sample_ Barcode | Total somatic mutations | Total somatic mutations per MB | Cohort | Tumor_Sample Barcode | Total somatic mutations | Total somatic mutations per MB |
|---|---|---|---|---|---|---|---|
| Cohort A | 41207403440 | 76 | 1.52 | Cohort B | SRR14886063 | 498 | 9.96 |
| Cohort A | 41207403442 | 44 | 0.88 | Cohort B | SRR14886064 | 32795 | 655.9 |
| Cohort A | 41207403444 | 122 | 2.44 | Cohort B | SRR14886065 | 1092 | 21.84 |
| Cohort A | 41207403445 | 57 | 1.14 | Cohort B | SRR14886066 | 535 | 10.7 |
| Cohort A | 41207403446 | 84 | 1.68 | Cohort B | SRR14886067 | 1553 | 31.06 |
| Cohort A | 41207403447 | 70 | 1.4 | Cohort B | SRR14886068 | 1050 | 21 |
| Cohort A | 41207403448 | 184 | 3.68 | Cohort B | SRR14886069 | 1566 | 31.32 |
| Cohort A | 41207403449 | 111 | 2.22 | Cohort B | SRR14886070 | 1660 | 33.2 |
| Cohort A | 41207403450 | 167 | 3.34 | Cohort B | SRR14886071 | 591 | 11.82 |
| Cohort A | 41207403461 | 494 | 9.88 | Cohort B | SRR14886072 | 1304 | 26.08 |
| Cohort A | 4384A | 3446 | 68.92 | Cohort B | SRR14886073 | 1892 | 37.84 |
| Cohort A | 4741E | 1197 | 23.94 | Cohort B | SRR14886074 | 1057 | 21.14 |
| Cohort A | 4968E | 1521 | 30.42 | Cohort B | SRR14886075 | 2825 | 56.5 |
| Cohort A | 4975M | 1243 | 24.86 | Cohort B | SRR14886076 | 633 | 12.66 |
| Cohort A | 5058F | 5356 | 107.12 | Cohort B | SRR14886077 | 254 | 5.08 |
| Cohort A | 5103G | 5617 | 112.34 | Cohort B | SRR14886078 | 597 | 11.94 |
| Cohort A | 5212E | 1825 | 36.5 | Cohort B | SRR14886079 | 3158 | 63.16 |
| Cohort A | 5286G | 1835 | 36.7 | Cohort B | SRR14886080 | 309 | 6.18 |
| Cohort A | 5288G | 4885 | 97.7 | Cohort B | SRR14886081 | 3984 | 79.68 |
| Cohort A | 5521F | 1356 | 27.12 | Cohort B | SRR14886082 | 771 | 15.42 |
| Cohort A | 6331D | 994 | 19.88 | Cohort B | SRR14886083 | 1802 | 36.04 |
| Cohort A | 6702E | 1923 | 38.46 | Cohort B | SRR14886084 | 2823 | 56.46 |
| Cohort A | 6750C | 1661 | 33.22 | Cohort B | SRR14886085 | 2453 | 49.06 |
| Cohort A | 7736J | 900 | 18 | Cohort B | SRR14886086 | 671 | 13.42 |
| Cohort A | 8412E | 1902 | 38.04 | Cohort B | SRR14886087 | 508 | 10.16 |
| Cohort A | 9465C | 2329 | 46.58 | Cohort B | SRR14886088 | 332 | 6.64 |
| Cohort A | 9472C | 1232 | 24.64 | Cohort B | SRR14886089 | 614 | 12.28 |
| Cohort A | 9640E | 501 | 10.02 | Cohort B | SRR14886090 | 795 | 15.9 |
| Cohort A | 9675D | 276 | 5.52 | Cohort C | ERR1429982 | 3247 | 64.94 |
| Cohort B | SRR14886040 | 1670 | 33.4 | Cohort C | ERR1429983 | 365 | 7.3 |
| Cohort B | SRR14886041 | 2127 | 42.54 | Cohort C | ERR1429984 | 350 | 7 |
| Cohort B | SRR14886042 | 16317 | 326.34 | Cohort C | ERR1429985 | 315 | 6.3 |
| Cohort B | SRR14886043 | 359 | 7.18 | Cohort C | ERR1429986 | 383 | 7.66 |
| Cohort B | SRR14886044 | 1456 | 29.12 | Cohort C | ERR1429987 | 397 | 7.94 |
| Cohort B | SRR14886045 | 391 | 7.82 | Cohort C | ERR1429988 | 310 | 6.2 |
| Cohort B | SRR14886046 | 240 | 4.8 | Cohort C | ERR1429989 | 410 | 8.2 |
| Cohort B | SRR14886047 | 221 | 4.42 | Cohort C | ERR1429990 | 339 | 6.78 |
| Cohort B | SRR14886048 | 539 | 10.78 | Cohort C | ERR1429991 | 272 | 5.44 |
| Cohort B | SRR14886049 | 1846 | 36.92 | Cohort C | ERR1429992 | 330 | 6.6 |
| Cohort B | SRR14886050 | 2372 | 47.44 | Cohort C | ERR1429993 | 912 | 18.24 |
| Cohort B | SRR14886051 | 1318 | 26.36 | Cohort C | ERR1429995 | 310 | 6.2 |
| Cohort B | SRR14886052 | 2399 | 47.98 | Cohort C | ERR1429996 | 233 | 4.66 |
| Cohort B | SRR14886053 | 1483 | 29.66 | Cohort C | ERR1429997 | 318 | 6.36 |
| Cohort B | SRR14886054 | 14163 | 283.26 | Cohort C | ERR1429998 | 330 | 6.6 |
| Cohort B | SRR14886055 | 2098 | 41.96 | Cohort C | ERR1429999 | 1134 | 22.68 |
| Cohort B | SRR14886056 | 27139 | 542.78 | Cohort C | ERR1430000 | 525 | 10.5 |
| Cohort B | SRR14886057 | 27513 | 550.26 | Cohort C | ERR1430001 | 4609 | 92.18 |
| Cohort B | SRR14886058 | 26898 | 537.96 | Cohort C | ERR1430002 | 3963 | 79.26 |
| Cohort B | SRR14886059 | 1430 | 28.6 | Cohort C | ERR1430003 | 341 | 6.82 |
| Cohort B | SRR14886060 | 5246 | 104.92 | Cohort C | ERR1430004 | 230 | 4.6 |
| Cohort B | SRR14886061 | 1841 | 36.82 | Cohort C | ERR1430005 | 229 | 4.58 |
| Cohort B | SRR14886062 | 2497 | 49.94 | | | | |

**Supplementary table 3**: Distribution of somatic transition and transversion mutations across samples

| Cohort | Samples | Proportion of mutation type | | | | | | Proportion of Ti and Tv | |
|---|---|---|---|---|---|---|---|---|---|
| | | C>A (%) | C>G (%) | C>T (%) | T>C (%) | T>A (%) | T>G (%) | Ti | Tv |
| Cohort A | 41207403440 | 10.00 | 1.82 | 70.91 | 12.73 | 1.82 | 2.73 | 83.64 | 16.36 |
| Cohort A | 41207403442 | 11.94 | 11.94 | 61.19 | 7.46 | 5.97 | 1.49 | 68.66 | 31.34 |
| Cohort A | 41207403444 | 10.06 | 8.28 | 59.76 | 11.83 | 7.69 | 2.37 | 71.60 | 28.40 |
| Cohort A | 41207403445 | 5.00 | 8.75 | 76.25 | 6.25 | 3.75 | 0.00 | 82.50 | 17.50 |
| Cohort A | 41207403446 | 4.65 | 5.43 | 77.52 | 7.75 | 3.10 | 1.55 | 85.27 | 14.73 |
| Cohort A | 41207403447 | 15.69 | 5.88 | 63.73 | 7.84 | 6.86 | 0.00 | 71.57 | 28.43 |
| Cohort A | 41207403448 | 9.89 | 12.09 | 60.07 | 9.89 | 5.86 | 2.20 | 69.96 | 30.04 |
| Cohort A | 41207403449 | 13.29 | 12.59 | 58.04 | 7.69 | 4.90 | 3.50 | 65.73 | 34.27 |
| Cohort A | 41207403450 | 8.05 | 18.22 | 61.02 | 6.78 | 3.39 | 2.54 | 67.80 | 32.20 |
| Cohort A | 41207403461 | 6.26 | 32.02 | 57.64 | 2.04 | 0.73 | 1.31 | 59.68 | 40.32 |
| Cohort A | 4384A | 4.26 | 1.74 | 86.54 | 4.57 | 2.05 | 0.85 | 91.11 | 8.89 |
| Cohort A | 4741E | 3.42 | 0.78 | 92.77 | 1.46 | 1.12 | 0.45 | 94.23 | 5.77 |
| Cohort A | 4968E | 2.11 | 1.14 | 90.81 | 3.47 | 1.49 | 0.97 | 94.29 | 5.71 |
| Cohort A | 4975M | 3.63 | 5.17 | 83.89 | 4.75 | 1.33 | 1.23 | 88.64 | 11.36 |
| Cohort A | 5058F | 4.93 | 1.60 | 86.09 | 4.25 | 2.22 | 0.91 | 90.33 | 9.67 |
| Cohort A | 5103G | 6.44 | 2.25 | 83.78 | 4.04 | 2.29 | 1.20 | 87.82 | 12.18 |
| Cohort A | 5212E | 4.40 | 1.11 | 87.08 | 4.12 | 2.18 | 1.11 | 91.20 | 8.80 |
| Cohort A | 5286G | 2.34 | 1.40 | 88.30 | 3.89 | 2.63 | 1.44 | 92.19 | 7.81 |
| Cohort A | 5288G | 5.58 | 2.48 | 83.23 | 5.04 | 2.52 | 1.14 | 88.28 | 11.72 |
| Cohort A | 5521F | 3.36 | 1.04 | 89.88 | 3.06 | 1.58 | 1.09 | 92.94 | 7.06 |
| Cohort A | 6331D | 3.29 | 0.92 | 92.04 | 2.56 | 0.79 | 0.39 | 94.61 | 5.39 |
| Cohort A | 6702E | 7.87 | 3.56 | 76.70 | 7.50 | 2.71 | 1.67 | 84.19 | 15.81 |
| Cohort A | 6750C | 5.27 | 2.47 | 84.57 | 4.06 | 2.43 | 1.21 | 88.62 | 11.38 |
| Cohort A | 7736J | 8.82 | 2.00 | 80.91 | 3.91 | 3.27 | 1.09 | 84.82 | 15.18 |
| Cohort A | 8412E | 4.69 | 3.05 | 85.65 | 3.29 | 2.19 | 1.13 | 88.93 | 11.07 |
| Cohort A | 9465C | 5.49 | 1.49 | 85.34 | 3.75 | 2.62 | 1.32 | 89.09 | 10.91 |
| Cohort A | 9472C | 5.48 | 2.86 | 82.64 | 5.31 | 2.00 | 1.71 | 87.95 | 12.05 |
| Cohort A | 9640E | 6.10 | 3.25 | 79.35 | 6.10 | 2.99 | 2.21 | 85.45 | 14.55 |
| Cohort A | 9675D | 3.32 | 3.32 | 82.86 | 6.91 | 2.81 | 0.77 | 89.77 | 10.23 |
| Cohort B | SRR14886040 | 32.69 | 3.84 | 26.09 | 30.78 | 2.30 | 4.31 | 56.86 | 43.14 |
| Cohort B | SRR14886041 | 18.98 | 6.62 | 48.94 | 18.37 | 3.48 | 3.60 | 67.31 | 32.69 |
| Cohort B | SRR14886042 | 43.73 | 5.99 | 39.84 | 7.07 | 2.77 | 0.58 | 46.92 | 53.08 |
| Cohort B | SRR14886043 | 10.24 | 13.19 | 50.00 | 18.75 | 2.60 | 5.21 | 68.75 | 31.25 |
| Cohort B | SRR14886044 | 26.10 | 6.43 | 50.75 | 11.28 | 2.95 | 2.49 | 62.03 | 37.97 |
| Cohort B | SRR14886045 | 7.87 | 8.35 | 58.11 | 15.75 | 5.98 | 3.94 | 73.86 | 26.14 |
| Cohort B | SRR14886046 | 9.60 | 11.30 | 48.87 | 20.90 | 4.80 | 4.52 | 69.77 | 30.23 |
| Cohort B | SRR14886047 | 9.90 | 9.64 | 48.98 | 20.81 | 4.82 | 5.84 | 69.80 | 30.20 |
| Cohort B | SRR14886048 | 10.18 | 9.88 | 41.14 | 23.32 | 6.62 | 8.86 | 64.46 | 35.54 |
| Cohort B | SRR14886049 | 6.39 | 5.67 | 65.82 | 13.16 | 3.89 | 5.07 | 78.98 | 21.02 |
| Cohort B | SRR14886050 | 7.33 | 6.47 | 68.94 | 10.22 | 2.68 | 4.37 | 79.15 | 20.85 |
| Cohort B | SRR14886051 | 4.10 | 7.33 | 71.73 | 10.47 | 3.32 | 3.05 | 82.20 | 17.80 |
| Cohort B | SRR14886052 | 5.41 | 5.74 | 69.24 | 11.13 | 4.27 | 4.21 | 80.37 | 19.63 |
| Cohort B | SRR14886053 | 5.30 | 5.69 | 67.58 | 12.93 | 3.69 | 4.82 | 80.51 | 19.49 |
| Cohort B | SRR14886054 | 6.86 | 3.10 | 79.16 | 5.08 | 4.01 | 1.79 | 84.24 | 15.76 |
| Cohort B | SRR14886055 | 23.87 | 5.14 | 58.74 | 8.45 | 1.97 | 1.83 | 67.20 | 32.80 |
| Cohort B | SRR14886056 | 28.03 | 3.59 | 59.31 | 6.36 | 2.17 | 0.53 | 65.67 | 34.33 |
| Cohort B | SRR14886057 | 33.44 | 5.82 | 48.87 | 8.93 | 2.29 | 0.65 | 57.80 | 42.20 |
| Cohort B | SRR14886058 | 42.26 | 5.50 | 40.25 | 8.55 | 2.78 | 0.66 | 48.80 | 51.20 |
| Cohort B | SRR14886059 | 8.07 | 6.82 | 67.86 | 9.72 | 3.66 | 3.88 | 77.57 | 22.43 |
| Cohort B | SRR14886060 | 8.47 | 7.53 | 55.35 | 13.30 | 7.53 | 7.82 | 68.65 | 31.35 |
| Cohort B | SRR14886061 | 4.72 | 4.44 | 74.39 | 8.53 | 4.02 | 3.91 | 82.91 | 17.09 |
| Cohort B | SRR14886062 | 12.61 | 8.60 | 43.99 | 14.74 | 11.14 | 8.92 | 58.73 | 41.27 |
| Cohort B | SRR14886063 | 9.29 | 10.64 | 45.13 | 21.84 | 4.93 | 8.17 | 66.97 | 33.03 |
| Cohort B | SRR14886064 | 34.26 | 4.81 | 51.11 | 6.95 | 2.34 | 0.53 | 58.06 | 41.94 |
| Cohort B | SRR14886065 | 11.70 | 11.25 | 48.11 | 14.17 | 7.28 | 7.48 | 62.29 | 37.71 |
| Cohort B | SRR14886066 | 10.82 | 9.08 | 56.72 | 15.30 | 3.73 | 4.35 | 72.01 | 27.99 |
| Cohort B | SRR14886067 | 12.24 | 10.15 | 44.24 | 18.18 | 7.89 | 7.30 | 62.42 | 37.58 |
| Cohort B | SRR14886068 | 10.05 | 9.56 | 49.20 | 17.83 | 6.62 | 6.74 | 67.03 | 32.97 |
| Cohort B | SRR14886069 | 12.54 | 9.95 | 40.32 | 19.21 | 8.43 | 9.55 | 59.53 | 40.47 |
| Cohort B | SRR14886070 | 9.89 | 9.58 | 46.43 | 16.86 | 9.80 | 7.46 | 63.28 | 36.72 |
| Cohort B | SRR14886071 | 7.88 | 10.24 | 48.41 | 19.55 | 6.65 | 7.27 | 67.96 | 32.04 |
| Cohort B | SRR14886072 | 17.38 | 5.13 | 59.25 | 11.89 | 3.10 | 3.25 | 71.14 | 28.86 |
| Cohort B | SRR14886073 | 11.76 | 10.59 | 42.42 | 17.03 | 9.74 | 8.46 | 59.45 | 40.55 |
| Cohort B | SRR14886074 | 29.88 | 5.87 | 50.38 | 10.01 | 2.00 | 1.86 | 60.39 | 39.61 |
| Cohort B | SRR14886075 | 8.07 | 7.27 | 59.30 | 12.62 | 6.81 | 5.94 | 71.92 | 28.08 |
| Cohort B | SRR14886076 | 8.70 | 26.69 | 49.04 | 10.11 | 2.93 | 2.53 | 59.15 | 40.85 |
| Cohort B | SRR14886077 | 8.86 | 10.23 | 51.82 | 19.09 | 3.86 | 6.14 | 70.91 | 29.09 |
| Cohort B | SRR14886078 | 9.53 | 9.71 | 50.27 | 18.78 | 5.63 | 6.08 | 69.06 | 30.94 |
| Cohort B | SRR14886079 | 13.92 | 11.02 | 34.72 | 18.27 | 11.84 | 10.24 | 52.99 | 47.01 |
| Cohort B | SRR14886080 | 9.33 | 10.82 | 52.80 | 18.47 | 4.66 | 3.92 | 71.27 | 28.73 |
| Cohort B | SRR14886081 | 9.88 | 7.26 | 56.23 | 11.59 | 8.30 | 6.73 | 67.82 | 32.18 |
| Cohort B | SRR14886082 | 7.24 | 8.22 | 61.03 | 13.59 | 3.91 | 6.02 | 74.61 | 25.39 |
| Cohort B | SRR14886083 | 7.64 | 6.99 | 63.94 | 11.32 | 4.66 | 5.46 | 75.25 | 24.75 |
| Cohort B | SRR14886084 | 12.56 | 9.81 | 51.09 | 12.08 | 8.18 | 6.28 | 63.17 | 36.83 |
| Cohort B | SRR14886085 | 7.84 | 2.82 | 83.05 | 4.40 | 0.92 | 0.98 | 87.45 | 12.55 |
| Cohort B | SRR14886086 | 8.35 | 12.66 | 50.73 | 17.43 | 6.61 | 4.22 | 68.17 | 31.83 |
| Cohort B | SRR14886087 | 9.99 | 9.32 | 49.61 | 17.40 | 6.85 | 6.85 | 67.00 | 33.00 |
| Cohort B | SRR14886088 | 10.83 | 9.36 | 49.36 | 16.51 | 6.79 | 7.16 | 65.87 | 34.13 |
| Cohort B | SRR14886089 | 12.11 | 9.45 | 50.82 | 16.22 | 4.93 | 6.47 | 67.04 | 32.96 |
| Cohort B | SRR14886090 | 8.75 | 7.57 | 55.36 | 17.67 | 4.57 | 6.07 | 73.03 | 26.97 |
| Cohort C | ERR1429982 | 29.19 | 8.02 | 37.12 | 12.09 | 8.65 | 4.93 | 49.21 | 50.79 |
| Cohort C | ERR1429983 | 26.35 | 8.78 | 40.71 | 15.88 | 5.07 | 3.21 | 56.59 | 43.41 |
| Cohort C | ERR1429984 | 26.39 | 6.10 | 38.96 | 18.49 | 5.75 | 4.31 | 57.45 | 42.55 |
| Cohort C | ERR1429985 | 19.44 | 8.53 | 44.44 | 20.04 | 3.57 | 3.97 | 64.48 | 35.52 |
| Cohort C | ERR1429986 | 16.87 | 14.11 | 43.37 | 17.38 | 4.99 | 3.27 | 60.76 | 39.24 |

| | | | | | | | | | |
|---|---|---|---|---|---|---|---|---|---|
| Cohort C | ERR1429987 | 28.55 | 7.41 | 37.50 | 16.67 | 6.17 | 3.70 | 54.17 | 45.83 |
| Cohort C | ERR1429988 | 13.96 | 7.65 | 47.42 | 19.50 | 5.35 | 6.12 | 66.92 | 33.08 |
| Cohort C | ERR1429989 | 19.28 | 8.09 | 45.72 | 18.35 | 4.82 | 3.73 | 64.07 | 35.93 |
| Cohort C | ERR1429990 | 19.82 | 9.46 | 40.89 | 20.71 | 5.00 | 4.11 | 61.61 | 38.39 |
| Cohort C | ERR1429991 | 14.73 | 11.21 | 46.37 | 17.80 | 6.81 | 3.08 | 64.18 | 35.82 |
| Cohort C | ERR1429992 | 12.28 | 8.90 | 48.58 | 20.11 | 5.69 | 4.45 | 68.68 | 31.32 |
| Cohort C | ERR1429993 | 26.29 | 9.43 | 34.06 | 13.71 | 11.35 | 5.15 | 47.77 | 52.23 |
| Cohort C | ERR1429995 | 21.56 | 7.98 | 42.32 | 19.56 | 4.19 | 4.39 | 61.88 | 38.12 |
| Cohort C | ERR1429996 | 15.27 | 9.61 | 37.19 | 26.85 | 5.67 | 5.42 | 64.04 | 35.96 |
| Cohort C | ERR1429997 | 13.51 | 10.28 | 46.57 | 20.77 | 4.84 | 4.03 | 67.34 | 32.66 |
| Cohort C | ERR1429998 | 16.07 | 10.12 | 43.06 | 19.64 | 6.35 | 4.76 | 62.70 | 37.30 |
| Cohort C | ERR1429999 | 33.33 | 8.49 | 32.95 | 15.37 | 6.80 | 3.06 | 48.32 | 51.68 |
| Cohort C | ERR1430000 | 15.54 | 8.64 | 41.80 | 19.00 | 9.84 | 5.18 | 60.79 | 39.21 |
| Cohort C | ERR1430001 | 18.71 | 11.27 | 36.93 | 21.34 | 5.04 | 6.71 | 58.27 | 41.73 |
| Cohort C | ERR1430002 | 12.73 | 11.72 | 41.21 | 23.43 | 5.25 | 5.66 | 64.65 | 35.35 |
| Cohort C | ERR1430003 | 13.07 | 10.50 | 46.34 | 21.78 | 4.55 | 3.76 | 68.12 | 31.88 |
| Cohort C | ERR1430004 | 12.99 | 11.03 | 50.25 | 16.91 | 4.17 | 4.66 | 67.16 | 32.84 |
| Cohort C | ERR1430005 | 10.32 | 12.53 | 48.65 | 19.90 | 4.18 | 4.42 | 68.55 | 31.45 |

**Supplementary table 4:** Recurrently mutated genes identified through MutSig2CV algorithm

| gene | p | q | codelen | nnei | nncd | nsil | nmis | nstp | nspl | nind | nnon | npat | nsite | pCV | pCL | pFN |
|---|---|---|---|---|---|---|---|---|---|---|---|---|---|---|---|---|
| *TP53* | 6.66E-16 | 1.26E-11 | 1890 | 1 | 1 | 11 | 54 | 14 | 3 | 3 | 74 | 61 | 51 | 1.75E-12 | 3.00E-05 | 1.00E-05 |
| *CDKN2A* | 1.68E-12 | 1.58E-08 | 1002 | 0 | 5 | 1 | 9 | 13 | 2 | 4 | 28 | 23 | 19 | 5.31E-09 | 1.03E-02 | 1.00E-05 |
| *RPL8* | 1.12E-08 | 7.04E-05 | 796 | 4 | 0 | 5 | 7 | 0 | 5 | 18 | 30 | 17 | 15 | 1.39E-06 | 1.00E-04 | 6.51E-01 |
| *GPI* | 8.31E-08 | 3.92E-04 | 1866 | 2 | 0 | 5 | 14 | 0 | 2 | 8 | 24 | 16 | 15 | 4.09E-04 | 1.10E-04 | 5.94E-03 |
| *GOLGA6L20/L9* | 1.74E-07 | 6.56E-04 | 2666 | 0 | 5 | 1 | 9 | 17 | 0 | 0 | 26 | 23 | 9 | 8.89E-04 | 1.00E-05 | 1 |
| *CASP8* | 2.47E-07 | 7.24E-04 | 1749 | 6 | 2 | 3 | 15 | 6 | 2 | 4 | 27 | 22 | 26 | 1.65E-07 | 5.58E-01 | 2.50E-02 |
| *MYOM1* | 2.69E-07 | 7.24E-04 | 5206 | 3 | 0 | 18 | 43 | 4 | 12 | 5 | 64 | 30 | 43 | 1.76E-04 | 2.00E-05 | 2.27E-01 |
| *USP17* | 8.68E-07 | 2.05E-03 | 9552 | 0 | 2 | 1 | 7 | 13 | 0 | 0 | 20 | 19 | 11 | 4.86E-03 | 1.00E-05 | 7.21E-02 |
| *SLPI* | 2.60E-06 | 5.45E-03 | 413 | 2 | 0 | 0 | 2 | 0 | 5 | 0 | 7 | 7 | 3 | 6.42E-04 | 1.88E-02 | 2.14E-04 |
| *TRIM48* | 4.47E-06 | 8.43E-03 | 696 | 4 | 0 | 3 | 27 | 0 | 2 | 1 | 30 | 24 | 9 | 2.78E-02 | 1.00E-05 | 1.00E-05 |
| *ALDOA* | 8.03E-06 | 1.38E-02 | 1127 | 1 | 1 | 0 | 9 | 0 | 0 | 7 | 16 | 12 | 11 | 1.08E-03 | 4.00E-04 | 5.01E-01 |
| *MDH2* | 9.34E-06 | 1.47E-02 | 1051 | 7 | 0 | 4 | 11 | 2 | 1 | 2 | 16 | 14 | 10 | 6.78E-03 | 4.30E-04 | 9.49E-02 |
| *CD5* | 1.12E-05 | 1.58E-02 | 1529 | 1 | 0 | 5 | 17 | 2 | 0 | 1 | 20 | 17 | 12 | 7.39E-02 | 1.00E-05 | 3.89E-01 |
| *TM9SF2* | 1.18E-05 | 1.58E-02 | 2056 | 1 | 0 | 4 | 9 | 1 | 4 | 2 | 16 | 11 | 13 | 7.45E-04 | 4.50E-03 | 5.28E-02 |
| *ALG10* | 1.25E-05 | 1.58E-02 | 1430 | 4 | 0 | 0 | 12 | 2 | 0 | 0 | 14 | 12 | 9 | 8.35E-02 | 1.00E-05 | 9.83E-01 |
| *RPL38* | 1.72E-05 | 2.03E-02 | 231 | 3 | 4 | 0 | 1 | 0 | 0 | 4 | 5 | 5 | 2 | 2.35E-02 | 2.80E-03 | 1.43E-03 |
| *HM13* | 1.83E-05 | 2.03E-02 | 1400 | 0 | 0 | 2 | 10 | 2 | 6 | 4 | 22 | 14 | 19 | 2.61E-05 | 3.50E-02 | 3.91E-01 |
| *DYNC1LI1* | 2.35E-05 | 2.46E-02 | 1622 | 0 | 2 | 2 | 37 | 1 | 0 | 1 | 39 | 26 | 14 | 1.64E-01 | 1.00E-05 | 1.00E-05 |
| *NUP85* | 2.48E-05 | 2.47E-02 | 2045 | 1 | 0 | 6 | 16 | 1 | 3 | 2 | 22 | 17 | 15 | 1.58E-02 | 9.00E-05 | 1.17E-01 |
| *NOTCH1* | 2.93E-05 | 2.76E-02 | 7800 | 0 | 4 | 38 | 73 | 16 | 4 | 12 | 105 | 47 | 104 | 2.08E-06 | 7.88E-01 | 7.56E-01 |
| *FXYD3* | 3.54E-05 | 3.18E-02 | 636 | 0 | 6 | 1 | 3 | 0 | 0 | 8 | 11 | 9 | 5 | 2.55E-01 | 7.90E-04 | 8.30E-04 |
| *FAM160B1* | 3.98E-05 | 3.41E-02 | 2389 | 1 | 0 | 0 | 17 | 2 | 2 | 1 | 22 | 17 | 22 | 2.89E-06 | 1 | 5.77E-01 |
| *C5orf39* | 4.17E-05 | 3.42E-02 | 582 | 10 | 0 | 1 | 8 | 0 | 0 | 0 | 8 | 8 | 4 | 8.12E-03 | 2.25E-04 | 4.45E-01 |
| *SAT1* | 4.82E-05 | 3.79E-02 | 536 | 0 | 4 | 5 | 2 | 0 | 1 | 9 | 12 | 8 | 7 | 5.93E-02 | 1.70E-04 | 3.12E-02 |
| *LEO1* | 5.13E-05 | 3.81E-02 | 2047 | 16 | 0 | 9 | 10 | 3 | 8 | 2 | 23 | 19 | 16 | 1.15E-03 | 1.90E-03 | 1.31E-01 |
| *KRT14* | 5.26E-05 | 3.81E-02 | 1449 | 3 | 1 | 5 | 16 | 1 | 4 | 7 | 28 | 17 | 23 | 4.04E-03 | 5.00E-04 | 8.25E-01 |
| *NDFIP1* | 5.80E-05 | 4.05E-02 | 694 | 1 | 0 | 4 | 1 | 0 | 1 | 2 | 4 | 4 | 4 | 4.34E-02 | 1.00E-04 | 1.07E-02 |
| *SAMD9* | 6.28E-05 | 4.23E-02 | 4774 | 5 | 0 | 6 | 32 | 0 | 0 | 0 | 32 | 20 | 26 | 4.74E-01 | 1.00E-05 | 7.91E-01 |
| *FZD3* | 6.59E-05 | 4.29E-02 | 2021 | 7 | 0 | 4 | 16 | 3 | 1 | 3 | 23 | 16 | 23 | 4.99E-06 | 1 | 9.76E-01 |
| *KCNK9* | 7.47E-05 | 4.70E-02 | 1133 | 2 | 0 | 6 | 17 | 0 | 1 | 1 | 19 | 13 | 16 | 2.20E-02 | 1.00E-04 | 4.41E-01 |
| *SVIL* | 8.05E-05 | 4.86E-02 | 6785 | 7 | 5 | 24 | 56 | 8 | 5 | 2 | 71 | 35 | 67 | 1.35E-03 | 7.50E-03 | 4.76E-01 |
| *HLX* | 8.83E-05 | 4.86E-02 | 1479 | 5 | 0 | 7 | 32 | 0 | 0 | 2 | 34 | 24 | 23 | 2.08E-03 | 1.60E-03 | 9.96E-01 |
| *TSPAN8* | 8.86E-05 | 4.86E-02 | 742 | 0 | 0 | 2 | 2 | 0 | 2 | 6 | 10 | 8 | 7 | 1.72E-02 | 6.65E-03 | 5.33E-03 |
| *PCNX* | 8.97E-05 | 4.86E-02 | 7166 | 5 | 1 | 12 | 50 | 6 | 1 | 4 | 61 | 23 | 58 | 6.97E-01 | 1.00E-05 | 6.21E-01 |
| *POLG* | 9.08E-05 | 4.86E-02 | 3810 | 6 | 1 | 16 | 30 | 2 | 4 | 5 | 41 | 23 | 38 | 2.12E-02 | 1.78E-04 | 9.82E-01 |
| *CYHR1/ZFTRAF1* | 9.80E-05 | 4.86E-02 | 1492 | 0 | 1 | 9 | 6 | 1 | 0 | 0 | 7 | 7 | 7 | 7.67E-01 | 1 | 1.00E-05 |
| *KRT5* | 1.01E-04 | 4.86E-02 | 1805 | 8 | 0 | 7 | 15 | 0 | 0 | 9 | 24 | 19 | 17 | 1.08E-02 | 2.29E-04 | 5.90E-01 |
| *POU2F1* | 1.03E-04 | 4.86E-02 | 2292 | 8 | 5 | 8 | 17 | 3 | 0 | 4 | 24 | 17 | 21 | 6.22E-02 | 7.00E-05 | 3.41E-01 |
| *HNRNPA2B1* | 1.04E-04 | 4.86E-02 | 1126 | 0 | 0 | 1 | 7 | 0 | 0 | 5 | 12 | 10 | 9 | 1.74E-03 | 5.50E-03 | 1.17E-01 |
| *MPZL2* | 1.04E-04 | 4.86E-02 | 668 | 0 | 0 | 3 | 7 | 0 | 0 | 0 | 7 | 7 | 4 | 8.16E-01 | 1.00E-05 | 5.36E-01 |
| *MUC12* | 1.06E-04 | 4.86E-02 | 16052 | 0 | 7 | 41 | 109 | 4 | 2 | 4 | 119 | 52 | 101 | 8.33E-01 | 1.00E-05 | 9.65E-01 |

| | |
|---|---|
| rank | Position of the gene as sorted ascending by p-/q-value. |
| gene | HUGO symbol of the gene (RefSeq hg19) |
| longname | HUGO description of the gene |
| codelen | ORF length of the gene |
| nnei | Number of neighboring genes in the bagel used to estimate BMR |
| nncd | Number of noncoding mutations |
| nsil | Number of silent (synonymous) mutations in the gene |
| nmis | Number of missense mutations in the gene |
| nstp | Number of nonsense mutations in the gene |
| nspl | Number of splice site mutations in the gene (defined as +/- 2 bases from the donor/acceptor site) |
| nind | Number of insertions or deletions in the gene |
| nnon | Number of nonsilent mutations in the gene (including all indels and splice site mutations, even if the codon change is synonymous in the latter case) |
| npat | Number of patients with mutations in the gene |
| nsite | Number of uniquely mutated sites in the gene (does not multiply count recurrently mutated positions) |
| pCV | Abundance p-value |
| pCL | Clustering p-value |
| pFN | Functional (conservation) p-value |
| p | Overall p-value obtained from Fisher combination of pCV, pCL, and pFN |

**Supplementary table 5**: Recurrently mutated genes identified through dNdScv algorithm

| Gene name | pglobal cv | qglobal cv | n syn | n mis | n non | n spl | n ind | wmis cv | wnon cv | wspl cv | wind cv | pmis cv | ptrunc cv | pallsubs cv | pind cv | qmis cv | qtrunc cv | qallsubs cv | qind cv |
|---|---|---|---|---|---|---|---|---|---|---|---|---|---|---|---|---|---|---|---|
| *TP53* | 0 | 0 | 4 | 55 | 14 | 2 | 4 | 8.799583 | 23.81503 | 23.81503 | 3.376968 | 2.21E-13 | 2.86E-13 | 0 | 0.047261 | 4.36E-09 | 1.44E-09 | 0 | 1 |
| *GOLGA6L20/GOLGA6L9* | 0 | 0 | 1 | 6 | 14 | 0 | 1 | 11.65127 | 210.5518 | 210.5518 | 4.862131 | 0.000136 | 0 | 0 | 0.18289 | 0.15856 | 0 | 0 | 1 |
| *USP17L22* | 0 | 0 | 0 | 3 | 13 | 0 | 0 | 6.085276 | 358.9284 | 358.9284 | 0 | 0.029618 | 0 | 0 | 1 | 0.704028 | 0 | 0 | 1 |
| *CDKN2A.p16INK4a* | 0 | 0 | 0 | 6 | 13 | 2 | 4 | 3.600103 | 174.34 | 174.34 | 7.427996 | 0.040585 | 0 | 0 | 0.004276 | 0.704028 | 0 | 0 | 1 |
| *C1orf134* | 1.44E-11 | 5.77E-08 | 3 | 16 | 0 | 0 | 1 | 24.40213 | 0 | 0 | 5.395293 | 4.34E-13 | 0.64107 | 2.93E-12 | 0.166685 | 4.36E-09 | 0.823151 | 1.18E-08 | 1 |
| *TRIM48* | 2.08E-09 | 6.96E-06 | 4 | 23 | 0 | 2 | 1 | 11.57933 | 5.863518 | 5.863518 | 2.536652 | 4.55E-11 | 0.062734 | 2.71E-10 | 0.31679 | 3.05E-07 | 0.823151 | 9.08E-07 | 1 |
| *PRAMEF20* | 3.13E-08 | 8.99E-05 | 1 | 15 | 0 | 0 | 0 | 19.36476 | 0 | 0 | 0 | 2.12E-10 | 0.752232 | 1.47E-09 | 1 | 1.06E-06 | 0.835786 | 4.21E-06 | 1 |
| *CASP8* | 4.29E-08 | 0.000108 | 0 | 14 | 7 | 1 | 4 | 5.844285 | 28.50361 | 28.50361 | 3.461473 | 0.000327 | 5.41E-08 | 4.62E-08 | 0.044193 | 0.262483 | 0.000181 | 0.000103 | 1 |
| *NOTCH1* | 5.91E-08 | 0.000132 | 13 | 33 | 12 | 3 | 16 | 1.491832 | 9.397765 | 9.397765 | 2.193772 | 0.168683 | 1.10E-08 | 7.06E-08 | 0.040499 | 0.786251 | 4.44E-05 | 0.000142 | 1 |
| *PRAMEF17* | 1.90E-07 | 0.000381 | 2 | 10 | 0 | 0 | 3 | 13.64922 | 0 | 0 | 12.07509 | 5.58E-07 | 0.728501 | 3.16E-06 | 0.003095 | 0.001868 | 0.82973 | 0.004889 | 1 |
| *PRAMEF15* | 2.09E-07 | 0.000381 | 5 | 12 | 0 | 0 | 2 | 12.21359 | 0 | 0 | 8.360981 | 6.63E-08 | 0.69175 | 3.94E-07 | 0.027328 | 0.000266 | 0.823931 | 0.00072 | 1 |
| *LEO1* | 3.20E-07 | 0.000536 | 9 | 2 | 2 | 8 | 2 | 0.175777 | 7.359507 | 7.359507 | 1.108719 | 0.005484 | 1.82E-05 | 3.36E-08 | 0.504986 | 0.704028 | 0.036478 | 8.43E-05 | 1 |
| *CDKN2A.p14arf* | 4.50E-07 | 0.000695 | 2 | 17 | 0 | 0 | 4 | 7.545161 | 0 | 0 | 8.757182 | 2.35E-06 | 0.595697 | 9.90E-06 | 0.00245 | 0.004758 | 0.823151 | 0.01047 | 1 |
| *RPL8* | 5.48E-07 | 0.000786 | 2 | 3 | 0 | 5 | 7 | 0.938857 | 24.58415 | 24.58415 | 6.390529 | 0.92771 | 9.52E-06 | 3.41E-05 | 0.000876 | 0.989603 | 0.021251 | 0.029097 | 0.761658 |
| *RP11-166B2.1* | 8.64E-06 | 0.011094 | 10 | 14 | 0 | 0 | 5 | 3.865312 | 0 | 0 | 10.56163 | 0.000519 | 0.396065 | 0.001372 | 0.000409 | 0.327187 | 0.823151 | 0.56247 | 0.429327 |
| *SNRNP70* | 8.84E-06 | 0.011094 | 3 | 8 | 1 | 0 | 13 | 1.285879 | 1.954576 | 1.954576 | 10.11095 | 0.620845 | 0.563088 | 0.784225 | 7.33E-07 | 0.938015 | 0.823151 | 0.915893 | 0.014728 |
| *FAT1* | 9.63E-06 | 0.011384 | 24 | 29 | 13 | 0 | 17 | 0.607956 | 4.137514 | 4.137514 | 1.584572 | 0.060693 | 0.000147 | 4.29E-06 | 0.147074 | 0.713799 | 0.210783 | 0.006152 | 1 |
| *SLPI* | 1.02E-05 | 0.011428 | 0 | 0 | 0 | 5 | 0 | 0 | 48.89037 | 48.89037 | 0 | 0.206619 | 5.38E-07 | 6.73E-07 | 1 | 0.804974 | 0.001544 | 0.001127 | 1 |
| *AL161915.1* | 1.09E-05 | 0.011481 | 4 | 8 | 0 | 0 | 3 | 8.388163 | 0 | 0 | 14.06495 | 7.63E-05 | 0.702459 | 0.000352 | 0.002035 | 0.104424 | 0.824532 | 0.23584 | 0.99074 |
| *NKG2-E* | 1.28E-05 | 0.012862 | 3 | 10 | 0 | 0 | 4 | 6.233468 | 0 | 0 | 10.6517 | 0.000212 | 0.510983 | 0.00069 | 0.00124 | 0.202451 | 0.823151 | 0.357552 | 0.76556 |
| *HM13* | 1.34E-05 | 0.012862 | 0 | 4 | 1 | 5 | 4 | 1.701392 | 25.14558 | 25.14558 | 3.097323 | 0.444372 | 2.75E-06 | 1.52E-05 | 0.059469 | 0.897906 | 0.006904 | 0.015221 | 1 |
| *KRT18* | 1.58E-05 | 0.014404 | 17 | 0 | 0 | 2 | 4 | 0 | 1.652196 | 1.652196 | 2.33415 | 3.00E-06 | 0.525902 | 8.99E-06 | 0.119025 | 0.005482 | 0.823151 | 0.010357 | 1 |
| *PRAMEF19* | 1.91E-05 | 0.016704 | 2 | 5 | 1 | 0 | 4 | 6.240832 | 15.45973 | 15.45973 | 16.44622 | 0.005052 | 0.062174 | 0.005082 | 0.000259 | 0.704028 | 0.823151 | 0.915893 | 0.328949 |
| *KM-PA-2* | 2.74E-05 | 0.02198 | 4 | 12 | 0 | 1 | 2 | 7.661528 | 6.878319 | 6.878319 | 5.296514 | 9.03E-06 | 0.150651 | 3.19E-05 | 0.060628 | 0.013962 | 0.823151 | 0.029097 | 1 |
| *LGALS3* | 3.14E-05 | 0.024272 | 2 | 4 | 0 | 0 | 9 | 1.575382 | 0 | 0 | 11.9624 | 0.484076 | 0.510479 | 0.608009 | 3.69E-06 | 0.908559 | 0.823151 | 0.915893 | 0.037048 |
| *RP11-863K10.7* | 3.87E-05 | 0.028814 | 0 | 13 | 0 | 0 | 1 | 11.10854 | 0 | 0 | 1.767014 | 1.50E-06 | 0.642669 | 6.74E-06 | 0.416728 | 0.004305 | 0.823151 | 0.00903 | 1 |
| LPAR4 | 6.51E-05 | 0.046719 | 1 | 19 | 0 | 0 | 1 | 7.741262 | 0 | 0 | 1.094927 | 1.96E-06 | 0.638234 | 8.63E-06 | 0.570872 | 0.004758 | 0.823151 | 0.010357 | 1 |

gene_name Gene symbol being tested.
n_syn Number of observed synonymous mutations.
n_mis Number of observed missense mutations.
n_non Number of observed nonsense (stop-gain) mutations.
n_spl Number of observed splice-site mutations.
n_ind Number of observed indels.
wmis_cv Estimated dN/dS ratio for missense mutations (covariate-corrected)
wnon_cv Estimated dN/dS ratio for nonsense mutations.
wspl_cv Estimated dN/dS ratio for splice-site mutations.
wind_cv Estimated dN/dS ratio for indels.
pmis_cv P-value for enrichment of missense mutations.
ptrunc_cv P-value for enrichment of truncating mutations (nonsense + splice + indel)
pallsubs_cv P-value for enrichment across all substitutions (missense + nonsense + splice)
pind_cv P-value for enrichment of indels.
pglobal_cv Global p-value for enrichment across all mutation types.
qmis_cv FDR-adjusted (q-value) for missense enrichment.
qtrunc_cv FDR-adjusted q-value for truncating enrichment.
qallsubs_cv FDR-adjusted q-value for all substitutions.
qind_cv FDR-adjusted q-value for indel enrichment.
qglobal_cv Global FDR-adjusted q-value across all mutation types.

**Supplementary table 6:** Pathways enriched among significantly mutated genes

| Wiki-pathways | | | | | | | |
|---|---|---|---|---|---|---|---|
| **#term ID** | **term description** | **observed gene count** | **background gene count** | **strength** | **signal** | **false discovery rate** | **matching proteins in your network (labels)** |
| WP4674 | Head and neck squamous cell carcinoma | 5 | 72 | 1.42 | 0.89 | 0.0014 | *TP53,CASP8,FAT1, CDKN2A,NOTCH1* |
| **Disease gene association** | | | | | | | |
| **#term ID** | **term description** | **observed gene count** | **background gene count** | **strength** | **signal** | **false discovery rate** | **matching proteins in your network (labels)** |
| DOID:0050687 | Cell type cancer | 10 | 451 | 0.92 | 0.83 | 0.00026 | *KRT14,FZD3,KRT5,TP53,CD5, CASP8,KRT18,FAT1,CDKN2A,NOTCH1* |
| DOID:305 | Carcinoma | 9 | 307 | 1.05 | 0.91 | 0.00026 | *KRT14,FZD3,KRT5,TP53,CD5,KRT18, FAT1,CDKN2A,NOTCH1* |
| DOID:5520 | Head and neck squamous cell carcinoma | 4 | 11 | 2.14 | 1.23 | 0.00026 | *TP53,FAT1,CDKN2A,NOTCH1* |
| DOID:1749 | Squamous cell carcinoma | 5 | 51 | 1.57 | 1.13 | 0.00027 | *KRT5,TP53,FAT1,CDKN2A,NOTCH1* |
| DOID:0060058 | Lymphoma | 6 | 105 | 1.34 | 1.04 | 0.0003 | *KRT14,KRT5,TP53,CD5,CASP8,NOTCH1* |
| DOID:0060060 | non-Hodgkin lymphoma | 5 | 61 | 1.49 | 1.04 | 0.00046 | *KRT14,KRT5,TP53,CD5,NOTCH1* |
| DOID:1612 | Breast cancer | 5 | 62 | 1.49 | 1.04 | 0.00046 | *KRT5,TP53,CD5,CASP8,CDKN2A* |
| DOID:2513 | Basal cell carcinoma | 4 | 27 | 1.75 | 1.1 | 0.00046 | *KRT14,KRT5,TP53,CDKN2A* |
| DOID:2531 | Hematologic cancer | 7 | 190 | 1.14 | 0.91 | 0.00046 | *KRT14,KRT5,TP53,CD5,CASP8,CDKN2A,NOTCH1* |
| DOID:4159 | Skin cancer | 5 | 63 | 1.48 | 1.04 | 0.00046 | *KRT14,KRT5,TP53,CASP8,CDKN2A* |
| DOID:8557 | Oropharynx cancer | 3 | 7 | 2.21 | 1.13 | 0.00055 | *KRT5,TP53,CDKN2A* |
| DOID:0050686 | Organ system cancer | 11 | 757 | 0.74 | 0.64 | 0.00097 | *KRT14,FZD3,KRT5,TP53,LEO1,CD5,CASP8,* |
| DOID:74 | Hematopoietic system disease | 9 | 473 | 0.86 | 0.7 | 0.00097 | *KRT14,KRT5,TP53,CD5,CASP8,GPI,CDKN2A, ALDOA,NOTCH1* |
| DOID:1240 | Leukemia | 5 | 104 | 1.26 | 0.82 | 0.0018 | *TP53,CD5,CASP8,CDKN2A,NOTCH1* |
| DOID:0050743 | Mature T-cell and NK-cell lymphoma | 3 | 15 | 1.88 | 0.89 | 0.0024 | *KRT14,KRT5,CD5* |
| DOID:14566 | Disease of cellular proliferation | 12 | 1101 | 0.62 | 0.49 | 0.0042 | *KRT14,FZD3,KRT5,LGALS3,TP53,LEO1,CD5, CASP8,KRT18,FAT1,CDKN2A,NOTCH1* |
| DOID:0060735 | Epidermolysis bullosa simplex Dowling-Meara type | 2 | 2 | 2.58 | 0.77 | 0.0067 | *KRT14,KRT5* |
| DOID:0111346 | Epidermolysis bullosa simplex with mottled pigmentation | 2 | 2 | 2.58 | 0.77 | 0.0067 | *KRT14,KRT5* |
| DOID:11166 | Obsolete papillomavirus infectious disease | 2 | 2 | 2.58 | 0.77 | 0.0067 | *TP53,CDKN2A* |
| DOID:7039 | Borst-Jadassohn intraepidermal carcinoma | 2 | 3 | 2.4 | 0.71 | 0.0097 | *KRT14,KRT5* |
| DOID:3459 | Breast carcinoma | 3 | 29 | 1.59 | 0.65 | 0.0113 | *KRT5,TP53,CDKN2A* |
| DOID:11054 | Urinary bladder cancer | 3 | 38 | 1.48 | 0.54 | 0.0231 | *TP53,FAT1,CDKN2A* |
| DOID:3012 | Li-Fraumeni syndrome | 2 | 6 | 2.1 | 0.56 | 0.0247 | *TP53,CDKN2A* |
| DOID:3498 | Pancreatic ductal adenocarcinoma | 2 | 6 | 2.1 | 0.56 | 0.0247 | *TP53,CDKN2A* |
| DOID:6498 | Seborrheic keratosis | 2 | 6 | 2.1 | 0.56 | 0.0247 | *KRT14,KRT5* |
| DOID:8691 | Mycosis fungoides | 2 | 6 | 2.1 | 0.56 | 0.0247 | *KRT14,KRT5* |
| DOID:1909 | Melanoma | 3 | 46 | 1.39 | 0.49 | 0.0334 | *TP53,CASP8,CDKN2A* |
| DOID:2893 | Cervix carcinoma | 2 | 9 | 1.93 | 0.48 | 0.0413 | *TP53,CDKN2A* |
| DOID:9561 | Nasopharyngeal disease | 2 | 9 | 1.93 | 0.48 | 0.0413 | *TP53,CDKN2A* |
| DOID:0060085 | Organ system benign neoplasm | 5 | 237 | 0.9 | 0.4 | 0.0421 | *KRT14,KRT5,LGALS3,TP53,CDKN2A* |
| DOID:0060108 | Brain glioma | 2 | 10 | 1.88 | 0.48 | 0.0421 | *TP53,CDKN2A* |
| DOID:1037 | Lymphoid leukemia | 3 | 53 | 1.33 | 0.45 | 0.0421 | *CD5,CDKN2A,NOTCH1* |
| DOID:3347 | Osteosarcoma | 2 | 10 | 1.88 | 0.48 | 0.0421 | *TP53,LEO1* |

**Supplementary table 7:** *De-novo* hotspot mutations identified in this study

| Hugo_Symbol | AA | log10 (pvalue) | C | R-AA | TMG | MAFR | SNP_ID | V-AA | CC | Genomic_Position | Mutability | Mu protein | qvalue | Is_repeat | Entropy | | | TP |
|---|---|---|---|---|---|---|---|---|---|---|---|---|---|---|---|---|---|---|
| | | | | | | | | | | | | | | | pad12 | pad24 | pad36 | |
| *DYNC1LI1* | 448 | -40.58 | 20 | F:20 | 45 | 0.44 | novel:20 | A:20 | TTc/GCc:20 | 3:32570057_20 | NA | 0.04 | 9.46E-38 | FALSE | 1.32 | 1.27 | 1.36 | FALSE |
| *GOLGA6L9* | 289 | -36.14 | 16 | Q:16 | 28 | 0.39 | rs1367026482:16 | *:16 | Caa/Taa:16 | 15:83103190_16 | 0.03 | 0.05 | 2.07E-33 | FALSE | 1.26 | 1.29 | 1.27 | FALSE |
| *HLX* | 235 | -28.59 | 15 | A:15 | 42 | 0.28 | novel:15 | V:15 | gCA/gTC:15 | 1:221054647_15 | NA | 0.06 | 8.51E-26 | FALSE | 1.30 | 1.34 | 1.31 | FALSE |
| *DYNC1LI1* | 447 | -21.78 | 12 | F:12 | 45 | 0.44 | novel:12 | A:12 | TTc/GCc:12 | 3:32570060_12 | NA | 0.04 | 2.94E-19 | FALSE | 1.28 | 1.30 | 1.36 | FALSE |
| *CD5* | 461 | -19.92 | 10 | H:10 | 24 | 0.29 | rs386754125:10 | C:10 | CAc/TGc:10 | 11:60892605_10 | NA | 0.05 | 4.07E-17 | FALSE | 1.23 | 1.19 | 1.31 | FALSE |
| *ZIC2* | 324 | -17.51 | 7 | I:7 | 18 | 0.60 | novel:7 | V:6\|L:1 | Atc/Gtc:6\|Atc/Ctc:1 | 13:100635288_7 | 0.02 | 0.06 | 1.12E-14 | FALSE | 1.32 | 1.21 | 1.29 | FALSE |
| *NBPF14* | 1469 | -16.61 | 6 | A:6 | 25 | 0.14 | rs1400687470:6 | V:6 | gCa/gTa:6 | 1:146445430_6 | 0.03 | 0.05 | 6.28E-13 | FALSE | 1.12 | 1.34 | 1.35 | FALSE |
| *CCDC60* | 547 | -13.59 | 7 | S:7 | 22 | 0.17 | novel:7 | K:7 | aGC/aAA:7 | 12:119978507_7 | NA | 0.05 | 9.68E-11 | FALSE | 1.26 | 1.24 | 1.29 | FALSE |
| *DMBT1* | 229 | -13.89 | 6 | E:6 | 78 | 0.51 | rs780403364:6 | D:6 | gaA/gaT:6 | 10:124339101_6 | 0.02 | 0.05 | 2.74E-10 | FALSE | 1.30 | 1.34 | 1.34 | FALSE |
| *KIF4B* | 184 | -12.99 | 6 | A:6 | 38 | 0.20 | rs372621246:6 | T:6 | Gcc/Acc:6 | 5:154393969_6 | 0.03 | 0.04 | 9.81E-10 | FALSE | 1.12 | 1.25 | 1.32 | FALSE |
| *SAMD9* | 286 | -12.74 | 6 | E:6 | 38 | 0.67 | rs765431657:6 | D:6 | gaG/gaT:6 | 7:92734553_6 | 0.04 | 0.04 | 2.32E-09 | FALSE | 1.36 | 1.29 | 1.31 | FALSE |
| *DMBT1* | 230 | -12.01 | 6 | S:6 | 78 | 0.53 | novel:6 | S:6 | tcCAgt/tcTGgt:6 | 10:124339104_6 | NA | 0.05 | 6.95E-09 | FALSE | 1.30 | 1.34 | 1.32 | FALSE |
| *DMBT1* | 231 | -12.01 | 6 | S:6 | 78 | 0.53 | novel:6 | G:6 | tcCAgt/tcTGgt:6 | 10:124339104_6 | NA | 0.05 | 6.95E-09 | FALSE | 1.30 | 1.34 | 1.32 | FALSE |
| *ANKRD50* | 1305 | -11.49 | 5 | D:5 | 43 | 0.26 | rs569034621:5 | G:5 | gAt/gGt:5 | 4:125590518_5 | 0.02 | 0.04 | 3.59E-08 | FALSE | 1.17 | 1.27 | 1.31 | FALSE |
| *ZBED3* | 155 | -10.47 | 6 | L:6 | 13 | 0.14 | | A:6 | CTg/GCg:6 | 5:76373240_6 | NA | 0.05 | 4.75E-08 | FALSE | 1.17 | 1.17 | 1.14 | FALSE |
| *ARPP21* | 343 | -11.08 | 6 | G:6 | 25 | 0.03 | rs765166691:6 | E:6 | gGg/gAg:6 | 3:35763129_6 | 0.06 | 0.04 | 5.20E-08 | FALSE | 1.26 | 1.28 | 1.30 | FALSE |
| *CEP55* | 430 | -10.43 | 5 | T:5 | 14 | 0.14 | rs572620662:5 | I:5 | aCt/aTt:5 | 10:95287804_5 | 0.03 | 0.04 | 1.15E-07 | FALSE | 1.17 | 1.25 | 1.28 | FALSE |
| *ZNF394* | 561 | -10.49 | 6 | L:6 | 31 | 0.56 | rs1463661588:5\|<br>novel:1 | L:1 | ctA/ctG:5\|ctATaa/ctGCaa:1 | 7:99091155_5\|<br>7:99091154_1 | NA | 0.05 | 1.26E-07 | FALSE | 1.26 | 1.27 | 1.33 | FALSE |
| *PACS1* | 274 | -10.48 | 5 | I:5 | 46 | 0.46 | rs1445829899:5 | T:5 | aTt/aCt:5 | 11:65984006_5 | 0.02 | 0.05 | 2.38E-07 | FALSE | 1.17 | 1.30 | 1.30 | FALSE |
| *POLQ* | 187 | -10.17 | 5 | A:5 | 88 | 0.69 | rs2048474462:5 | T:5 | Gca/Aca:5 | 3:121258352_5 | 0.03 | 0.04 | 2.93E-07 | FALSE | 1.28 | 1.29 | 1.32 | FALSE |
| *MAP3K19* | 1282 | -10.40 | 5 | H:5 | 43 | 0.33 | novel:5 | Y:5 | Cac/Tac:5 | 2:135738467_5 | 0.03 | 0.04 | 4.08E-07 | FALSE | 1.26 | 1.36 | 1.34 | FALSE |
| *DNAH14* | 1098 | -10.49 | 5 | K:5 | 98 | 0.29 | novel:5 | K:5 | aaAAac/aaGTac:5 | 1:225270408_5 | NA | 0.04 | 6.66E-07 | FALSE | 1.23 | 1.29 | 1.34 | FALSE |
| *DNAH14* | 1099 | -10.49 | 5 | N:5 | 98 | 0.29 | novel:5 | Y:5 | aaAAac/aaGTac:5 | 1:225270408_5 | NA | 0.04 | 6.66E-07 | FALSE | 1.23 | 1.29 | 1.34 | FALSE |
| *KRT5* | 232 | -9.79 | 5 | S:5 | 23 | 0.73 | rs200333163:5 | N:5 | aGc/aAc:5 | 12:52912805_5 | 0.04 | 0.05 | 6.70E-07 | FALSE | 1.16 | 1.19 | 1.27 | FALSE |
| *PER1* | 699 | -10.00 | 5 | V:5 | 49 | 0.56 | novel:5 | L:5 | Gtg/Ctg:5 | 17:8049399_5 | 0.04 | 0.05 | 9.92E-07 | FALSE | 1.23 | 1.23 | 1.20 | FALSE |
| *RPL8* | - | -9.15 | 5 | SS:5 | 17 | 0.77 | rs112091443:5 | SS:5 | :5 | 8:146015348_5 | 0.04 | 0.06 | 1.12E-06 | FALSE | 1.28 | 1.32 | 1.37 | FALSE |
| *PLXNB3* | 769 | -9.82 | 5 | Q:5 | 77 | 0.00 | novel:4\|<br>rs782126891:1 | *:4\|R:1 | Cag/Tag:4\|cAg/cGg:1 | X:153036816_4\|<br>X:153036817_1 | 0.04 | 0.05 | 2.33E-06 | FALSE | 1.23 | 1.24 | 1.27 | FALSE |
| *ATP2A3* | 745 | -9.39 | 5 | A:5 | 52 | 0.19 | rs764213441:4\|<br>novel:1 | T:4\|V:1 | Gct/Act:4\|gCt/gTt:1 | 17:3840798_4\|<br>17:3840797_1 | 0.04 | 0.06 | 3.04E-06 | FALSE | 1.16 | 1.17 | 1.31 | FALSE |
| *TGFBI* | 29 | -9.17 | 5 | P:5 | 31 | 0.06 | novel:5 | T:5 | Ccc/Acc:5 | 5:135364829_5 | 0.04 | 0.05 | 3.25E-06 | FALSE | 1.23 | 1.21 | 1.14 | FALSE |
| *EEF2KMT* | 233 | -8.46 | 5 | V:5 | 23 | 0.49 | rs112774542:5 | I:5 | Gtc/Atc:5 | 16:5140130_5 | 0.04 | 0.05 | 7.31E-06 | FALSE | 1.26 | 1.27 | 1.30 | FALSE |
| *CPSF2* | 71 | -8.61 | 5 | L:5 | 27 | 0.86 | rs1595053184:5 | I:5 | Ctc/Atc:5 | 14:92600416_5 | 0.06 | 0.04 | 1.21E-05 | FALSE | 1.26 | 1.24 | 1.25 | FALSE |
| *FLNB* | 217 | -9.14 | 5 | P:5 | 95 | 0.42 | rs2106965787:5 | L:5 | cCt/cTt:5 | 3:58067366_5 | 0.06 | 0.05 | 1.60E-05 | FALSE | 1.26 | 1.32 | 1.28 | FALSE |
| *DEPDC1B* | 512 | -8.17 | 5 | W:5 | 21 | 0.28 | rs781414143:5 | *:5 | tgG/tgA:5 | 5:59893634_5 | 0.06 | 0.04 | 2.45E-05 | FALSE | 1.23 | 1.33 | 1.29 | FALSE |
| *TP53* | 248 | -7.39 | 9 | R:9 | 85 | 0.46 | rs121912651:5\|<br>rs11540652:4 | W:5\|Q:3\|L:1 | Cgg/Tgg:5\|cGg/cAg:3\|cGg/cTg:1 | 17:7577539_5\|<br>17:7577538_4 | 0.15 | 0.05 | 5.23E-05 | FALSE | 1.26 | 1.31 | 1.34 | TRUE |
| *MATN1* | 438 | -7.37 | 5 | K:5 | 20 | 0.64 | novel:5 | N:5 | aaG/aaT:5 | 1:31188050_5 | 0.11 | 0.05 | 1.45E-04 | FALSE | 1.18 | 1.25 | 1.28 | FALSE |
| *SLFN13* | 435 | -7.27 | 5 | S:5 | 32 | 0.22 | rs778646600:5 | F:5 | tCc/tTc:5 | 17:33769200_5 | 0.11 | 0.04 | 1.79E-04 | FALSE | 1.26 | 1.32 | 1.34 | FALSE |
| *SDHA* | 127 | -7.19 | 5 | V:5 | 23 | 0.73 | rs777600956:3\|<br>novel:2 | M:3 | Gtg/Atg:3\|gtG/gtC:2 | 5:225600_3\|<br>5:225602_2 | 0.15 | 0.05 | 3.05E-04 | FALSE | 1.28 | 1.24 | 1.31 | FALSE |
| *ARHGAP39* | 296 | -7.15 | 5 | S:5 | 49 | 0.23 | rs771995320:4\|<br>novel:1 | Y:1 | tcC/tcT:4\|tCc/tAc:1 | 8:145773582_4\|<br>8:145773583_1 | 0.14 | 0.06 | 5.99E-04 | FALSE | 1.23 | 1.17 | 1.20 | FALSE |
| *RNF148* | 13 | -6.31 | 5 | S:5 | 17 | 0.19 | rs762346587:5 | F:5 | tCt/tTt:5 | 7:122342767_5 | 0.11 | 0.04 | 9.46E-04 | FALSE | 1.23 | 1.31 | 1.27 | FALSE |
| *TP53* | 245 | -5.69 | 6 | G:6 | 85 | 0.76 | rs121912656:4\|<br>rs28934575:2 | D:3\|S:2\|V:1 | gGc/gAc:3\|Ggc/Agc:2\|gGc/gTc:1 | 17:7577547_4\|<br>17:7577548_2 | 0.08 | 0.05 | 1.76E-03 | FALSE | 1.26 | 1.33 | 1.30 | TRUE |
| *CDC20* | 162 | -6.25 | 5 | R:5 | 25 | 0.33 | rs200111540:5 | Q:5 | cGg/cAg:5 | 1:43825697_5 | 0.15 | 0.05 | 1.91E-03 | FALSE | 1.26 | 1.31 | 1.31 | FALSE |
| *CDK16* | 9 | -5.69 | 5 | R:5 | 32 | 0.40 | | L:5 | cGg/cTg:5 | X:47082982_5 | 0.15 | 0.05 | 6.80E-03 | FALSE | 1.23 | 1.26 | 1.34 | FALSE |
| *EEF2KMT* | 231 | -5.18 | 5 | A:5 | 23 | 0.56 | rs74684799:5 | T:5 | Gcg/Acg:5 | 16:5140136_5 | 0.17 | 0.05 | 6.93E-03 | FALSE | 1.32 | 1.29 | 1.30 | FALSE |
| *ADAMTS16* | 630 | -5.47 | 5 | R:5 | 67 | 0.24 | rs377507889:4\|<br>rs200111235:1 | H:4\|C:1 | cGc/cAc:4\|Cgc/Tgc:1 | 5:5235165_4\|<br>5:5235164_1 | 0.20 | 0.05 | 3.22E-02 | FALSE | 1.32 | 1.36 | 1.36 | FALSE |

AA: Amino acid position; C = mutation count; R-AA = Reference amino-acid; TMG= Total mutation in gene; MAFR= Median allele freq rank; V-AA = variant amino-acid; CC= Codon changes

**Supplementary table 8:** Known hotspot mutations identified in the current study

| | | | | | Number of patients with mutation in >= 10% of reads | | | | Number (%) of control tissue samples with mutation in >= 10% of reads. "." means mutation was not identified in normal tissue samples |
|---|---|---|---|---|---|---|---|---|---|
| Chromosome | Location (hg38) | Alleles | Amino acid change | Gene | Cohort A | Cohort B | Cohort C | Total (%) | |
| *2* | *203867991* | *A>G* | *T17A* | *CTLA4* | *15* | *27* | *17* | *59 (57%)* | 16 (57%) |
| 3 | 179199088 | G>A | R88Q | *PIK3CA* | 0 | 0 | 1 | 1 (1%) | . |
| 3 | 179199157 | A>G | K111R | *PIK3CA* | 0 | 1 | 0 | 1 (1%) | . |
| 3 | 179203764 | A>C | N345T | *PIK3CA* | 0 | 1 | 1 | 2 (2%) | . |
| 3 | 179204513 | G>A | R357Q | *PIK3CA* | 0 | 1 | 0 | 1 (1%) | . |
| 3 | 179210192 | T>C | C420R | *PIK3CA* | 0 | 1 | 0 | 1 (1%) | . |
| 3 | 179210291 | G>A | E453K | *PIK3CA* | 0 | 1 | 0 | 1 (1%) | . |
| 3 | 179218294 | G>A | E542K | *PIK3CA* | 0 | 1 | 0 | 1 (1%) | . |
| 3 | 179218303 | G>A | E545K | *PIK3CA* | 1 | 1 | 0 | 2 (2%) | . |
| 3 | 179230256 | A>G | D939G | *PIK3CA* | 0 | 1 | 0 | 1 (1%) | . |
| 3 | 179234302 | G>C | G1049R | *PIK3CA* | 0 | 1 | 0 | 1 (1%) | . |
| *8* | *127738434* | *A>C* | *T73P* | *MYC* | *0* | *32* | *21* | *53 (51%)* | . |
| 9 | 21971018 | G>T | P114H | *CDKN2A* | 0 | 0 | 1 | 1 (1%) | . |
| 9 | 21971029 | C>T | W110* | *CDKN2A* | 1 | 0 | 0 | 1 (1%) | . |
| 9 | 21971030 | C>T | W110* | *CDKN2A* | 0 | 0 | 1 | 1 (1%) | . |
| 9 | 21971112 | G>A | H83Y | *CDKN2A* | 0 | 2 | 1 | 3 (3%) | . |
| 9 | 21971121 | G>A | R80* | *CDKN2A* | 4 | 4 | 0 | 8 (8%) | . |
| 9 | 21971187 | G>A | R58* | *CDKN2A* | 1 | 0 | 0 | 1 (1%) | . |
| 9 | 21974793 | G>T | S12* | *CDKN2A* | 0 | 0 | 2 | 2 (2%) | . |
| 9 | 77922196 | T>A | T96S | *GNAQ* | 4 | 5 | 0 | 9 (9%) | 2 (7%) |
| 11 | 72238043 | C>A | A1185D | *INPPL1* | 0 | 4 | 15 | 19 (18%) | . |
| *12* | *120978847* | *A>C* | *I27L* | *HNF1A* | *15* | *34* | *16* | *65 (63%)* | 15 (54%) |
| 12 | 120988846 | C>T | R114C | *HNF1A* | 0 | 1 | 0 | 1 (1%) | . |
| 13 | 20988344 | ->GGGGCG | P479_A480dup | *LATS2* | 18 | 0 | 0 | 18 (17%) | 15 (54%) |
| 13 | 109782884 | C>T | G1057D | *IRS2* | 13 | 27 | 5 | 45 (44%) | 11 (39%) |
| 15 | 40382870 | T>G | V12G | *KNSTRN* | 0 | 0 | 15 | 15 (15%) | . |
| 17 | 7670684 | C>G | R342P | *TP53* | 0 | 1 | 0 | 1 (1%) | . |
| 17 | 7670685 | G>A | R342* | *TP53* | 1 | 1 | 0 | 2 (2%) | . |
| 17 | 7670700 | G>A | R337C | *TP53* | 0 | 1 | 1 | 2 (2%) | . |
| 17 | 7673537 | G>A | Q331* | *TP53* | 0 | 0 | 1 | 1 (1%) | . |
| 17 | 7673704 | G>A | R306* | *TP53* | 1 | 0 | 2 | 3 (3%) | . |
| 17 | 7673764 | C>T | E286K | *TP53* | 1 | 0 | 0 | 1 (1%) | . |
| 17 | 7673767 | C>A | E285* | *TP53* | 1 | 0 | 0 | 1 (1%) | . |
| 17 | 7673775 | C>T | R282Q | *TP53* | 1 | 0 | 0 | 1 (1%) | . |
| 17 | 7673776 | G>A | R282W | *TP53* | 2 | 0 | 0 | 2 (2%) | . |
| 17 | 7673782 | T>A | R280* | *TP53* | 0 | 1 | 0 | 1 (1%) | . |
| 17 | 7673805 | A>C | V272G | *TP53* | 0 | 1 | 0 | 1 (1%) | . |
| 17 | 7674197 | TGA>- | I255del | *TP53* | 0 | 0 | 1 | 1 (1%) | . |
| 17 | 7674217 | C>A | R249M | *TP53* | 0 | 1 | 0 | 1 (1%) | . |
| 17 | 7674220 | C>T | R248Q | *TP53* | 1 | 1 | 1 | 3 (3%) | . |
| 17 | 7674221 | G>A | R248W | *TP53* | 0 | 5 | 0 | 5 (5%) | . |
| 17 | 7674229 | C>A | G245V | *TP53* | 1 | 0 | 0 | 1 (1%) | . |
| 17 | 7674229 | C>T | G245D | *TP53* | 0 | 1 | 1 | 2 (2%) | . |
| 17 | 7674230 | C>T | G245S | *TP53* | 0 | 1 | 1 | 2 (2%) | . |
| 17 | 7674257 | A>T | Y236N | *TP53* | 0 | 0 | 1 | 1 (1%) | . |
| 17 | 7674263 | A>T | Y234N | *TP53* | 1 | 1 | 0 | 2 (2%) | . |
| 17 | 7674268 | A>T | I232N | *TP53* | 0 | 2 | 0 | 2 (2%) | . |
| 17 | 7674893 | C>T | R213Q | *TP53* | 1 | 0 | 1 | 2 (2%) | . |
| 17 | 7674894 | G>A | R213* | *TP53* | 0 | 1 | 1 | 2 (2%) | . |
| 17 | 7674945 | G>A | R196* | *TP53* | 0 | 1 | 0 | 1 (1%) | . |
| 17 | 7675071 | G>A | R181C | *TP53* | 0 | 1 | 0 | 1 (1%) | . |
| 17 | 7675077 | G>A | H179Y | *TP53* | 0 | 1 | 0 | 1 (1%) | . |
| 17 | 7675088 | C>T | R175H | *TP53* | 1 | 1 | 2 | 4 (4%) | . |
| 17 | 7675093 | C>- | P177_C182del | *TP53* | 1 | 0 | 0 | 1 (1%) | . |
| 17 | 7675094 | A>G | V173A | *TP53* | 1 | 0 | 0 | 1 (1%) | . |
| 17 | 7675130 | G>T | A161D | *TP53* | 0 | 0 | 1 | 1 (1%) | . |
| 17 | 7675140 | G>A | R158C | *TP53* | 0 | 1 | 0 | 1 (1%) | . |
| 17 | 7675143 | C>A | V157F | *TP53* | 0 | 0 | 1 | 1 (1%) | . |
| 17 | 7675175 | C>T | W146* | *TP53* | 0 | 1 | 0 | 1 (1%) | . |
| 17 | 7675995 | G>A | T125M | *TP53* | 0 | 1 | 1 | 2 (2%) | . |
| 17 | 7676055 | C>T | G105D | *TP53* | 0 | 4 | 0 | 4 (4%) | . |

**Supplementary table 9:** List of the CNVs identified.

| Cohort A, Amplification | | | |
|---|---|---|---|
| **cytoband** | **q value** | **wide peak boundaries** | **gene** |
| 11q13.2 | 1.0565E-06 | chr11:67981801-68059008 | *ALDH3B1,CHKA,NDUFS8,TCIRG1,UNC93B1,MIR4691,MIR6753,MIR7113* |
| 7p11.2 | 0.000034596 | chr7:53035731-56088899 | *CCT6A,**EGFR**,GBAS,HPVC1,PHKG1,PSPH,SEC61G,SUMF2,MRPS17,LANCL2,VOPP1,VSTM2A,POM121L12,VSTM2A-OT1,SEPT14,ZNF713,FKBP9P1,LINC01446,SNORA15,EGFR-AS1,LOC100996654,LINC01445,ELDR* |
| 9p13.3 | 0.030693 | chr9:35395165-35863227 | *CA9,CD72,NPR2,RMRP,TESK1,TLN1,TPM2,RGP1,RUSC2,CREB3,UNC13B,SPAG8,SIT1,TMEM8B,GBA2,HINT2,ARHGEF39,LINC00950,ATP8B5P,CCDC107,FAM221B,MSMP,FAM166B,MIR4667,LOC101926948,MIR6852,MIR6853* |
| **Cohort B, Amplification** | | | |
| **cytoband** | **q value** | **wide peak boundaries** | **gene** |
| 12q12 | 9.1954E-21 | chr12:40486116-40488194 | *MUC19* |
| 7q22.1 | 7.4474E-14 | chr7:101017456-101034348 | *MUC12,MUC17,LOC102724094* |
| 6p22.2 | 5.2522E-13 | chr6:26199856-26258924 | *HIST1H1D,HIST1H2AE,HIST1H2BG,HIST1H2BF,HIST1H2BH,HIST1H3E,HIST1H4F,HIST1H4E,HIST1H4G,HIST1H3F* |
| 11q13.2 | 3.7095E-11 | chr11:66328021-67455782 | *ACTN3,GRK2,BBS1,SLC29A2,PC,PPP1CA,PTPRCAP,RAD9A,RBM4,RPS6KB2,SPTBN2,CTSF,RIN1,CCS,RCE1,DPP3,RBM14,B4GAT1,KDM2A,CLCF1,BRMS1,RHOD,SSH3,CCDC87,CABP4,CORO1B,CARNS1,POLD4,MRPL11,LRFN4,C11orf80,RBM4B,SYT12,PELI3,ZDHHC24,C11orf86,NPAS4,ANKRD13D,TBC1D10C,GPR152,LOC100130987,MIR3163,RBM14-RBM4,LOC101928069,MIR6860,LOC102724064* |
| 8q24.3 | 7.5105E-11 | chr8:143873236-144276833 | *CYC1,GRINA,PLEC,GPAA1,BOP1,OPLAH,HGH1,EXOSC4,SHARPIN,EPPK1,MAF1,PARP10,WDR97,SPATC1,SCX,MIR661,MROH1,MIR6846,MIR6847,MIR7112* |
| 5p15.1 | 4.2759E-10 | chr5:14759537-17275829 | *MYO10,BASP1,FBXL7,FAM134B,ANKH,ZNF622,LOC285696,MARCH11,MIR887,MIR4637,LOC101929454,CTD-2350J17.1,LOC101929505,LOC101929524* |
| 19p13.2 | 5.7824E-09 | chr19:8961232-8972039 | *MUC16* |
| 1q21.3 | 5.7824E-09 | chr1:152710412-152761181 | *KPRP,C1orf68* |
| 6q25.3 | 2.4644E-06 | chr6:159222310-159234404 | *FNDC1* |
| 19q13.31 | 2.9344E-06 | chr19:43013505-43075489 | *PSG2,PSG11* |
| 1p34.1 | 3.7234E-07 | chr1:43910865-44228958 | *ATP6V0B,DPH2,ST3GAL3,SLC6A9,B4GALT2,ARTN,IPO13,DMAP1,ERI3,KLF17,CCDC24,ERI3-IT1* |
| 14q11.2 | 7.0917E-06 | chr14:21467821-22780674 | *DAD1,OXA1L,SALL2,SLC7A7,TOX4,OR10G3,OR10G2,OR4E2,OR4E1,METTL3,ABHD4,RAB2B* |
| 16q22.1 | 0.000091421 | chr16:67817885-68255714 | *CTRL,LCAT,NFATC3,PSKH1,PSMB10,SLC12A4,NUTF2,EDC4,PLA2G15,DUS2,DDX28,TSNAXIP1,THAP11,DPEP2,DPEP3,ESRP2,CENPT,NRN1L,LOC100131303,MIR6773* |
| 10q22.2 | 0.00025067 | chr10:73753352-73799318 | *SEC24C,ZSWIM8,CHCHD1,FUT11,ZSWIM8-AS1* |
| 9q34.13 | 0.000032224 | chr9:131188490-131648533 | *RAPGEF1,NUP214,POMT1,SNORD62A,UCK1,PRRC2B,PLPP7,FAM78A* |
| 3q29 | 0.00062719 | chr3:196218805-198295559 | *BDH1,DLG1,MELTF,PAK2,PCYT1A,RPL35A,RUBCN,NCBP2,UBXN7,PIGX,PIGZ,IQCG,FYTTD1,LRCH3,CEP19,LMLN,TM4SF19,NCBP2-AS2,RNF168,SLC51A,FBXO45,SENP5,LOC220729,TCTEX1D2,SMCO1,WDR53,ANKRD18DP,NRROS,FAM157A,MIR922,MFI2-AS1,DLG1-AS1,TM4SF19-TCTEX1D2,MIR4797,LMLN-AS1,UBXN7-AS1,TM4SF19-AS1,LINC01063* |
| 20q13.33 | 3.1581E-06 | chr20:63668692-63874347 | *TPD52L2,TNFRSF6B,ARFRP1,RTEL1,LIME1,SLC2A4RG,ZGPAT,ZBTB46,ABHD16B,RTEL1-TNFRSF6B,ZBTB46-AS1* |
| 17p13.2 | 0.0013158 | chr17:4732452-4884989 | *PLD2,PSMB6,TM4SF5,MINK1,CXCL16,ZMYND15,VMO1,GLTPD2,MED11* |
| 16p13.3 | 0.0016862 | chr16:625064-667565 | *RAB40C,METTL26,MCRIP2,WFIKKN1,WDR90,LOC105371038* |
| 5q35.3 | 0.000079032 | chr5:181195431-181538259 | *RACK1,OR4F16,TRIM7,TRIM52,TRIM41,SNORD95,SNORD96A,OR4F29,LOC100132062,TRIM52-AS1,MIR4638,CTC-338M12.4* |
| 17q25.3 | 0.006434 | chr17:78411537-78985238 | *LGALS3BP,TIMP2,DNAH17,CYTH1,PGS1,USP36,CEP295NL,DNAH17-AS1,LOC101928710* |
| 20q11.23 | 0.00071901 | chr20:38082186-38234441 | *TGM2,RPRD1B,KIAA1755* |
| 12p13.31 | 0.014179 | chr12:8857184-9166162 | *A2M,PHC1,M6PR,PZP,KLRG1,A2ML1,A2M-AS1,LINC00612* |
| 15q25.3 | 0.01492 | chr15:85758232-87943023 | *NTRK3,KLHL25,AGBL1,LINC00052,AGBL1-AS1,MIR1276,MIR548AP,LOC101929679,LINC01584,LOC102724452,LOC105370954* |
| 11p15.5 | 0.0054854 | chr11:1-248984 | *PSMD13,SIRT3,BET1L,RIC8A,ODF3,SCGB1C1,LINC01001,MIR6743* |
| 2q35 | 0.025947 | chr2:215428841-219002991 | *AAMP,BCS1L,CRYBA2,CYP27A1,FN1,IGFBP2,IGFBP5,CXCR1,CXCR2,CXCR2P1,RPL37A,SLC11A1,TNP1,TNS1,VIL1,WNT6,XRCC5,ZNF142,CDK5R2,CNOT9,TTLL4,ARPC2,PNKD,STK36,SMARCAL1,PRKAG3,FEV,MREG,PECR,MARCH4,USP37,CTDSP1,TMBIM1,RNF25,WNT10A,PLCD4,TMEM169,PKI55,LINC00608,GPBAR1,CFAP65,RUFY4,CATIP,MIR26B,MIR375,LINC00607,DIRC3,LOC100129175,LINC01280,LOC101928327,CATIP-AS1,LINC01494,MIR6513,MIR6809,MIR6810,LOC102724849,MIR9500,CATIP-AS2,LINC01614,LOC105373876,DIRC3-AS1,LOC105373878* |
| 22q13.33 | 0.025947 | chr22:50623156-50818468 | *ACR,ARSA,RABL2B,SHANK3,RPL23AP82,LOC105373100* |
| 11p13 | 0.016798 | chr11:33542578-43317792 | *CAT,CD44,CD59,ELF5,LMO2,CAPRIN1,RAG1,RAG2,SLC1A2,TRAF6,PDHX,API5,FJX1,KIAA1549L,ABTB2,PAMR1,FBXO3,EHF,COMMD9,APIP,TRIM44,NAT10,LRRC4C,PRR5L,C11orf74,LDLRAD3,HNRNPKP3,C11orf91,LOC100507144,LOC100507205,MIR3973,MIR1343,FBXO3-AS1,LOC101928510,LINC01493,LOC101928591,LINC01499,LOC103312105,LOC105376633* |
| 12q14.3 | 0.010286 | chr12:65453631-70619723 | *CPM,IFNG,LYZ,MDM2,CNOT2,PTPRB,RAP1B,YEATS4,HMGA2,DYRK2,CCT2,FRS2,CPSF6,IRAK3,GRIP1,KCNMB4,IL22,TMBIM4,SLC35E3,IL26,CAND1,MDM1,NUP107,LLPH,HELB,RAB3IP,BEST3,MYRFL,RPSAP52,MSRB3,LRRC10,LOC100129940,LOC100130075,MIR1279,SNORA70G,MIR3913-2,MIR3913-1,LOC100507065,LOC100507175,LOC100507195,LOC100507250,IFNG-AS1,LOC101927901,LINC01479,LOC101928002,LINC01481,MIR6074,MIR6502,LOC102724421,LLPH-AS1,LOC105369187* |
| 14q11.2 | 0.00028223 | chr14:19739156-19936454 | *OR4K5,OR4K1,OR4N2,OR4K2,OR4Q3,OR4M1* |
| 9q21.13 | 0.0048599 | chr9:73149588-94941714 | *ANXA1,AUH,CKS2,CTSL,CTSLP8,DAPK1,ECM2,S1PR3,FBP1,GAS1,GCNT1,GNAQ,HNRNPK,IARS,NFIL3,NINJ1,NTRK2,ROR2,OMD,OGN,PCSK5,PHF2,RORB,SYK,TLE1,TLE4,FBP2,GNA14,SEMA4D,SPTLC1,GADD45G,SPIN1,FAM120A,VPS13A,AGTPBP1,BICD2,CDK20,OSTF1,PSAT1,UBQLN1,PCA3,GOLM1,SHC3,NUTM2F,DIRAS2,ASPN,NMRK1,NOL8,C9orf40,RFK,KIF27,BARX1,NAA35,SLC28A3,IPPK,WNK2,SECISBP2,ZCCHC6,RMI1,GKAP1,ISCA1,ZNF484,CEP78,C9orf64,CARD19,MFSD14B,C9orf3,FGD3,CARNMT1,PTPDC1,ANKRD19P,TRPM6,NXNL2,RASEF,MIRLET7DHG,FAM120AOS,LINC00475,PRUNE2,ZNF169,SUSD3,FRMD3,C9orf47,SPATA31E1,LOC286238,MIR4290HG,LINC01501,SPATA31D5P,SPATA31D4,SPATA31D3,SPATA31D1,LOC389765,C9orf153,CTSL3P,LOC392364,C9orf170,CENPP,MIRLET7A1,MIRLET7D,MIRLET7F1,MIR7-1,IDNK,LOC440173,SPATA31C1,FOXB2,C9orf129,LOC494127,LOC642943,SPATA31C2,RPSAP9,SNORA84,LOC100128076,LOC100128361,UNQ6494,LOC100129316,LINC00484,LOC100132077,VPS13A-AS1,MIR2278,MIR4291,MIR3153,MIR4290,MIR4289,MIR3910-1,MIR3910-2,MIR3651,GAS1RR,MIR4670,MIR548AU,LOC101927358,C9orf41-AS1,GNA14-AS1,LOC101927450,LINC01507,LOC101927502,LOC101927575,LOC101927623,LOC101927847,LINC01508,LOC101927954,PCAT7,LOC101929748,LOC102724156,RORB-AS1,LOC105376114* |
| 5q31.3 | 0.0023294 | chr5:141467371-154063231 | *ADRB2,ANXA6,ATOX1,CAMK2A,CD74,CDX1,CSF1R,CSNK1A1,DIAPH1,DPYSL3,SLC26A2,FAT2,FGF1,GLRA1,GM2A,GPX3,GRIA1,NR3C1,NDST1,HTR4,MFAP3,PCDH1,PCDHGC3,PDE6A,PDGFRB,POU4F3,PPP2R2B,RPS14,SLC6A7,SPARC,SPINK1,TCOF1,PCDHGB4,HDAC3,RNF14,PCDHGA8,KIAA0141,JAKMIP2,GNPDA1,G3BP1,TNIP1,FAM114A2,TCERG1,SPINK5,SYNPO,ABLIM3,HMGXB3,ARHGAP26,PCDHGA12,CCDC69,IL17B,DCTN4,PCDH12,LARS,RBM27,RBM22,PCDHGC5,PCD* |

| | | | |
|---|---|---|---|
| | | | HGC4,PCDHGB7,PCDHGB6,PCDHGB5,PCDHGB3,PCDHGB2,PCDHGB1,PCDHGA11,PCDHGA10,PCDHGA9,PCDHGA7,PCDHGA6,PCDHGA5,PCDHGA4,PCDHGA3,PCDHGA2,PCDHGA1,NMUR2,KCTD16,HMHB1,ARAP3,PCYOX1L,SH3TC2,NDFIP1,FBXO38,YIPF5,TIGD6,SPRY4,SPINK7,SMIM3,FCHSD1,ZNF300,MYOZ3,SCGB3A2,PPARGC1B,AFAP1L1,GRPEL2,GPR151,ZNF300P1,SLC36A2,SPINK13,JAKMIP2-AS1,PRELID2,SH3RF2,PLAC8L1,STK32A,SLC36A1,LOC255187,RELL2,SLC36A3,ARSI,IRGM,C5orf46,ARHGEF37,SPINK6,MIR143,MIR145,SPINK14,MIR378A,GRXCR2,SPINK9,LOC644762,MIR584,CARMN,LOC729080,LOC100505658,SPRY4-IT1,LOC100652758,MIR5197,ARHGAP26-AS1,PPP2R2B-IT1,ARHGAP26-IT1,LOC101926941,LOC101926975,CTB-113P19.1,CTB-12O2.1,LINC01470,MIR6499,LOC102546294,LOC102546298,GRPEL2-AS1 |
| 1q21.1 | 0.0074128 | chr1:110239416-152094427 | ADORA3,AMPD1,RHOC,ARNT,ATP1A1,ATP5F1,BCL9,CAPZA1,CASQ2,CD2,CD53,CD58,CHI3L2,CTSK,CTSS,DRD5P2,ECM1,ENSA,FCGR1A,FCGR1B,FMO5,GJA5,GJA8,HMGCS2,HSD3B1,HSD3B2,IGSF3,KCNA2,KCNA3,KCNA10,KCND3,MCL1,MOV10,NGF,NHLH2,NOTCH2,NRAS,OVGP1,PDZK1,PI4KB,PRKAB2,PSMB4,PSMD4,PTGFRN,RAP1A,RFX5,RNU1-4,RORC,S100A10,S100A11,SLC16A1,SYCP1,TBX15,VPS72,TSHB,TUFT1,WNT2B,CSDE1,HIST2H2AA3,HIST2H2AC,HIST2H2BE,HIST2H4A,PIP5K1A,ANXA9,TTF2,ITGA10,PEX11B,SELENBP1,SLC16A4,PRPF3,CD101,SEC22B,CHD1L,PDE4DIP,LRIG2,SETDB1,SV2A,RBM8A,TSPAN2,SF3B4,BCAS2,WARS2,CEPT1,PIAS3,SEMA6C,LAMTOR5,POLR3C,TXNIP,AP4B1,PHTF1,WDR3,MTMR11,MAN1A2,MLLT11,TDRKH,ADAM30,CD160,CELF3,DDX20,VPS45,POGZ,RPRD2,CA14,NBPF14,PTPN22,PHGDH,RNVU1-7,RNU1-3,RNU1-2,RNU1-1,CHIA,RNF115,TMOD4,CERS2,BOLA1,APH1A,PLEKHO1,HAO2,ACP6,GPR89B,TRIM33,OAZ3,MRPS21,ADAMTSL4,RSBN1,GDAP2,FAM46C,ST7L,C1orf56,GOLPH3L,SLC22A15,LRIF1,FAM63A,CTTNBP2NL,FAM212B,CDC42SE1,OLFML3,OTUD7B,LINC00869,TMIGD3,CGN,ZNF687,PRUNE1,RBM15,DCLRE1B,MRPL9,SCNM1,WDR77,TNFAIP8L2,C1orf54,VTCN1,DENND2D,SIKE1,TARS2,TRIM45,SNX27,ANP32E,VANGL1,REG4,HORMAD1,POLR3GL,PROK1,ATP1A1-AS1,ZNF697,GNRHR2,THEM4,GABPB2,TCHHL1,MAB21L3,LIX1L,HSD3BP4,DRAM2,PIFO,C1orf162,SYT6,CIART,HFE2,ANKRD35,NBPF12,BNIPL,CHIAP2,DENND2C,NUDT4P2,PDIA3P1,NBPF11,NUDT17,SPAG17,HIPK1,AKR7A2P1,MAGI3,FAM19A3,RIIAD1,THEM5,NBPF15,ANKRD34A,HIST2H2AB,PPM1J,HIST2H3A,HIST2H2BC,HIST2H2BA,LINGO4,NBPF7,C1orf137,NOTCH2NL,LINC01138,LOC388692,LYSMD1,NBPF9,LOC440600,LOC440602,BCL2L15,NUDT4P1,HIST2H2BF,PGCP1,NBPF18P,FAM72C,ADAMTSL4-AS1,CYMP,LOC643355,LOC643441,LINC00622,PPIAL4G,FAM231D,NBPF13P,PPIAL4D,LOC645166,EMBP1,SRGAP2B,SRGAP2C,PPIAL4A,GPR89A,PPIAL4C,HIST2H3D,FAM72B,MIR554,FAM72D,NBPF8,LINC00623,LOC728989,PPIAL4E,PFN1P2,PDZK1P1,MIR942,LINC01160,LOC100131107,LOC100132057,LOC100132111,NBPF10,FCGR1CP,C2CD4D,AP4B1-AS1,NBPF20,HYDIN2,LINC00624,MIR320B1,MIR4256,MIR4257,LOC100505824,FAM212B-AS1,SLC16A1-AS1,LOC100507670,TNFAIP8L2-SCNM1,MIR548AC,MIR5087,SRGAP2-AS1,KCND3-AS1,FALEC,HAO2-IT1,KCND3-IT1,LOC100996251,LOC100996263,LINC01356,SRGAP2D,LAMTOR5-AS1,RNVU1-8,LOC101927429,LOC101927468,GBAT2,LOC101928718,HIPK1-AS1,LOC101928977,LOC101928979,LOC101928995,LOC101929023,LOC101929099,LOC101929147,NBPF25P,LOC101929798,RNVU1-4,RNVU1-14,RNVU1-15,RNVU1-20,RNVU1-17,RNVU1-3,RNVU1-1,RNVU1-6,RNVU1-19,MIR6878,MIR6736,MIR6077,CH17-408M7.1,LOC103091866,LINC01525,WARS2-IT1,LOC105371433,LOC105378933 |
| 1p36.11 | 2.5423E-07 | chr1:1-248956422 | hsa-mir-664,hsa-mir-3119-1,hsa-mir-551a,ABCA4,ABL2,ACADM,ACTA1,ACTN2,ADAR,ADORA1,ADORA3,PARP1,ADSS,AGL,AGT,AK2,AK4,ALDH9A1,ALPL,ALX3,AMPD1,AMPD2,AMY1A,AMY1B,AMY1C,AMY2A,AMY2B,APCS,APOA2,FASLG,ARF1,RHOC,ARNT,ASTN1,SERPINC1,ATF3,RERE,ATP1A1,ATP1A2,ATP1A4,ATP1B1,ATP2B4,ATP5F1,ATP6V0B,AVPR1B,ADGRB2,BCL9,BGLAP,BMP8B,BRDT,C1QA,C1QB,C1QC,C4BPA,C4BPB,C8A,C8B,CA6,CACNA1E,CACNA1S,CAPN2,CAPZA1,CAPZB,CASP9,CASQ1,CASQ2,RUNX3,CD1A,CD1B,CD1C,CD1D,CD1E,CD2,CD247,CD5L,TNFRSF8,CD34,CD48,CD53,CD58,CDA,CDK11B,CDC20,CDC42,CDKN2C,CD52,CENPF,RCC1,CHI3L1,CHI3L2,CHIT1,CHML,LYST,CHRM3,CHRNB2,CKS1B,CLCA1,CLCN6,CLCNKA,CLCNKB,CLK2,PLK3,CNN3,CNR2,COL8A2,COL9A2,COL11A1,COL16A1,COPA,CORT,CPT2,CR1,CR1L,CR2,CRABP2,CRP,CRYZ,CSF1,CSF3R,CSRP1,CTBS,CTH,CTPS1,CTSE,CTSK,CTSS,CYP2J2,CYP4A11,CYP4B1,DAB1,CD55,DBT,GADD45A,DDOST,DHX9,DFFA,DFFB,DHCR24,DIO1,DLSTP1,DPH2,DPT,DPYD,DR1,DRD5P2,DVL1,E2F2,ECE1,ECM1,S1PR1,EDN2,PHC2,EIF2D,EFNA1,EFNA3,EFNA4,CELSR2,MEGF6,EPHA2,ELAVL4,ELF3,ELK4,ENO1,ENSA,EPB41,EPHA8,EPHB2,EPHX1,EPRS,EPS15,ESRRG,ETV3,EXTL1,EXTL2,EYA3,F3,F5,F13B,FAAH,FABP3,FCER1A,FCER1G,FCGR1A,FCGR1B,FCGR2A,FCGR2B,FCGR3A,FCGR3B,FDPS,FGR,FH,FHL3,FOXE3,FOXD2,FLG,FMO1,FMO2,FMO3,FMO4,FMO5,FMOD,MTOR,NR5A2,FUCA1,ACKR1,IFI6,GABRD,GALE,GALNT2,GBA,GBAP1,GBP1,GBP2,GBP3,GFI1,GJA4,GJA5,GJA8,GJB3,GJB5,GCLM,GLUL,GNAI3,GNAT2,GNB1,GNG4,GNG5,SFN,GPR3,GPR25,GPX7,GRIK3,GSTM1,GSTM2,GSTM3,GSTM4,GSTM5,GTF2B,GUCA2A,GUCA2B,GUK1,H3F3A,HCRTR1,HDAC1,HDGF,CFH,CFHR1,CFHR2,ZBTB48,MR1,HLX,HMGN2,HMGCL,HMGCS2,HNRNPU,HPCA,HSD3B1,HSD3B2,HSD11B1,HSPA6,HSPA7,IGSF3,HSPG2,HTR1D,HTR6,ID3,IFI16,CYR61,IL6R,IL10,IL12RB2,TNFRSF9,ILF2,INPP5B,INSRR,IPP,IRF6,ITPKB,IVL,JAK1,JUN,KCNA2,KCNA3,KCNA10,KCNC4,KCND3,KCNH1,KCNJ9,KCNJ10,KCNK1,KCNK2,KCNN3,KISS1,LAD1,LAMB3,LAMC1,LAMC2,STMN1,LBR,LCK,LEPR,LGALS8,LMNA,LMX1A,LOR,LY9,TACSTD2,MAGOH,MARK1,MATN1,MCL1,CD46,SMCP,MDM4,MEF2D,MFAP2,MGST3,MNDA,MOV10,MPL,MPZ,MSH4,MTF1,MTHFR,MTR,MTX1,MUC1,MUTYH,MYBPH,MYCL,MYOC,MYOG,PPP1R12B,NASP,NBL1,NCF2,NDUFS2,NDUFS5,NEK2,NFIA,NFYC,NGF,NHLH1,NHLH2,NID1,NIT1,NOTCH2,NPPA,NPPB,NPR1,NRAS,NRDC,YBX1,NTRK1,ROR1,DDR2,NVL,OPRD1,ORC1,OVGP1,PAFAH2,PRDX1,PAX7,PBX1,CDK18,PDC,PDE4B,PDZK1,PEX10,PEX14,PFDN2,PFKFB2,PGD,PGM1,PIGC,PIGR,PIK3C2B,PIK3CD,PI4KB,PIN1P1,PKLR,PKP1,PLA2G2A,PLA2G4A,PLA2G5,PLOD1,PLXNA2,EXOSC10,PRRX1,POU2F1,POU3F1,PPOX,PPP1R8,PPP2R5A,PPT1,PRCC,PRELP,PRKAA2,PRKAB2,PRKACB,PKN2,PRKCZ,PROX1,PSEN2,PSMA5,PSMB2,PSMB4,PSMD4,PTAFR,PTGER3,PTGFR,PTGFRN,PTGS2,QSOX1,PTPN7,PTPN14,PTPRC,PTPRF,PEX19,ABCD3,RAB3B,RAB4A,RAB13,RABGGTB,RABIF,RAP1A,RAP1GAP,RBBP4,RBBP5,REN,RFX5,RGS1,RGS2,RGS4,RGS7,RGS13,RGS16,RHCE,RHD,RIT1,RLF,RNASEL,RNF2,RNPEP,RNU1-4,SNORA73A,SNORD21,RORC,RPA2,RPE65,RPL5,RPL11,RPL22,RPS6KA1,RPS8,RPS27,RSC1A1,RXRG,RYR2,S100A1,SORT1,S100A2,S100A3,S100A4,S100A5,S100A6,S100A7,S100A8,S100A9,S100A10,S100A11,S100A12,S100A13,SARS,SCNN1D,SCP2,XCL1,SDHB,SDHC,SELE,SELL,SELP,SFPQ,SRSF4,SHC1,ST3GAL3,STIL,SKI,SLAMF1,SLC1A7,SLC2A1,SLC2A5,SLC6A9,SLC9A1,SLC16A1,SNRPE,SOAT1,UAP1,SPRR1A,SPRR1B,SPRR2A,SPRR2B,SPRR2C,SPRR2D,SPRR2E,SPRR2F,SPRR2G,SPRR3,SPTA1,SRM,SRP9,TROVE2,SSR2,AURKAPS1,STXBP3,XCL2,SYCP1,TAF12,TAF13,TAL1,TARBP1,CNTN2,TBCE,TBX15,TCEA3,TCEB3,VPS72,TGFB2,LEFTY2,TGFBR3,THBS3,TCHH,TIE1,TLR5,GPR137B,TNFRSF1B,TNNI1,TNNT2,TNR,TOP1P1,TP53BP2,TP73,TPM3,TPR,TRAF5,CCT3,TSHB,TSNAX,TTC4,TUFT1,TNFSF4,TNFRSF4,UCK2,UQCRH,UROD,USF1,USP1,USH2A,VCAM1,WNT2B,WNT9A,ZSCAN20,ZNF124,ZBTB17,SLC30A1,SLC30A2,LUZP1,PRDM2,DNALI1,LRP8,LAPTM5,BSND,CSDE1,EVI5,DAP3,NPHS2,BTG2,PTP4A2,ARID1A,HIST3H3,CDC7,HIST2H2AA3,HIST2H2AC,HIST2H2BE,HIST2H4A,PIP5K1A,TAGLN2,BCAR3,ANXA9,SNHG3,NR0B2,RAD54L,GNPAT,DYRK3,TTF2,CDC42BPA,RGS5,PPFIA4,PIK3R3,MMP23B,MMP23A,KCNAB2,ITGA10,LMO4,FCN3,BLZF1,CDC14A,DEGS1,KMO,YARS,MKNK1,AKR7A2,PLPP3,RTCA,PTCH2,ALDH4A1,EIF3I,EIF4G3,VAMP4,PEA15,B4GALT3,B4GALT2,B3GALT2,TNFRSF25,ADAM15,PABPC4,TNFRSF14,TNFRSF18,FPGT,PEX11B,CREG1,CD84,PER3,FUBP1,EIF2B3,BCL10,RAB29,SELENBP1,TNF |

| | | | |
|---|---|---|---|
| | | | SF18,TAF1A,MPZL1,SH2D2A,ARTN,MAP3K6,ANGPTL1,DIRAS3,TBX19,FCGR2C,SLC16A4,PRPF3,KCNQ4,EXO1,ARHGEF2,DEDD,ZMYM4,ZMYM6,XPR1,FCMR,DHRS3,MAPKAPK2,GPR37L1,GPR52,SRSF11,VAMP3,ZFYVE9,CD101,SEP15,ZRANB2,SNRNP40,ARHGAP29,AIM2,GGPS1,RASAL2,THEMIS2,ADAMTS4,TMEM59,SEC22B,CHD1L,H6PD,SOX13,PRDX6,CLCA3P,CLCA2,ISG15,IKBKE,PLCH2,HS2ST1,PDE4DIP,IPO13,SDC3,SLC25A44,KIAA0040,KDM4A,CROCC,PUM1,NOS1AP,TMEM63A,CEP104,RIMS3,TOMM20,EFCAB14,URB2,ARHGEF11,DNAJC6,CEP350,CEP170,LRIG2,SETDB1,ZC3H11A,SMG7,PLPPR4,UBAP2L,SV2A,KLHL21,SLC35E2,DENND4B,RABGAP1L,TMCC2,FAM20B,ZBTB40,LPGAT1,MFN2,KIF14,RBM8A,THRAP3,NR1I3,AKT3,INSL5,PIGK,SCAMP3,PTPRU,ARPC5,TSPAN2,TSPAN1,CELA3A,PDZK1IP1,WASF2,PATJ,PRG4,ANGPTL7,GPA33,STX6,HNRNPR,SRRM1,CNKSR1,SF3B4,ZMPSTE24,UBE4B,BCAS2,AKR1A1,WARS2,BPNT1,CEPT1,PIAS3,TESK2,TIMM17A,LRRN2,PPIE,VAV3,HAX1,MAD2L2,PPIH,ZBTB18,C1orf61,CAP1,LRRC41,SEMA6C,LAMTOR5,SLC19A2,IFI44,POLR3C,IVNS1ABP,TXNIP,PDPN,LEFTY1,SPHAR,PMVK,KHDRBS1,GMEB1,FAM189B,AP4B1,NUDC,PHTF1,MASP2,CAPN9,NES,KDM5B,AHCYL1,SRSF10,SDCCAG8,CFHR4,CFHR3,WDR3,OCLM,JTB,MTMR11,MAN1A2,UTS2,SF3A3,MLLT11,IFI44L,EBNA1BP2,SLC27A3,KIF2C,IL24,TDRKH,RER1,DNAJB4,ADAM30,RCAN3,FAF1,CD160,GLMN,HHLA3,CELF3,MST1P2,DDX20,DUSP10,MST1L,PADI2,PMF1,DUSP12,VPS45,LYPLA2,PARK7,CTRC,ACOT7,COG2,CLCA4,MTF2,DNAJC8,NTNG1,PLEKHA6,CLSTN1,FOXJ3,KIAA0907,WDR47,KIFAP3,ATF6,SCMH1,AKR7A3,TTC39A,SPEN,KDM1A,RBM34,USP33,WDTC1,KIF21B,NMNAT2,EMC1,KIF1B,NFASC,POGZ,COLGALT2,MAST2,NCDN,CLCC1,SLC35D1,RGL1,PLEKHM2,SYT11,PRRC2C,FBXO28,RPRD2,OTUD3,KAZN,CAMTA1,ADGRL2,CAMSAP2,KIAA1107,ZCCHC11,SZT2,DNAJC16,UBR4,USP24,SRGAP2,SMG5,NCSTN,ATP13A2,CRB1,ITGB3BP,GPR161,TARDBP,CELA3B,SLC35A3,ABCB10,ICMT,MACF1,LRRC8B,ZNF281,SNAPIN,LPAR3,PADI4,DDAH1,TMEM50A,OPN3,PHLDA3,RUSC1,CA14,KPNA6,SSBP3,STX12,DSTYK,RAB3GAP2,CFAP45,LMOD1,NBPF14,MPC2,TRIM58,INTS7,OLFML2B,AHCTF1,C1orf43,CLIC4,NSL1,SYF2,RWDD3,PARS2,MMACHC,ZZZ3,ACOT11,CHD5,DNM3,TOR1AIP1,CHTOP,SZRD1,LDLRAP1,SERBP1,NOC2L,OR1C1,PTPN22,PHGDH,FBXO2,LCE2B,OR2M4,OR2L2,OR2L1P,OPTC,FBXO6,PLA2G2D,AK5,MYCBP,OR10J1,HEYL,AGO1,OR2T1,RPS6KC1,SNORA73B,SNORD81,SNORD79,SNORD80,SNORA66,SNORD47,SNORD45B,SNORD45A,SNORD44,SNORD55,RNU11,RNU6-1,RNU5F-1,RNU5E-1,RNU5D-1,RNVU1-7,RNU1-3,RNU1-2,RNU1-1,RNF11,USP21,FOXD3,DIEXF,TRAPPC3,TAF5L,CACYBP,HSPB7,CHIA,SLC39A1,DISC2,DISC1,ARHGEF16,AHDC1,RNF115,KLHL20,TEKT2,SMPDL3B,ANGPTL3,LAMTOR2,FLVCR1,UBE2T,LINC00339,CNIH4,SSU72,TMOD4,GNL2,GPSM2,UBIAD1,PYCR2,NME7,ALG6,NENF,PADI1,IL19,CERS2,SLC25A24,PLA2G2E,DNTTIP2,WRAP73,CRNN,G0S2,IL20,SLC45A1,DCAF8,HP1BP3,F11R,TMED5,RRP15,GLRX2,BOLA1,DESI2,CELA2B,ZNF593,ZBTB7B,ZNF691,TXNDC12,TNNI3K,RRNAD1,ADIPOR1,SCCPDH,SH3GLB1,MECR,APH1A,UTP11,TRIM17,KCTD3,SDF4,MRTO4,PLEKHO1,HAO2,ACP6,TMEM69,MRPL37,IER5,HOOK1,SNX7,UCHL5,SUCO,HPCAL4,YTHDF2,GPR89B,HSD17B7,UFC1,DTL,ZCCHC17,TRIM33,METTL13,DPH5,HSPB11,OAZ3,PADI3,CYB5R1,CMPK1,ARID4B,CD244,GPR88,ERRFI1,DPM3,WNT4,YIPF1,FBXO42,MRPS21,TRMT13,TMCO1,ADAMTSL4,CRCT1,RNF186,SPATA6,EGLN1,MXRA8,L1TD1,HES2,MAP10,RSBN1,ZNHIT6,GPN2,LEPROT,FBLIM1,MED18,TRIT1,GIPC2,SWT1,GDAP2,FAM46C,GON4L,GPATCH4,PALMD,FNBP1L,ST7L,PQLC2,CASZ1,LAX1,RHBDL2,DUSP23,ADPRHL2,TRNAU1AP,C1orf27,C1orf109,C1orf56,CPSF3L,C1orf123,C1orf159,MARC2,AURKAIP1,TTC22,MRPL20,AIM1L,SUSD4,KIF26B,TMEM51,RALGPS2,GPATCH2,BSDC1,XKR8,TMEM39B,PRPF38B,HEATR1,CDCA8,LRRC8D,MSTO1,DARS2,ARHGEF10L,PRMT6,RNF220,VPS13D,EVA1B,GOLPH3L,ATAD3A,TMEM57,KLHDC8A,TRIM62,ETNK2,RAVER2,PANK4,KIRREL,TMEM206,YY1AP1,ECHDC2,FGGY,MCOLN3,SLC22A15,YOD1,CAMK2N1,BATF3,SLC30A10,UBE2Q1,RNPC3,ITLN1,ASAP3,POMGNT1,PNRC2,LRRC40,DEPDC1,PIGV,ZNF692,NBPF1,IARS2,MAP7D1,IPO9,NDC1,NECAP2,IQCC,C1orf112,HHAT,DNAJC11,ENAH,NUP133,RCOR3,C1orf106,LRIF1,FAM63A,ADCY10,DCAF6,ASH1L,LENEP,CTTNBP2NL,RCC2,FAM212B,DMAP1,AJAP1,GNG12,SLC50A1,TMEM234,MTFR1L,SERTAD4,KYAT3,ERO1B,FMN2,SLAMF8,CDC42SE1,UBQLN4,TMEM167B,OLFML3,SMYD2,LHX9,OTUD7B,COQ8A,CTNNBIP1,RSRP1,AGTRAP,PITHD1,RAB25,PGLYRP4,ZNF695,RHBG,MAN1C1,SCYL3,GJC2,CAMK1G,NIPAL3,SEPN1,ATP8B2,TP73-AS1,VANGL2,LINC00869,PBXIP1,S100A14,TMIGD3,PLEKHG5,GATAD2B,AMIGO1,LRRC47,ODF2L,CGN,KIAA1324,DISP3,IGSF9,LRRC7,SIPA1L2,KIF17,ZNF687,ZSWIM5,POGK,KIAA1522,HCN3,CACHD1,MIER1,KIAA1614,BRINP2,HES4,CCDC181,GRHL3,SLAMF7,ZP4,CADM3,PTBP2,RHOU,PRUNE1,DNASE2B,DLGAP3,IL22RA1,HIVEP3,KLHL12,LGR6,GPBP1L1,HAPLN2,MIIP,GAS5,PAPPA2,CELA2A,BCAN,GPATCH3,TNN,MRPS14,DMRTB1,DMRTA2,CLSPN,PRDM16,OXCT2,RRAGC,ADGRL4,TINAGL1,P3H1,TFB2M,SEMA4A,TOR3A,RFWD2,GREM2,RGS18,PLA2G2F,MFSD14A,NUCKS1,SMAP2,ACBD3,SMYD3,ATPAF1,MARC1,S100PBP,MEAF6,RBM15,EXO5,CEP85,ARV1,NMNAT1,ELOVL1,AIDA,VWA1,DCLRE1B,MRPS15,MRPL9,PINK1,JMJD4,MARCKSL1,PRAMEF1,PRAMEF2,INTS3,NADK,ZFP69B,COA7,PHACTR4,AUNIP,SCNM1,ERI3,C1orf50,WDR77,C1orf116,LINC00626,CCDC28B,C1orf35,EFHD2,MMEL1,RSG1,FCRL2,DLEU2L,OR4F5,NKAIN1,TTC13,EPS8L3,CDC73,MRPL24,MUL1,PGBD5,TNFAIP8L2,C1orf54,TMEM53,AKIRIN1,HECTD3,BEND5,VTCN1,YRDC,ZYG11B,NOL9,PPCS,LIN28A,SH3D21,TTLL7,SNIP1,C1orf115,HHIPL2,VASH2,AGMAT,WDR78,ZMYM1,LINC00115,ZNF669,RPAP2,ZNF672,MORN1,FAM110D,KIAA0319L,DHDDS,PAQR6,DENND2D,WLS,PCNX2,GPR157,TRIM46,MROH9,RPF1,SIKE1,ZC3H12A,SPSB1,TARS2,WDR26,TRIM45,EDEM3,FLAD1,TRAF3IP3,CPTP,ZNF436,TAS1R2,TAS1R1,SH3BP5L,NPL,GJA9,OR4F16,OR6N2,OR6K2,OR2L5,OR2G3,OR2G2,OR2C3,SYNC,CFHR5,TRIM11,C1orf21,ACTL8,ANKRD13C,NECTIN4,SNX27,ANP32E,SHCBP1L,TRMT1L,TSSK3,CCNL2,NUAK2,VANGL1,ST6GALNAC5,ISG20L2,HYI,FCRL5,FCRL4,SH3BGRL3,DDX59,NUF2,RASSF5,SESN2,ESPN,TAS1R3,ATAD3B,HMCN1,GPR61,MIXL1,STK40,SPRTN,TM2D1,FCAMR,REG4,OBSCN,TMEM222,TEX35,PLEKHN1,HORMAD1,TOMM40L,SYDE2,ZNF644,USP48,ZMYND12,NBPF3,LRRC8C,ZDHHC18,SGIP1,POLR3GL,SLC25A33,TMEM79,NTPCR,EFCAB2,DDI2,ACBD6,LZIC,PROK1,MLK4,EFCAB7,ZBTB37,NT5C1A,LCE3D,TRIM63,LINC00260,PSRC1,FAM167B,LINC00467,FOXD2-AS1,PERM1,CROCCP2,FCRLA,ANKRD36BP1,ZNF496,HPDL,ATP1A1-AS1,AGBL4,MFSD2A,C1orf198,PPP1R15B,ATG4C,MAEL,PRPF38A,SYTL1,IGSF21,LSM10,C1orf94,DISP1,SNHG12,STRIP1,RGS8,SLC45A3,DOCK7,CFAP74,WNT3A,NAV1,SEC16B,AQP10,LHX4,SLAMF9,KIAA2013,THAP3,STPG1,PYGO2,ANGEL2,SPOCD1,ZNF697,IGFN1,NUP210L,BTF3L4,UBXN11,NEXN,CENPL,DUSP27,RCSD1,MEX3A,METTL18,GORAB,C1orf105,TMEM183A,HIST3H2A,PIGM,IGSF8,C1orf158,LEMD1,ZNF670,FBXO44,ATPIF1,SNORD46,SNORD38A,SNORD38B,DNAJA1P5,GLMP,MED8,KTI12,SMIM12,AZIN2,TMEM54,HENMT1,TOE1,NLRP3,ERMAP,PGLYRP3,CSMD2,MYSM1,GNRHR2,CROCCP3,FHAD1,SLAMF6,OSBPL9,SLC26A9,LOC115110,OMA1,RAB42,FCRL1,FCRL3,LRRC42,GBP4,GBP5,FAM46B,FMO9P,COX20,RBP7,TSEN15,FAM129A,SNAP47,ACAP3,TADA1,THEM4,SH2D1B,SSX2IP,UBE2J2,OLFM3,GABPB2,TCHHL1,RPTN,CCDC163,TDRD10,SHE,KDF1,CCSAP,LRRC38,AADACL3,PUSL1,B3GALT6,WDR63,KLHDC9,AXDND1,MAB21L3,IFFO2,SLC44A3,ATXN7L2,C1orf194,LYPLAL1,OR2M5,OR2M3,OR2T12,OR14C36,OR2T34,OR2T10,OR2T4,OR2T11,ATP6V1G3,ASB17,TYW3,ERICH3,LRRIQ3,TPRG1L,FAM213B,MYOM3,DMBX1,OR10J5,TMCO2,ZNF684,TCEANC2,PODN,LRRC39,GJB4,HMGB4,RNF19B,A3GALT2,DCST2,DNAH14,OR2B11,ZNF648,TEDDM1,C1orf122,OSCP1,C1orf216,KLHDC7A,VWA5B1,UBXN10,C1orf87,ARL8A,SYT2,LINC00628,GOLT1A,UHMK1,FCRLB,WDR64,C1orf131,LIX1L,HSD3BP4,SPATA17,EDARADD,KLF17,TMEM125,TSACC,IQGAP3,NAXE,ARHGEF19,MRPL55,HIST3H2BB,DRAM2,PIFO,C1orf162,OR10T2,OR6P1,OR10X1,OR10Z1,OR6K6,OR6N1,TATDN3,S100A16,NEK7,ACTRT2,MIB2,ITLN2,SYT6,C1orf74,CREB3L4,C1orf127,BROX,SAMD11,LOC148413,SAMD13,C1orf52,PHF13,CIART,TMEM56,NBPF4,UBE2U,SLC35F3,LINC00337,LOC148696,LOC148709,PTPRVP,HFE2,ANKRD35,FAM163A,OVAAL,B3GALNT2,MFSD4A,PM20D1,GCSAML,GCSAML-AS1,SLC30A7,CCDC27,ZNF436-AS1,KCNN,MOB3C,GLIS1,NBPF12,LELP1,RC3H1,MGC27382,DCDC2B,ZNF362,LINC01225,DCST1,CNIH3,LINC01341,MANEAL,IL23R,METTL11B,FAM78B,SHISA4,EXOC8,LOC149373,PDIK1L,BNI |

| | | | |
|---|---|---|---|
| | | | PL,CLDN19,CFAP57,C1orf210,CCDC24,BTBD19,CCDC17,LRRC71,C1orf64,RNF187,CHIAP2,PYHIN1,SPATA45,FAM71A,SLC2A7,DENND2C,GBP6,PLPPR5,FNDC7,DENND1B,TDRD5,TOR1AIP2,CALML6,IFNLR1,CYP4Z2P,CITED4,SLC25A3P1,LEXM,SPRR4,KANK4,SASS6,SDE2,CNST,FAM43B,HFM1,PAQR7,TTC24,CCDC185,UBL4B,NUDT4P2,PDIA3P1,OXCT2P1,AGO3,AGO4,LCE4A,ALG14,FAM76A,LINC00466,C1orf168,TMEM201,TMEM61,CYP4Z1,FAAP20,CDCP2,SLC5A9,CC2D1B,NBPF11,NUDT17,FLJ23867,TXLNA,TCTEX1D1,PLD5,C1orf100,SPAG17,SLFNL1,KRTCAP2,CRTC2,TMEM51-AS1,IBA57,HIPK1,SLC44A5,ATAD3C,AKR7L,AKR7A2P1,TMEM9,FNDC5,EPHX4,LINC01342,TTLL10,AKNAD1,SLC41A1,OR2T6,LCE5A,TMCO4,MCOLN2,COL24A1,LOC255654,PCSK9,SYT14,ST6GALNAC3,C1orf101,ZNF683,ARHGAP30,CFAP126,NEGR1,ASPM,MDS2,CYP4X1,MAGI3,TIPRL,NPHP4,BEST4,PGBD2,FAM19A3,RIIAD1,THEM5,C1orf167,OR2L13,SLC9C2,OR14A16,CYP4A22,C1orf185,LINC01226,NBPF15,LINC00303,LEMD1-AS1,LOC284578,LOC284581,FAM41C,FAM102B,SYPL2,CYB561D1,ANKRD34A,RUSC1-AS1,LOC284632,LOC284648,SMG7-AS1,RSPO1,EPHA10,LOC284661,C1orf204,C1orf111,LINC01142,ZNF326,BTBD8,RIMKLA,SLC25A34,ESPNP,LIN9,VN1R5,HIST2H2AB,PPM1J,HIST2H3A,HIST2H2BC,HIST2H2BA,FAM151A,S100A7A,RAB7B,LINGO4,FLG-AS1,RXFP4,ANKRD45,LINC01343,C1orf174,KLHL17,TMEM240,TMEM52,BRINP3,MTMR9LP,ZBTB8OS,TFAP2E,ZNF678,PRSS38,LINC01141,CCDC190,LINC01140,LOC339529,LINC01139,LOC339539,C1orf228,ZFP69,RD3,LOC343052,AADACL4,PRAMEF5,HNRNPCL1,PRAMEF10,CCDC18,OR6F1,OR2W3,OR2T8,OR2T3,MYBPHL,OR10R2,FCRL6,KCNT2,BARHL2,NBPF7,TCTEX1D4,OR2T29,SERINC2,FAM159A,FAM131C,LCE1A,LCE1B,LCE1C,LCE1D,LCE1E,LCE1F,LCE2A,LCE2C,LCE2D,LCE3A,LCE3B,LCE3C,LCE3E,PADI6,RGSL1,BMP8A,IRF2BP2,ZBTB41,CYCSP52,DRAXIN,SPATA21,SVBP,TEX38,MROH7,MIGA1,NEXN-AS1,PEAR1,SFT2D2,MIA3,STUM,FAM89A,AGRN,RPS10P7,CENPS,CATSPER4,NSUN4,GPR153,ILDR2,FAM132A,HES5,SMIM1,RNF207,TMEM82,TRNP1,CD164L2,TRABD2B,LDLRAD1,GBP7,C1orf146,FAM69A,SLC6A17,C1orf137,NOTCH2NL,LINC01138,LOC388692,LYSMD1,HRNR,FLG2,C1orf189,FMO6P,LINC00272,C1orf53,TMEM81,CAPN8,COA6,C1orf229,OR2M1P,HES3,PRAMEF12,PRAMEF8,PRAMEF18,PRAMEF17,PLA2G2C,SKINT1L,UOX,FRRS1,VHLL,OR10K2,OR10K1,OR6Y1,OR6K3,VSIG8,OR11L1,OR2L8,OR2AK2,OR2L3,OR2M2,OR2T33,OR2M7,OR2G6,TMEM200B,FAM87B,PRAMEF4,PRAMEF13,SH2D5,NCMAP,LINC01144,C1orf141,GBP1P1,FLJ27354,MIR137HG,C1orf226,LOC400794,C1orf220,LINC01344,C1orf140,NBPF9,FAM177B,RNF223,LDLRAD2,ZNF847P,OR2T2,OR2T5,OR14I1,CSMD2-AS1,OR2T27,OR2T35,APOBEC4,MIR101-1,MIR137,MIR181B1,MIR186,MIR194-1,MIR197,MIR199A2,MIR200A,MIR200B,MIR205,MIR181A1,MIR214,MIR215,MIR29B2,MIR29C,MIR30C1,MIR30E,MIR34A,MIR9-1,MGC34796,RGS21,LHX8,LINC01555,LINC00982,PRAMEF11,PRAMEF6,HNRNPCL2,UQCRHL,MINOS1,SLC2A1-AS1,FAM183A,ZYG11A,FLJ31662,LOC440600,LOC440602,BCL2L15,NUDT4P1,HIST2H2BF,ETV3L,LRRC52,LOC440700,LOC440704,C1orf186,TRIM67,MAP1LC3C,H3F3AP4,GTF2IP20,ANKRD65,PRAMEF7,PRAMEF25,PGCP1,NBPF18P,OR10J3,BECN2,OR2W5,OR13G1,MIR135B,KPRP,LCE6A,SUMO1P3,GEMIN8P4,RBMXL1,LURAP1,PEF1,MIR429,LINC00862,FAM72C,DUSP5P1,SERTAD4-AS1,ADAMTSL4-AS1,C1orf145,PRR9,IBA57-AS1,MIR488,CYB5RL,SNORD74,SSBP3-AS1,RPL31P11,MGAT4EP,MIR205HG,SPATA42,FLVCR1-AS1,CYMP,KIAA0754,LOC643355,LOC643441,LYPLAL1-AS1,LINC01128,TMEM88B,FNDC10,RPS14P3,LINC00622,PPIAL4G,FAM231D,NBPF13P,ACTG1P20,PIK3CD-AS1,NFIA-AS1,PPIAL4D,LOC645166,SNRPD2P2,PRAMEF26,PRAMEF33P,PRAMEF19,PRAMEF20,FAM138A,ASH1L-AS1,POU5F1P4,MT1HL1,RPS7P5,S100A7L2,LACTBL1,LOC646268,LOC646471,LOC646626,LYPD8,SETSIP,EMBP1,SRGAP2B,ASCL5,ACTG1P4,PRAMEF34P,HNRNPCL3,ZBTB8A,NBPF6,LOC653160,SRGAP2C,PPIAL4A,GPR89A,PPIAL4C,HIST2H3D,PRAMEF22,PRAMEF15,WASH7P,FAM72B,PCP4L1,FAM138C,SCARNA3,SCARNA2,SCARNA4,SCARNA1,FAM138D,SNORA14B,SNORA36B,SNORA80E,SNORA44,SNORA55,SNORA61,SNORA77,SNORA59A,SNORA16A,SNORD45C,SNORA16B,SNORD75,SNORD76,SNORD77,SNORD78,SNORD103C,SNORD99,SNORD103A,MIR551A,MIR552,MIR553,MIR554,MIR555,MIR556,MIR557,MIR92B,C1orf195,ZBTB8B,LINC01137,PPIEL,TGFB2-AS1,CCDC30,CDK11A,SLC35E2B,LINC01346,FAM72D,NBPF8,LINC00623,RPS15AP10,LOC728989,FAAHP1,C1orf234,HSD52,PRAMEF14,FAM72A,FAM231A,FAM231C,FLJ37453,LOC729737,OR4F29,LOC729867,LOC729930,LOC729970,LOC729987,LOC730102,LOC730159,LINC01136,PPIAL4E,LINC01347,LINC01348,PFN1P2,MIR765,PDZK1P1,MIR942,MIR190B,MIR760,MIR921,FAM229A,C1orf132,LINC01349,LOC100129046,BTNL10,LOC100129138,MATN1-AS1,LINC01160,C1orf68,MSTO2P,LOC100129534,LOC100129620,LOC100129924,EFCAB14-AS1,LOC100130331,LOC100130417,NFYC-AS1,LINC01135,LOC100131107,TSTD1,RPL21P28,MIR181A1HG,CCDC18-AS1,CADM3-AS1,LOC100132057,LOC100132062,FOXO6,LOC100132111,LOC100132147,NBPF10,FCGR1CP,ZRANB2-AS1,KDM4A-AS1,FAM231B,LOC100133331,LINC01134,LOC100147773,RNA5S1,RNA5S2,RNA5S3,RNA5S4,RNA5S5,RNA5S6,RNA5S7,RNA5S9,RNA5S10,RNA5S11,RNA5S12,RNA5S13,RNA5S14,RNA5S15,RNA5S16,RNA5S17,C2CD4D,DDX11L1,LOC100287497,AP4B1-AS1,LINC00582,TTC34,LGALS8-AS1,GM140,LOC100288069,GS1-279B7.1,NBPF20,HYDIN2,GNG12-AS1,LINC00624,MIR320B1,MIR1182,MIR1537,MIR548N,MIR1256,MIR1231,MIR548F3,MIR1278,MIR1295A,MIR1976,MIR664A,MIR1290,MIR1302-2,MIR1262,RASAL2-AS1,LINC00184,TSNAX-DISC1,MIR320B2,MIR1255B2,MIR761,NPPA-AS1,ZBED6,LOC100422212,MIR1302-9,MIR1302-10,MIR3119-1,MIR4259,MIR3123,MIR3115,MIR3117,MIR3124,MIR3120,MIR4260,MIR4255,MIR3116-1,MIR4253,MIR1302-11,MIR3116-2,MIR3122,MIR4251,MIR1273D,MIR4252,MIR4256,MIR4257,MIR3119-2,MIR4258,MIR4254,MIR3121,MIR3659,MIR3917,MIR3620,MIR3658,MIR3916,MIR3605,MIR3671,MIR3675,LINC01133,LOC100505666,SPATA1,LINC01364,LOC100505795,LOC100505824,PROX1-AS1,LOC100505887,LOC100505918,ENO1-AS1,RTCA-AS1,LOC100506022,LOC100506023,GAS5-AS1,TMEM35B,TAF1A-AS1,FAM212B-AS1,SLC16A1-AS1,ITPKB-IT1,PCAT6,LOC100506730,LOC100506747,LINC01353,LINC01354,LOC100506801,LINC01132,CHRM3-AS2,TCEB3-AS1,LOC100506985,SLFNL1-AS1,MKNK1-AS1,LOC100507564,LOC100507634,LOC100507670,APITD1-CORT,FPGT-TNNI3K,GJA9-MYCBP,MROH7-TTC4,PMF1-BGLAP,LHX4-AS1,TMEM56-RWDD3,MINOS1-NBL1,ZNF670-ZNF695,TNFAIP8L2-SCNM1,MIR4695,MIR4428,MIR1273G,MIR1273F,MIR4420,MIR4419A,MIR3972,MIR4421,MIR4753,MIR4454,MIR4422,MIR4666A,MIR4781,MIR378G,MIR4424,MIR4794,MIR4677,MIR4426,MIR4735,MIR4425,MIR4671,MIR548AC,MIR4654,MIR4427,MIR4684,MIR4711,MIR4689,MIR5096,MIR4418,MIR4632,MIR2682,MIR5095,MIR4742,MIR4423,MIR4417,MIR378F,DNM3OS,RCAN3AS,MIR1295B,MIR5581,MIR5585,MIR5698,MIR5087,MIR5191,MIR5697,MIR5008,MIR5584,MIR5187,NEGR1-IT1,ZRANB2-AS2,LINC00538,PINK1-AS,SRGAP2-AS1,HLX-AS1,DPYD-AS1,DPYD-AS2,MTOR-AS1,VAV3-AS1,IPO9-AS1,KCNC4-AS1,CHRM3-AS1,KCND3-AS1,FALEC,UBE2Q1-AS1,LINC00853,HAO2-IT1,ERI3-IT1,DNM3-IT1,KCND3-IT1,AGBL4- |

| | | | |
|---|---|---|---|
| | | | IT1,LINC00210,LOC100996251,LOC100996263,FOXD3-AS1,LINC01355,NFIA-AS2,LOC100996583,LOC100996630,LOC100996635,LINC01356,SRGAP2D,HNRNPCL4,LAMTOR5-AS1,RNVU1-8,BLACAT1,DAB1-AS1,LINC01358,LOC101926944,LOC101926964,ROR1-AS1,LINC01359,LOC101927139,LOC101927143,LOC101927164,DEPDC1-AS1,LOC101927244,LINC01360,ERICH3-AS1,LOC101927342,LOC101927412,LOC101927429,LOC101927434,LOC101927468,LOC101927478,LINC01361,LOC101927532,LOC101927560,LOC101927587,LOC101927604,LOC101927683,LOC101927765,LOC101927787,LOC101927844,LOC101927851,LOC101927876,GBAT2,PKN2-AS1,LOC101927895,LOC101928009,UBXN10-AS1,LOC101928034,LOC101928043,LOC101928068,LINC01057,LOC101928098,LOC101928118,LOC101928120,LOC101928163,LOC101928177,LOC101928226,LOC101928241,LOC101928270,LOC101928303,LOC101928324,LOC101928370,LOC101928372,LOC101928404,LOC101928436,LOC101928460,LOC101928476,LINC01363,LOC101928565,LOC101928596,LOC101928626,LOC101928650,LOC101928673,LOC101928696,LOC101928718,LOC101928728,LOC101928751,LOC101928778,HIPK1-AS1,LOC101928973,LOC101928977,LOC101928979,LOC101928995,LOC101929023,PIK3CD-AS2,LINC01350,LOC101929099,LINC01351,LOC101929147,LOC101929181,LINC01031,LOC101929224,LOC101929406,LOC101929441,LOC101929464,LOC101929516,LOC101929536,LOC101929541,LOC101929565,LOC101929592,LOC101929626,LOC101929631,LINC01398,LOC101929721,LINC01352,LOC101929771,NBPF25P,LOC101929798,LOC101929901,LOC101929935,PRAMEF27,LOC101930114,RNVU1-4,RNVU1-14,RNVU1-15,RNVU1-20,RNVU1-17,RNVU1-3,RNVU1-1,RNU6-7,RNVU1-6,RNVU1-19,RNU6-8,LINC00970,MIR6068,MIR6079,MIR6084,MIR6723,MIR6726,MIR6727,MIR6728,MIR6731,MIR6732,MIR6733,MIR6735,MIR6737,MIR6738,MIR6740,MIR6742,MIR6878,MIR7641-2,MIR7846,MIR7852,MIR7856,MIR6859-2,MIR6859-3,MIR6736,MIR6769B,MIR6077,MIR6741,MIR6127,MIR6500,MIR6730,MIR6734,MIR6739,MIR6808,MIR6859-1,MIR8083,MIR6729,MIR7156,LOC102606465,LOC102723661,LOC102723727,LOC102723833,LOC102723886,LINC01389,TTC39A-AS1,LOC102724312,LOC102724450,LOC102724539,LOC102724552,CH17-408M7.1,LOC102724571,LOC102724601,LOC102724659,LOC102724661,LOC102724919,LINC01032,LINC01222,LOC103021295,LOC103021296,LOC103091866,TGFB2-OT1,RNU6-2,LINC01461,SPATA17-AS1,PACERR,LINC01036,LINC01221,LINC01397,TXNDC12-AS1,LINC01307,LINC01525,DISC1-IT1,WARS2-IT1,LOC105371433,LOC105371458,LOC105372897,LOC105376736,LOC105376805,LOC105378591,LOC105378614,LOC105378663,LOC105378683,LOC105378732,LOC105378828,LOC105378853,LOC105378933,LOC105748977,LINC01037,MIR34AHG,SNORD128,SCARNA26A,SCARNA26B,SNORA100,SNORA103,SNORA110,POP3,SCARNA18B |
| **Cohort C, Amplification** | | | |
| **cytoband** | **q value** | **wide peak boundaries** | **gene** |
| 19p13.2 | 1.50E-20 | chr19:8908931-8959215 | MUC16 |
| 19q13.41 | 4.93E-20 | chr19:52501084-53491625 | ZNF28,ZNF137P,ZNF701,ZNF83,ZNF415,ZNF665,ZNF702P,ZNF611,ZNF347,ZNF468,ZNF160,ZNF765,ZNF845,BIRC8,ZNF816,ZNF813,ZNF578,ERVV-1,TPM3P9,ZNF600,ZNF320,ZNF525,VN1R2,VN1R4,ZNF677,ZNF808,ZNF888,ZNF761,ZNF818P,ZNF321P,FAM90A27P,ERVV-2,ZNF816-ZNF321P |
| 1q21.3 | 2.93E-18 | chr1:152293714-152309028 | FLG |
| 11p15.4 | 2.41E-17 | chr11:5343526-5398181 | OR51B5,OR51B6,OR51M1 |
| 1q44 | 1.75E-16 | chr1:248180461-248348791 | OR2M4,OR2M3,OR2T12,OR14C36,OR2M2,OR2T33,OR2M7 |
| 14q11.2 | 2.96E-16 | chr14:1-19935931 | OR4K5,OR11H2,OR4K1,OR4N2,OR4K2,POTEG,OR11H12,OR4Q3,OR4M1,DUXAP10,POTEM,LOC642426,LINC01296,LOC100508046,POTEH-AS1,BMS1P17,LOC101929572,BMS1P22 |
| 9p11.2 | 3.14E-14 | chr9:38620231-41991626 | CNTNAP3,FAM201A,CBWD5,ANKRD18A,GLIDR,FAM74A1,ZNF658B,LOC440896,ANKRD20A3,ANKRD20A2,AQP7P3,LOC554249,PGM5P2,CBWD6,SPATA31A1,FOXD4L6,FGF7P3,CNTNAP3B,FRG1HP,GXYLT1P3,FAM95B1,FAM27E2,MIR1299,MIR4477A,LOC101929583,LOC102724238,LOC102724580,LOC102725126,LOC105376064 |
| 11q11 | 3.14E-11 | chr11:49942483-55563862 | TRIM48,OR4C15,OR4A5,OR4A16,OR4A15,OR4C46,OR4C13,OR4C12,OR4C45,TRIM51HP,LOC441601,LOC646813 |
| 1p36.21 | 6.04E-11 | chr1:12794103-13401877 | PRAMEF1,PRAMEF2,PRAMEF5,HNRNPCL1,PRAMEF10,PRAMEF8,PRAMEF18,PRAMEF17,PRAMEF4,PRAMEF13,PRAMEF11,PRAMEF6,HNRNPCL2,PRAMEF7,PRAMEF25,PRAMEF26,PRAMEF33P,PRAMEF19,PRAMEF34P,HNRNPCL3,PRAMEF22,PRAMEF15,PRAMEF14,HNRNPCL4,PRAMEF27 |
| 2q12.2 | 9.27E-11 | chr2:106423362-107900055 | ST6GAL2,RGPD4,RGPD3,RGPD4-AS1,MIR548AU,GACAT1 |
| 7q11.1 | 2.13E-10 | chr7:56095186-64346868 | CHCHD2,ZNF479,ZNF679,DKFZp434L192,NUPR2,LOC401357,ZNF716,ZNF727,GUSBP10,ZNF733P,LOC650226,ZNF736,ZNF735,LOC100130849,LOC100240728,LOC100287704,LOC100287834,MIR4283-2,MIR4283-1,MIR3147,LINC01005,LOC100653233,LOC101928401,LOC102724738,LOC105375297 |
| 7q22.1 | 1.47E-09 | chr7:100992706-101003594 | MUC12 |
| 19p12 | 2.04E-08 | chr19:20029620-29213084 | hsa-mir-1270-1,UQCRFS1,ZNF708,ZNF43,ZNF85,ZNF90,ZNF91,ZNF99,ZNF208,ZNF254,ZNF492,ZNF430,ZNF486,ZNF682,ZNF257,LINC00906,LINC00662,ZNF98,ZNF738,ZNF714,ZNF681,ZNF676,ZNF100,ZNF431,ZNF675,ZNF626,ZNF493,ZNF429,LOC374890,ZNF728,RPSAP58,LINC00664,GOLGA2P9,ZNF724,LOC641367,ZNF826P,ZNF726,HAVCR1P1,LINC01233,ZNF730,ZNF737,IPO5P1,ZNF729,MIR1270,LOC100420587,LINC01532,LOC100996349,LOC101927151,LOC101929124,LOC101929144,LOC101929164,LOC102724908,LOC102724958,LINC01224 |
| 9q34.3 | 2.06E-08 | chr9:136663584-137040667 | ABCA2,C8G,FUT7,PTGDS,TRAF2,EDF1,CLIC3,AGPAT2,PHPT1,EGFL7,FBXW5,RABL6,NPDC1,CCDC183,SNHG7,TMEM141,LCN8,FAM69B,MAMDC4,LCN6,LCN12,C9orf142,LCN15,C9orf172,LCNL1,C9orf139,MIR126,LCN10,SNORA17A,SNORA17B,LOC100128593,CCDC183-AS1,MIR4292,MIR4479,MIR6722 |
| 11q24.2 | 1.49E-07 | chr11:124185931-124310504 | OR8G2,OR8G1,OR8G5,OR8D1 |
| 16p13.3 | 1.21E-06 | chr16:2070929-2119351 | PKD1,TSC2,MIR1225,MIR6511B1,MIR6511B2,LOC105371049 |
| 20q13.33 | 3.25E-06 | chr20:63494947-63693222 | EEF1A2,PTK6,SRMS,GMEB2,STMN3,RTEL1,FNDC11,PPDPF,HELZ2,LOC100505771,RTEL1-TNFRSF6B |
| 10q11.23 | 3.96E-06 | chr10:49906172-50250025 | PABPC3,AMER2 |
| 13q12.13 | 3.96E-06 | chr13:25097161-25170254 | PARG,ASAH2,FAM21A,AGAP6,FAM21EP,TIMM23B |
| 20p13 | 3.96E-06 | chr20:1570743-1636469 | SIRPB1,SIRPG,SIRPG-AS1 |
| 9p24.3 | 2.52E-05 | chr9:1-179024 | FOXD4,CBWD1,FAM138A,FAM138C,WASH1,DDX11L5,MIR1302-2,MIR1302-9,MIR1302-10,MIR1302-11,PGM5P3-AS1 |
| 2q21.1 | 3.23E-05 | chr2:130349834-130666344 | PTPN18,CFC1,TISP43,LOC646743,CYP4F62P,POTEI,CFC1B,POTEJ,FAR2P2 |
| 17p11.2 | 5.03E-05 | chr17:16690246-16932830 | CCDC144A,USP32P1,KRT16P2,FAM106CP |
| 5p15.33 | 5.12E-05 | chr5:209256-265714 | SDHA,CCDC127 |
| 11q13.3 | 0.00024018 | chr11:68980039-70482927 | CCND1,CTTN,FGF3,FGF4,PPFIA1,FADD,FGF19,SHANK2,MYEOV,ANO1,MRGPRD,MRGPRF,TPCN2,ORAOV1,LOC338694,ANO1-AS2,MIR548K,MIR3164,MRGPRF-AS1,LINC01488,LOC101928443,LOC102724265 |
| 4q13.3 | 0.00030917 | chr4:69195776-69736775 | UGT2B4,UGT2B11,UGT2A1,SULT1B1,UGT2B28,UGT2A2,LOC105377267 |

| | | | |
|---|---|---|---|
| 8q24.3 | 0.00031793 | chr8:143868781-145138636 | *CYC1,GPT,GRINA,HSF1,TONSL,PLEC,RPL8,ZNF7,ZNF16,DGAT1,GPAA1,FOXH1,RECQL4,LRRC14,BOP1,FBXL6,OPLAH,COMMD5,CPSF1,CYHR1,VPS28,HGH1,EXOSC4,SLC39A4,ZNF250,C8orf33,SLC52A2,ARHGAP39,ZNF34,SHARPIN,EPPK1,SCRT1,MAF1,PARP10,PPP1R16A,ZNF251,KIFC2,MFSD3,ADCK5,ZNF252P,TMED10P1,ZNF252P-AS1,ZNF517,WDR97,TMEM249,SPATC1,C8orf82,LRRC24,SCX,MIR661,MROH1,MIR939,TONSL-AS1,MIR1234,LOC101928902,MIR6846,MIR6847,MIR6848,MIR7112,MIR6850,MIR6893,MIR6849* |
| 1q12 | 0.00046844 | chr1:117392320-145883191 | *DRD5P2,FCGR1B,HMGCS2,HSD3B1,HSD3B2,NOTCH2,PDZK1,RNU1-4,TBX15,SEC22B,WARS2,PIAS3,POLR3C,WDR3,MAN1A2,ADAM30,CD160,PHGDH,RNU1-3,RNU1-2,RNU1-1,RNF115,HAO2,GDAP2,FAM46C,REG4,ZNF697,HSD3BP4,ANKRD35,NUDT17,SPAG17,NBPF15,HIST2H2BA,NBPF7,LOC388692,FAM72C,LINC00622,FAM231D,PPIAL4D,LOC645166,EMBP1,SRGAP2B,SRGAP2C,PPIAL4A,GPR89A,FAM72B,FAM72D,NBPF8,LINC00623,PPIAL4E,PFN1P2,FCGR1CP,NBPF20,SRGAP2-AS1,HAO2-IT1,LOC100996263,SRGAP2D,LOC101927429,LOC101929147,NBPF25P,LOC101929798,RNVU1-4,RNVU1-14,RNVU1-15,RNVU1-20,RNVU1-17,RNVU1-19,MIR6736,LOC103091866,WARS2-IT1,LOC105378933* |
| 6p22.2 | 0.0007658 | chr6:26370430-26461764 | *BTN3A3,BTN2A2,BTN3A2,BTN3A1,BTN2A1,BTN2A3P* |
| 22q11.1 | 0.00097569 | chr22:1-17108849 | *IL17RA,ANKRD62P1-PARP4P3,POTEH,HSFY1P1,OR11H1,GAB4,CCT8L2,XKR3,TPTEP1,DUXAP8,CECR7,POTEH-AS1,BMS1P17,LOC101929350,LOC102723769,LOC102723780,LINC01297,BMS1P22* |
| 5q31.3 | 0.0010009 | chr5:140857912-140876752 | *AHSG,BCL6,CRYGS,DGKG,EHHADH,EIF4A2,EPHB3,ETV5,HRG,KNG1,LPP,MASP1,RFC4,SNORA63,TRA2B,ST6GAL1,SST,TP63,MAP3K13,ADIPOQ,IGF2BP2,VPS8,FETUB,DNAJB11,TBCCD1,P3H2,SENP2,RTP4,MAGEF1,TMEM41A,RPL39L,RTP1,LIPH,LOC253573,C3orf70,TPRG1,EHHADH-AS1,LPP-AS2,NMRAL1P1,RTP2,FLJ42393,MIR28,SNORD2,SNORA4,IGF2BP2-AS1,SNORA81,MIR944,LOC100131635,MIR1248,MIR5588,MIR548AQ,LPP-AS1,TPRG1-AS2,TPRG1-AS1,ADIPOQ-AS1,LOC101928992,LOC101929106,LOC101929130,LOC102724699,LOC105374266* |
| 17q21.2 | 0.0010009 | chr17:41140480-41388654 | *KRT33A,KRT33B,KRT34,KRTAP9-9,KRTAP4-6,KRTAP9-2,KRTAP9-3,KRTAP9-8,KRTAP17-1,KRTAP4-4,KRTAP9-4,KRTAP4-1,KRTAP4-5,KRTAP4-3,KRTAP4-2,KRTAP9-1,KRTAP9-7,KRTAP16-1,KRTAP9-6,KRTAP29-1* |
| 3q28 | 0.0010009 | chr3:184567453-189984497 | *PCDHA9,PCDHA12,PCDHA11,PCDHA10,PCDHA8,PCDHA7,PCDHA6,PCDHA5,PCDHA4,PCDHA3,PCDHA2,PCDHA1* |
| 3q11.2 | 0.0012314 | chr3:98169022-98479746 | *OR5K1,OR5H6,OR5H2,OR5H15,OR5K3,OR5K4* |
| 16q24.3 | 0.0018308 | chr16:88716882-88745888 | *PIEZO1,LOC339059,LOC100289580* |
| 2p11.2 | 0.0021736 | chr2:86842087-88037800 | *CD8B,PLGLB2,PLGLB1,KRCC1,LINC00152,LOC285074,RGPD1,RGPD2,ANAPC1P1,MIR4435-2,MIR4771-1,MIR4435-1* |
| 4p16.3 | 0.0026708 | chr4:262360-381704 | *ZNF141,ZNF732,MIR571* |
| 12q13.13 | 0.003091 | chr12:52452144-52514688 | *KRT5,KRT6A,KRT6C* |
| 6q26 | 0.0084126 | chr6:160438056-160712480 | *BCR,IGLL1,ZDHHC8P1,LOC388882,CES5AP1,LOC101929374,PCAT14* |
| 22q11.23 | 0.0084126 | chr22:23287941-23573557 | *CSNK2A1,FKBP1A,SOX12,TCF15,PSMF1,SNPH,RBCK1,SDCBP2,ANGPT4,TMEM74B,NSFL1C,TRIB3,NRSN2,FAM110A,ZCCHC3,SCRT2,SLC52A3,TBC1D20,C20orf96,SRXN1,SIRPB2,RSPO4,C20orf202,RAD21L1,NRSN2-AS1,SDCBP2-AS1,FKBP1A-SDCBP2,MIR6869,LOC105372493* |
| 20p13 | 0.0084126 | chr20:288961-1478564 | *LPA,PLG,SLC22A3,LPAL2* |
| 17q21.31 | 0.013533 | chr17:45973707-46931102 | *MAPT,NSF,WNT3,WNT9B,GOSR2,LRRC37A,ARL17A,STH,KANSL1,LRRC37A2,KANSL1-AS1,NSFP1,ARL17B* |
| 9p13.3 | 0.020773 | chr9:33101418-33558125 | *AQP3,AQP7,BAG1,B4GALT1,NFX1,SPINK4,CHMP5,NOL6,SUGT1P1,ANKRD18B,B4GALT1-AS1,MIR6851* |
| 15q13.2 | 0.028502 | chr15:27982674-32974165 | *APBA2,CHRNA7,TRPM1,OCA2,SCG5,TJP1,HERC2,ARHGAP11A,FAN1,FAM189A1,DKFZP434L187,GREM1,KLF13,MTMR10,NSMCE3,CHRFAM7A,ULK4P3,ULK4P1,ARHGAP11B,OTUD7A,LOC283710,GOLGA8G,GOLGA8IP,FMN1,HERC2P10,MIR211,HERC2P9,WHAMMP2,GOLGA8N,PDCD6IPP2,GOLGA8J,GOLGA8T,GOLGA8K,GOLGA8M,GOLGA8O,GOLGA6L7P,GOLGA8H,LOC100130111,LOC100131315,GOLGA8F,WHAMMP1,LOC100288637,LOC100289656,MIR1268A,MIR4509-1,MIR4509-2,LOC100996255,GOLGA8R,LOC101928042,LOC102725022,LOC105370757* |
| 12p13.2 | 0.044452 | chr12:10399540-10460401 | *KLRC1,KLRC2,KLRC3,KLRC4,KLRC4-KLRK1* |
| 7p22.3 | 0.046725 | chr7:1-936851 | *PDGFA,PRKAR1B,ADAP1,SUN1,GET4,DNAAF5,FAM20C,HRAT92,LOC442497,WI2-2373I1.2,LOC100507642,LOC101926963,LOC101927000,LOC102723672,LOC105375115* |
| **Cohort A, Deletions** | | | |
| **cytoband** | **q value** | **wide peak boundaries** | **gene** |
| 5q31.3 | 6.04E-13 | chr5:140787350-141404487 | *TAF7,PCDHGB4,PCDHGA8,PCDHA9,PCDHB5,PCDHB1,PCDHB18P,PCDHB17P,PCDHGB5,PCDHGB3,PCDHGB2,PCDHGB1,PCDHGA7,PCDHGA6,PCDHGA5,PCDHGA4,PCDHGA3,PCDHGA2,PCDHGA1,PCDHB15,PCDHB14,PCDHB13,PCDHB12,PCDHB11,PCDHB10,PCDHB9,PCDHB8,PCDHB7,PCDHB6,PCDHB4,PCDHB3,PCDHB2,PCDHAC2,PCDHAC1,PCDHA13,PCDHA12,PCDHA11,PCDHA10,PCDHA8,PCDHA7,PCDHA6,PCDHA5,PCDHA4,PCDHA3,PCDHA2,PCDHB16,SLC25A2,PCDHB19P,LOC101926905* |
| 19p12 | 3.73E-06 | chr19:19723177-24167696 | *hsa-mir-1270-1,ZNF708,ZNF43,ZNF85,ZNF90,ZNF91,ZNF99,ZNF208,ZNF254,ZNF253,ZNF492,ZNF430,ZNF93,ZNF486,ZNF682,ZNF257,ZNF98,ZNF738,ZNF714,ZNF681,ZNF676,ZNF100,ZNF431,ZNF675,ZNF626,LINC00663,ZNF493,ZNF429,LOC374890,ZNF728,RPSAP58,LINC00664,ZNF506,GOLGA2P9,ZNF724,LOC641367,ZNF826P,ZNF726,HAVCR1P1,LINC01233,ZNF730,ZNF737,IPO5P1,ZNF729,MIR1270,LOC100996349,LOC101929124,LOC101929144,LOC101929164,LINC01224* |
| 19q13.41 | 1.26E-05 | chr19:52374641-53642009 | *ZNF28,ZNF137P,ZNF331,ZNF701,ZNF83,ZNF415,ZNF665,ZNF702P,ZNF611,ZNF528,ZNF347,ZNF468,ZNF160,ZNF765,ZNF845,BIRC8,ZNF816,ZNF813,ZNF534,ZNF578,ERVV-1,TPM3P9,ZNF600,ZNF320,ZNF525,LOC284379,VN1R2,VN1R4,ZNF677,ZNF808,ZNF888,ZNF761,ZNF818P,ZNF321P,DPRX,FAM90A27P,ERVV-2,ZNF816-ZNF321P,ZNF528-AS1* |
| 19p13.2 | 0.00010841 | chr19:9083235-9102974 | *OR1M1* |
| 1q21.3 | 0.0041167 | chr1:152286652-152761269 | *FLG,LCE2B,CRNN,CRCT1,LCE3D,LCE4A,LCE5A,FLG-AS1,LCE2A,LCE2C,LCE2D,LCE3A,LCE3B,LCE3C,LCE3E,FLG2,C1orf68* |
| 11p15.4 | 0.0040238 | chr11:4390756-6210104 | *HBB,HBBP1,HBD,HBE1,HBG1,HBG2,TRIM22,OR52A1,UBQLN3,TRIM34,TRIM68,MMP26,OR51G1,OR51B4,OR51B2,OR52N1,OR51G2,OR51E2,TRIM5,TRIM6,OR52E2,OR52J3,OR51L1,OR51A7,OR51S1,OR51F2,OR52R1,OR52M1,OR52K2,OR52W1,OR56A4,OR56A1,C11orf40,OR52I2,OR51E1,UBQLNL,OR56B4,OR52B2,OR51F1,OR51B5,OR51V1,OR52L1,OR52B6,OR56B1,OR52K1,OR52I1,OR51D1,OR52A5,OR51B6,OR51M1,OR51Q1,OR51I1,OR51I2,OR52D1,OR52H1,OR52N4,OR52N5,OR52N2,OR52E6,OR52E8,OR52E4,OR56A3,OR56A5,OR51T1,OR51A4,OR51A2,TRIM6-TRIM34,OLFM5P,BGLT3* |
| 1p36.32 | 0.033219 | chr1:1-6571979 | *hsa-mir-551a,CDK11B,DFFB,DVL1,MEGF6,GABRD,GNB1,PEX10,PRKCZ,RPL22,SCNN1D,SKI,TP73,TNFRSF4,MMP23B,MMP23A,KCNAB2,TNFRSF25,TNFRSF14,TNFRSF18,ISG15,PLCH2,CEP104,SLC35E2,RER1,ACOT7,ICMT,CHD5,NOC2L,ARHGEF16,SSU72,WRAP73,SDF4,MXRA8,HES2,CPSF3L,C1orf159,AURKAIP1,MRPL20,ATAD3A,PANK4,AJAP1,TP73-AS1,PLEKHG5,LRRC47,HES4,PRDM16,VWA1,NADK,MMEL1,OR4F5,NOL9,LINC00115,MORN1,CPTP,OR4F16,CCNL2,ESPN,TAS1R3,ATAD3B,PLEKHN1,PERM1,CFAP74,LOC115110,ACAP3,UBE2J2,PUSL1,B3GALT6,TPRG1L,FAM213B,ACTRT2,MIB2,SAMD11,LOC148413,LINC00337,CCDC27,CALML6,FAAP20,ATAD3C,LINC01342,TTLL10,NPHP4,FAM41C,LOC284661,C1orf174,KLHL17,TM* |

| | | | |
|---|---|---|---|
| | | | EM240,TMEM52,AGRN,GPR153,FAM132A,HES5,SMIM1,RNF207,HES3,FAM87B,RNF223,MIR200A,MIR200B,LINC00982,ANKRD65,MIR429,LINC01128,TMEM88B,FNDC10,FAM138A,WASH7P,FAM138C,FAM138D,MIR551A,CDK11A,SLC35E2B,LINC01346,LOC729737,OR4F29,LOC100129534,LOC100130417,LOC100132062,LOC100133331,LINC01134,DDX11L1,TTC34,LOC100288069,MIR1302-2,MIR1302-9,MIR1302-10,MIR1302-11,MIR4251,MIR4252,MIR4689,MIR4417,LOC100996583,LOC101928626,MIR6723,MIR6726,MIR6727,MIR6859-2,MIR6859-3,MIR6808,MIR6859-1,LOC102724312,LOC102724450,LOC105378591 |
| 2q31.2 | 0.033219 | chr2:178513062-178629613 | TTN-AS1 |
| 4p16.3 | 0.048743 | chr4:1-2261097 | ATP5I,CTBP1,DGKQ,FGFR3,GAK,IDUA,LETM1,MYL5,PDE6B,WHSC1,NELFA,ZNF141,SLBP,MAEA,PCGF3,SPON2,TACC3,CPLX1,SLC26A1,FGFRL1,NKX1-1,PIGG,UVSSA,HAUS3,ABCA11P,MFSD7,TMEM175,CTBP1-AS2,TMEM129,ZNF595,FAM53A,ZNF721,ZNF718,CTBP1-AS,CRIPAK,RNF212,NAT8L,POLN,C4orf48,ZNF876P,ZNF732,SCARNA22,MIR571,MIR943,LOC100129917,LOC100130872,TMED11P,MIR4800,LOC105374344 |
| | | **Cohort B, Deletions** | |
| **cytoband** | **q value** | **wide peak boundaries** | **gene** |
| 5q31.3 | 4.33E-44 | chr5:141166694-141247754 | PCDHB18P,PCDHB14,PCDHB13,PCDHB12,PCDHB11,PCDHB10,PCDHB9,PCDHB8,PCDHB7,PCDHB16,PCDHB19P |
| 5p15.1 | 1.10E-18 | chr5:16877686-21463829 | CDH18,BASP1,LOC285696,LOC401177,LOC646241,LOC101929544,LOC102723526 |
| 9q12 | 1.53E-17 | chr9:38620231-68544822 | PGM5,ZNF658,SPATA31A7,CNTNAP3,ANKRD20A1,FAM201A,TMEM252,CBWD5,LOC286297,FOXD4L3,FOXD4L4,AQP7P1,FGF7P6,SPATA31A6,GLIDR,FAM74A1,FAM74A4,ZNF658B,LOC403323,LOC440896,ANKRD20A3,ANKRD20A2,AQP7P3,PTGER4P2-CDK2AP2P2,CBWD3,LOC554249,PGM5-AS1,PGM5P2,FRG1JP,LOC642929,LINC01189,CNTNAP3P2,CBWD6,FLJ43315,SPATA31A1,FOXD4L6,FOXD4L5,FGF7P3,SPATA31A3,SPATA31A5,FAM74A3,CNTNAP3B,LOC728673,ANKRD20A4,FAM27E3,LOC100132249,FRG1HP,GXYLT1P3,FAM27C,FAM95B1,FAM27B,FAM27E2,MIR1299,MIR4477A,FAM74A7,LOC101927827,LOC101928195,LOC101928381,LOC101929583,LOC102723709,LOC102724238,LOC102724580,LOC102725126,LINC01410,LOC103908605,LOC105376064,LOC105379450,XLOC_007697 |
| 7q22.1 | 1.53E-17 | chr7:100893871-100982969 | MUC3A |
| 10q26.3 | 4.16E-12 | chr10:133402336-133797422 | CYP2E1,SYCE1,SPRNP1,FRG2B,SPRN,SCART1,DUX4 |
| 13q34 | 4.19E-12 | chr13:113343949-113430088 | GRTP1-AS1,LOC101928841 |
| 9q34.3 | 1.75E-11 | chr9:136245616-136353414 | GPSM1,DKFZP434A062,CCDC187 |
| 19q13.41 | 1.41E-10 | chr19:51475470-53371767 | hsa-mir-643,SIGLEC6,FPR1,FPR2,FPR3,HAS1,PPP2R1A,ZNF28,ZNF137P,ZNF175,SIGLEC5,ZNF432,ZNF701,ZNF83,ZNF415,ZNF350,ZNF649,ZNF665,ZNF613,ZNF702P,ZNF614,ZNF611,ZNF528,ZNF347,ZNF577,SIGLEC12,ZNF616,ZNF766,ZNF468,ZNF160,ZNF845,BIRC8,ZNF816,SPACA6,ZNF480,ZNF534,ZNF578,ERVV-1,ZNF836,ZNF610,ZNF600,ZNF320,ZNF615,ZNF841,VN1R2,VN1R4,ZNF677,ZNF808,ZNF888,ZNF818P,ZNF321P,ZNF880,MIRLET7E,MIR125A,MIR99B,FAM90A27P,MIR643,CEACAM18,LINC01530,SIGLEC14,ERVV-2,ZNF816-ZNF321P,ZNF350-AS1,ZNF649-AS1,SPACA6P-AS,MIR6801,ZNF528-AS1 |
| 16p13.13 | 1.52E-10 | chr16:11350681-11549537 | LOC101927131 |
| 19p13.3 | 1.85E-09 | chr19:1-524586 | MADCAM1,PLPP2,SHC2,THEG,MIER2,OR4F17,TPGS1,C2CD4C,ODF3L2,WASH5P,LINC01002,FAM138A,FAM138C,MIR1302-2,MIR1302-9,MIR1302-10,MIR1302-11 |
| 11p15.5 | 8.35E-09 | chr11:1196636-1279748 | MUC5B,MIR6744 |
| 2q32.1 | 3.51E-08 | chr2:183093043-186595282 | ZC3H15,ZNF804A,NUP35,FSIP2,MIR548AE1,LOC101927196,LINC01473,LOC105373782 |
| 2q37.3 | 1.38E-07 | chr2:240625825-242193529 | AGXT,KIF1A,BOK,DTYMK,HDLBP,SEPT2,PDCD1,PPP1R7,FARP2,STK25,PASK,ATG4B,SNED1,ANO7,THAP4,GAL3ST2,C2orf54,ING5,NEU4,MTERF4,LOC200772,RTP5,LOC285095,AQP12A,AQP12B,D2HGDH,LOC728323,BOK-AS1,MIR3133,LINC01237,LOC102723927 |
| 21q22.3 | 1.53E-07 | chr21:44551468-44638274 | KRTAP10-4,KRTAP10-6,KRTAP10-7,KRTAP10-9,KRTAP10-5,KRTAP10-8,KRTAP10-3 |
| 16p13.3 | 1.16E-06 | chr16:2068302-2149949 | PKD1,MIR1225,MIR3180-5,MIR4516,MIR6511B1,MIR6511B2,LOC105371049 |
| 1p36.32 | 1.27E-06 | chr1:2205079-6525914 | hsa-mir-551a,DFFB,MEGF6,PEX10,RPL22,SKI,TP73,KCNAB2,TNFRSF25,TNFRSF14,PLCH2,CEP104,RER1,ACOT7,ICMT,CHD5,ARHGEF16,WRAP73,HES2,PANK4,AJAP1,TP73-AS1,PLEKHG5,LRRC47,PRDM16,MMEL1,MORN1,ESPN,LOC115110,TPRG1L,FAM213B,ACTRT2,LINC00337,CCDC27,NPHP4,LOC284661,C1orf174,GPR153,HES5,SMIM1,RNF207,HES3,LINC00982,MIR551A,LINC01346,LOC100129534,LINC01134,TTC34,MIR4251,MIR4252,MIR4689,MIR4417,LOC100996583,LOC102724450 |
| 14q32.33 | 2.44E-06 | chr14:104912717-104992001 | AHNAK2,PLD4 |
| 9p24.3 | 3.53E-06 | chr9:1-121604 | FOXD4,FAM138A,FAM138C,WASH1,DDX11L5,MIR1302-2,MIR1302-9,MIR1302-10,MIR1302-11,PGM5P3-AS1 |
| 7q36.3 | 4.76E-06 | chr7:158704060-159345973 | VIPR2,WDR60,ESYT2,LINC00689 |
| 12q24.33 | 9.21E-06 | chr12:132818787-133275309 | ZNF10,ZNF26,ZNF84,ZNF140,ZNF268,CHFR,ANHX,ZNF605,ZNF891,LOC101928530,LOC101928597 |
| 16p13.3 | 1.26E-05 | chr16:737846-1585453 | CLCN7,SSTR5,TPSAB1,UBE2I,CACNA1H,BAIAP3,TELO2,MSLN,TPSD1,TPSG1,SOX8,GNG13,CHTF18,TPSB2,UNKL,LMF1,TMEM204,GNPTG,RPUSD1,TSR3,SSTR5-AS1,C16orf91,PRR25,C1QTNF8,PTX4,CCDC154,MIR662,LMF1-AS1 |
| 13q12.11 | 2.43E-05 | chr13:1-24767991 | PARP4,ATP12A,FGF9,GJA3,GJB2,MIPEP,SGCG,TUBA3C,ZMYM2,IFT88,ZMYM5,SAP18,GJB6,FAM230C,SACS,LATS2,CRYL1,IL17D,MPHOSPH8,PSPC1,TNFRSF19,XPO4,MRPL57,TPTE2,EEF1AKMT1,SKA3,MICU2,SPATA13,ZDHHC20,ANKRD20A9P,C1QTNF9,LINC00442,TPTE2P6,C1QTNF9B,ANKRD20A19P,C1QTNF9B-AS1,BASP1P1,MIPEPP3,ANKRD26P3,LINC00421,MIR2276,LINC00540,SACS-AS1,LINC00327,MIR4499,LINC00408,LINC00539,LINC00566,LINC00350,LINC00417,LINC00424,SPATA13-AS1,LINC00621,LOC101928697,LINC00367,LINC01046,LINC01072 |
| 16q24.3 | 0.00010178 | chr16:89910188-90338345 | AFG3L1P,GAS8-AS1,GAS8,MC1R,TUBB3,PRDM7,DEF8,DBNDD1,CENPBD1,URAHP,FAM157C |
| 3p22.2 | 0.00012523 | chr3:37176132-38551259 | ACAA1,ACVR2B,GOLGA4,ITGA9,MYD88,PLCD1,SLC22A14,SLC22A13,DLEC1,EXOG,XYLB,OXSR1,CTDSPL,VILL,C3orf35,MIR26A1,ACVR2B-AS1,ITGA9-AS1 |
| 10q11.22 | 0.00024556 | chr10:43407684-48910096 | ALOX5,CTSLP2,GDF2,GDF10,MSMB,NPY4R,MAPK8,RBP3,CXCL12,ZNF22,ZNF32,NCOA4,ZNF239,GPRIN2,HNRNPA3P1,C10orf10,PTPN20,ARHGAP22,OR13A1,SYT15,RASSF4,ZFAND4,ZNF488,AGAP4,FRMPD2,ANTXRL,MARCH8,C10orf25,TMEM72-AS1,ZNF485,FAM21C,LINC00841,ANKRD30BP3,BMS1P5,ZNF32-AS1,ZNF32-AS3,ZNF32-AS2,GLUD1P7,AGAP12P,FAM35BP,LINC00619,FAM35DP,AGAP9,ZNF487,BMS1P6,TMEM72,LINC00842,FAM25C,ANXA8,AGAP7P,ANXA8L1,PARGP1,HNRNPA1P33,FRMPD2B,C10orf142,FAM25BP,FAM25G,RSU1P2,TIMM23,MIR3156-1,LINC00840,ANTXRLP1,CH17-360D5.1,LOC102724264,LOC102724323,LOC102724593,LOC105378292,LOC107001062 |
| 22q11.1 | 0.0024253 | chr22:16988368-17119409 | IL17RA,CECR7 |
| 15q11.2 | 0.0026654 | chr15:1-33155804 | APBA2,NBEAP1,CHRNA7,GABRA5,GABRB3,GABRG3,IPW,TRPM1,NDN,OCA2,SCG5,SNRPN,TJP1,UBE3A,MKRN3,PWAR5,HERC2,SNURF,ARHGAP11A,FAN1,CYFIP1,FAM189A1,NPAP1,DKFZP434L187,GREM1,KLF13,MAGEL2,MTMR10,NSMCE3,ATP10A,NIPA2,CHRFAM7A,ULK4P3,ULK4P1,ARHGAP11B,SNORD107,TUBGCP5,NIPA1,PWAR1,OTUD7A,LOC283683,GOLGA6L2,OR4N4,LOC283710,HERC2P3,GOLGA6L1,GOLGA8G,GOLGA8IP,SNORD108,SNORD109A,SNORD115-1,WHAMMP3,FMN1,SNORD64,PWAR4,PWARSN,LINC01193,IGHV1OR15- |

| | | | |
|---|---|---|---|
| | | | 1,GOLGA8EP,OR4M2,OR4N3P,HERC2P10,HERC2P2,MIR211,NF1P2,GOLGA6L22,HERC2P9,WHAMMP2,LINC00929,GOLGA8N,CHEK2P2,LOC646214,CXADRP2,PDCD6IPP2,IGHV1OR15-3,REREP3,GOLGA8S,GOLGA8J,GOLGA8T,GOLGA8K,GOLGA8M,GOLGA6L6,LOC727924,GOLGA8O,GOLGA6L7P,GOLGA8H,GOLGA8CP,PWRN1,PWRN2,SNORD116-1,SNORD116-2,SNORD116-3,SNORD116-4,SNORD116-5,SNORD116-8,SNORD116-10,SNORD116-11,SNORD116-12,SNORD116-13,SNORD116-14,SNORD116-15,SNORD116-16,SNORD116-17,SNORD116-18,SNORD116-20,SNORD116-21,SNORD116-22,SNORD116-23,SNORD116-24,SNORD116-25,SNORD115-2,SNORD116-26,SNORD116-27,SNORD115-3,SNORD115-4,SNORD115-5,SNORD115-6,SNORD115-7,SNORD115-8,SNORD115-10,SNORD115-11,SNORD115-12,SNORD115-13,SNORD115-14,SNORD115-15,SNORD115-16,SNORD115-17,SNORD115-18,SNORD115-20,SNORD115-21,SNORD115-22,SNORD115-23,SNORD115-25,SNORD115-26,SNORD115-29,SNORD115-30,SNORD115-31,SNORD115-32,SNORD115-33,SNORD115-34,SNORD115-35,SNORD115-36,SNORD115-37,SNORD115-38,SNORD115-39,SNORD115-41,SNORD115-44,SNORD116-28,SNORD116-29,SNORD115-48,SNORD115-24,SNORD115-27,SNORD115-28,SNORD115-45,SNORD115-47,LOC100128714,LOC100130111,LOC100131315,HERC2P7,GOLGA8F,GOLGA8DP,POTEB2,WHAMMP1,LOC100288637,LOC100289656,MIR1268A,MIR3118-3,MIR3118-4,MIR3118-2,MIR4509-1,MIR4509-2,MIR4508,MIR4715,MIR5701-1,SNORD116-30,SNORD115-46,LOC100996255,POTEB,GOLGA8R,LOC101927079,LOC101928042,PWRN3,GABRG3-AS1,POTEB3,LOC102725022,PWRN4,LOC105370757 |
| 15q26.3 | 0.0034128 | chr15:98764955-99145694 | SYNM,PGPEP1L,MIR4714,LUNAR1 |
| 17p11.2 | 0.0040021 | chr17:21252834-27427487 | KCNJ12,MAP2K3,WSB1,FAM27E5,FLJ36000,C17orf51,KCNJ18,MTRNR2L1,MIR4522,LOC105371703 |
| 17p13.1 | 0.0041359 | chr17:10189520-10745332 | MYH1,MYH2,MYH3,MYH4,MYH8,SCO1,MYH13,ADPRM,TMEM220,MYHAS,MAGOH2P |
| 3p13 | 0.0047148 | chr3:69386302-71199125 | MITF,FOXP1,SAMMSON |
| 17q25.3 | 0.0055908 | chr17:80241794-81255491 | NPTX1,AATK,BAIAP2,CEP131,RPTOR,RNF213,CHMP6,TEPSIN,ENDOV,NDUFAF8,AATK-AS1,BAIAP2-AS1,MIR338,MIR657,LOC100294362,MIR1250,MIR3065,MIR4730,LOC101928855,LOC105371925 |
| 12q12 | 0.0063108 | chr12:40361034-47964509 | CNTN1,NELL2,TWF1,VDR,ENDOU,SCAF11,YAF2,RAPGEF3,PDZRN4,PLEKHA8P1,IRAK4,PPHLN1,HDAC7,SLC38A2,SLC38A4,SLC48A1,RPAP3,ADAMTS20,SLC38A1,PUS7L,RACGAP1P,TMEM117,ZCRB1,PCED1B,PRICKLE1,ANO6,ARID2,MUC19,GXYLT1,AMIGO2,LINC00938,DBX2,PCED1B-AS1,LOC100288798,MIR4494,MIR4698,LOC101927038,LOC101927058,MIR7851,LOC105369738,LOC105369739,LOC105369747 |
| 11q25 | 0.0064173 | chr11:134261821-135086622 | B3GAT1,GLB1L2,GLB1L3,LOC283177,LOC100507548 |
| 20q13.33 | 0.0065933 | chr20:62161188-63592933 | CHRNA4,COL9A3,EEF1A2,KCNQ2,LAMA5,NTSR1,PTK6,RPS21,SRMS,OSBPL2,TCFL5,ADRM1,OGFR,DIDO1,HRH3,MTG2,SLCO4A1,YTHDF1,GID8,MRGBP,ARFGAP1,COL20A1,SLC17A9,LOC63930,FNDC11,PPDPF,BIRC7,CABLES2,HELZ2,BHLHE23,NKAIN4,MIR1-1HG,GATA5,RBBP8NL,C20orf166-AS1,MIR1-1,MIR124-3,MIR133A2,HAR1A,HAR1B,SLCO4A1-AS1,LOC100130587,DPH3P1,LINC00029,LINC01056,FLJ16779,MIR4326,MIR3196,MIR4758,LINC00659,OGFR-AS1,LAMA5-AS1 |
| 1q42.11 | 0.0066397 | chr1:222747990-225781330 | CAPN2,LBR,NVL,TLR5,TP53BP2,DEGS1,FBXO28,CNIH4,SUSD4,ENAH,WDR26,DISP1,DNAH14,CNIH3,CCDC185,CAPN8,GTF2IP20,LOC100287497,MIR320B2,MIR4742,LOC101927143,LOC101927164 |
| 2p25.3 | 0.0082197 | chr2:1-10880747 | ACP1,HPCAL1,ID2,ODC1,RPS7,RRM2,SOX11,ADAM17,TPO,TSSC1,PXDN,KLF11,ASAP2,TAF1B,ITGB1BP1,RNF144A,PDIA6,YWHAQ,MYT1L,SH3YL1,GRHL1,TRAPPC12,CPSF3,SNTG2,ADI1,ALLC,KIDINS220,COLEC11,NOL10,RSAD2,CMPK2,MBOAT2,TMEM18,LINC01105,CYS1,ATP6V1C2,RNASEH1,FAM150B,IAH1,LINC00298,LINC00299,LINC01115,C2orf48,RNF144A-AS1,LOC400940,LINC00487,FAM110C,LINC01249,DCDC2C,MYT1L-AS1,SNORA80B,MIR4261,LINC01304,RNASEH1-AS1,LOC100506274,ID2-AS1,LOC101060385,LOC101060391,LINC01250,LINC01247,LOC101929452,LOC101929551,LOC101929567,LOC101929715,LOC101929882,MIR7158,MIR7515,LINC01248,MIR7515HG,LINC01246,NRIR,LOC105373352,LOC105373394 |
| 11q13.4 | 0.0095881 | chr11:69810181-71840509 | DHCR7,CTTN,KRTAP5-9,PPFIA1,FADD,SHANK2,ANO1,NADSYN1,FAM86C1,KRTAP5-8,SHANK2-AS3,DEFB108B,ALG1L9P,KRTAP5-10,FLJ42102,KRTAP5-7,KRTAP5-11,ANO1-AS2,ZNF705E,MIR548K,MIR3664,SHANK2-AS1,LOC101928443,MIR6754 |
| 1q21.3 | 0.011914 | chr1:152220816-152343330 | FLG |
| 1p22.1 | 0.012527 | chr1:72276707-108697726 | ABCA4,ACADM,AGL,AMY1A,AMY1B,AMY1C,AMY2A,AMY2B,BRDT,CLCA1,CNN3,COL11A1,CRYZ,CTBS,DBT,DLSTP1,DPYD,DR1,S1PR1,EXTL2,F3,GBP1,GBP2,GBP3,GFI1,GCLM,GNG5,GTF2B,CYR61,MSH4,PRKACB,PKN2,PTGFR,ABCD3,RABGGTB,SNORD21,RPL5,TGFBR3,VCAM1,EVI5,CDC7,BCAR3,LMO4,CDC14A,RTCA,FPGT,FUBP1,BCL10,SEP15,ARHGAP29,CLCA3P,CLCA2,HS2ST1,PLPPR4,PIGK,VAV3,IFI44,IFI44L,DNAJB4,GLMN,CLCA4,MTF2,NTNG1,USP33,ADGRL2,KIAA1107,SLC35A3,LRRC8B,LPAR3,DDAH1,RWDD3,ZZZ3,AK5,SNORA66,SNORD45B,SNORD45A,SLC25A24,DNTTIP2,TMED5,TNNI3K,SH3GLB1,SNX7,DPH5,GPR88,TRMT13,ZNHIT6,GIPC2,PALMD,FNBP1L,LRRC8D,PRMT6,MCOLN3,RNPC3,KYAT3,ODF2L,PTBP2,DNASE2B,ADGRL4,MFSD14A,TTLL7,RPAP2,RPF1,ST6GALNAC5,SYDE2,ZNF644,LRRC8C,NEXN,DNAJA1P5,HENMT1,GBP4,GBP5,SSX2IP,OLFM3,WDR63,SLC44A3,ASB17,TYW3,ERICH3,LRRIQ3,LRRC39,SAMD13,C1orf52,TMEM56,NBPF4,SLC30A7,MGC27382,GBP6,PLPPR5,SASS6,HFM1,ALG14,SLC44A5,EPHX4,MCOLN2,COL24A1,ST6GALNAC3,FAM102B,ZNF326,BTBD8,LINC01140,CCDC18,BARHL2,MIGA1,NEXN-AS1,GBP7,C1orf146,FAM69A,UOX,FRRS1,GBP1P1,FLJ27354,MIR137HG,MIR137,LHX8,LINC01555,FLJ31662,GEMIN8P4,RBMXL1,LOC646626,SETSIP,ACTG1P4,NBPF6,SNORD45C,MIR553,LOC729930,LOC729970,LOC729987,MIR760,LINC01349,LOC100129046,LOC100129138,LOC100129620,CCDC18-AS1,MIR548N,SPATA1,LINC01364,RTCA-AS1,FPGT-TNNI3K,TMEM56-RWDD3,MIR378G,MIR2682,MIR4423,DPYD-AS1,DPYD-AS2,VAV3-AS1,LOC100996630,LOC100996635,LINC01360,ERICH3-AS1,LOC101927342,LOC101927412,LOC101927434,LINC01361,LOC101927560,LOC101927587,LOC101927844,PKN2-AS1,LINC01057,LOC101928098,LOC101928118,LOC101928241,LOC101928270,LOC101928370,LOC101928436,LOC101928476,MIR7852,MIR7856,MIR7156,LOC102606465,LOC102723661,LINC01461,LINC01307,LOC105378828,LOC105378853 |
| 4p16.3 | 0.021912 | chr4:1190476-1349699 | CTBP1,MAEA,CTBP1-AS2,CTBP1-AS,LOC100130872 |

| Cohort C, Deletions | | | |
|---|---|---|---|
| **cytoband** | **q value** | **wide peak boundaries** | **gene** |
| 7q22.1 | 2.16E-13 | chr7:100894284-100977066 | MUC3A |
| 13q12.11 | 1.79E-07 | chr13:20225604-23911117 | FGF9,MIPEP,SGCG,IFT88,SAP18,SACS,LATS2,CRYL1,IL17D,TNFRSF19,XPO4,MRPL57,EEF1AKMT1,SKA3,MICU2,ZDHHC20,C1QTNF9B,C1QTNF9B-AS1,BASP1P1,MIPEPP3,LINC00540,SACS-AS1,LINC00327,MIR4499,LINC00539,LINC00424,LINC00621,LINC00367,LINC01046 |
| 6q25.1 | 7.49E-06 | chr6:150402135-151409965 | AKAP12,MTHFD1L,PLEKHG1,ZBTB2,LOC102723831 |
| 12q12 | 0.00011263 | chr12:40348910-40918421 | MUC19 |
| 1q41 | 0.00017779 | chr1:219551076-222563950 | hsa-mir-664,EPRS,HLX,MARK1,AURKAPS1,BPNT1,DUSP10,RAB3GAP2,MARC2,SLC30A10,IARS2,MARC1,C1orf115,HHIPL2,C1orf140,MIR194-1,MIR215,SNORA36B,MIR664A,HLX-AS1,LINC01352,LOC101929771 |
| 16p13.13 | 0.00069911 | chr16:11330862-11547854 | RMI2,LOC101927131,LOC105371083 |
| 4q35.2 | 0.0014586 | chr4:183509616-190214555 | SLC25A4,CASP3,F11,ACSL1,FAT1,FRG1,IRF2,KLKB1,MTNR1A,TLR3,SORBS2,FAM149A,PDLIM3, |

| | | | |
|---|---|---|---|
| | | | UFSP2,LRP2BP,STOX2,CFAP97,TRAPPC11,CENPU,SNX25,ZFP42,ENPP6,RWDD4,PRIMPOL,TRIML2,CCDC110,CYP4V2,F11-AS1,LOC339975,TRIML1,ANKRD37,HELT,LINC01060,C4orf47,FRG2,SLED1,FLJ38576,LOC728175,MIR3945HG,DUX4,DBET,MIR3945,LINC01093,LOC100506272,MIR4455,LINC01262,LOC102723766,LVCAT8,LOC105377590,LINC01596 |
| 2q33.1 | 0.0057246 | chr2:201162804-201653877 | ADRB2,CSNK1A1,DIAPH1,DPYSL3,SLC26A2,FGF1,NR3C1,HTR4,PCDH1,PCDHGC3,PDE6A,POU4F3,PPP2R2B,SPINK1,TAF7,PCDHGB4,HDAC3,RNF14,PCDHGA8,PCDHA9,KIAA0141,JAKMIP2,GNPDA1,TCERG1,SPINK5,ABLIM3,ARHGAP26,PCDHGA12,PCDHB5,IL17B,PCDHB1,PCDH12,LARS,RBM27,PCDHB18P,PCDHB17P,PCDHGC5,PCDHGC4,PCDHGB7,PCDHGB6,PCDHGB5,PCDHGB3,PCDHGB2,PCDHGB1,PCDHGA11,PCDHGA10,PCDHGA9,PCDHGA7,PCDHGA6,PCDHGA5,PCDHGA4,PCDHGA3,PCDHGA2,PCDHGA1,PCDHGB8P,PCDHB15,PCDHB14,PCDHB13,PCDHB12,PCDHB11,PCDHB10,PCDHB9,PCDHB8,PCDHB7,PCDHB6,PCDHB4,PCDHB3,PCDHB2,PCDHAC2,PCDHAC1,PCDHA13,PCDHA12,PCDHA11,PCDHA10,PCDHA8,PCDHA7,KCTD16,PCDHB16,HMHB1,ARAP3,PCYOX1L,SH3TC2,NDFIP1,FBXO38,YIPF5,TIGD6,SPRY4,SLC25A2,PCDHB19P,SPINK7,FCHSD1,SCGB3A2,PPARGC1B,AFAP1L1,GRPEL2,GPR151,SPINK13,JAKMIP2-AS1,PRELID2,SH3RF2,PLAC8L1,STK32A,LOC255187,RELL2,C5orf46,ARHGEF37,SPINK6,MIR143,MIR145,SPINK14,MIR378A,GRXCR2,SPINK9,LOC644762,MIR584,CARMN,LOC729080,LOC100505658,SPRY4-IT1,MIR5197,ARHGAP26-AS1,PPP2R2B-IT1,ARHGAP26-IT1,LOC101926905,LOC101926941,LOC101926975,LOC102546294,GRPEL2-AS1 |
| 5q31.3 | 0.0057246 | chr5:140829810-150004264 | CASP8,CASP10,STRADB,TMEM237,TRAK2,ALS2CR12,ALS2CR11 |
| 8q22.3 | 0.0076956 | chr8:101659653-102249922 | RRM2B,NCALD,MIR5680,LOC104054148 |
| 14q24.3 | 0.0089258 | chr14:74949508-77678493 | ACYP1,ESRRB,FOS,GSTZ1,TGFB3,EIF2B2,SPTLC2,BATF,AHSA1,TMED10,C14orf1,VASH1,TTLL5,ANGEL1,MLH3,POMT2,FLVCR2,GPATCH2L,TMEM63C,NGB,VIPAS39,IRF2BPL,ZC2HC1C,CIPC,NEK9,IFT43,NOXRED1,JDP2,LRRC74A,ISM2,SAMD15,LOC283575,ZDHHC22,TMED8,LINC01220,MIR1260A,LOC100506603,MIR7641-2,LOC102724153,LOC102724190,LINC01629 |
| 17p13.2 | 0.0089258 | chr17:3405469-4680404 | ALOX15,ASPA,ATP2A3,CTNS,ITGAE,P2RX1,P2RX5,UBE2G1,TRPV1,OR1E2,OR3A3,MYBBP1A,ZZEF1,SHPK,TAX1BP3,ANKFY1,NCBP3,EMC6,GSG2,CAMKK1,SPATA22,CYB5D2,GGT6,SPNS2,TRPV3,SPNS3,SMTNL2,P2RX5-TAX1BP3,MIR5096,LOC103021295 |
| 5q35.3 | 0.0089258 | chr5:179025521-179113604 | ZNF354C |
| 19p13.3 | 0.013295 | chr19:5892933-6366580 | CAPS,MLLT1,RFX2,RANBP3,ACSBG2,ACER1,NDUFA11,VMAC,LOC100128568 |
| 9q33.3 | 0.0138 | chr9:126505949-126863308 | LMX1B,ZBTB43 |
| 19q13.41 | 0.028888 | chr19:45516447-56403739 | hsa-mir-643,hsa-mir-220c,AP2A1,KLK3,BAX,BCAT2,C5AR1,CA11,CALM3,CD33,SIGLEC6,CD37,CGB3,AP2S1,CRX,DBP,DMPK,DMWD,EMP3,ETFB,FCAR,FCGRT,FLT3LG,FPR1,FPR2,FPR3,FTL,FUT1,FUT2,GIPR,GPR4,GPR32,GRIN2D,ARHGAP35,GYS1,HAS1,FOXA3,HRC,PRMT1,IL11,IRF3,KCNA7,KCNC3,KCNJ14,KIR2DL1,KIR2DL2,KIR2DL3,KIR2DL4,KIR2DS1,KIR2DS3,KIR2DS4,KIR2DS5,KIR3DL1,KIR3DL2,KIR3DS1,KLK1,KLK2,LAIR1,LAIR2,LHB,LIG1,LIM2,MYBPC2,NDUFA3,NKG7,CNOT3,NOVA2,NPAS1,NTF4,NUCB1,POLD1,PPP2R1A,PPP5C,PRKCG,PRRG2,KLK7,KLK6,KLK10,PTGIR,PTPRH,RPL18,RPL28,RPS9,RPS11,RRAS,CLEC11A,SEPW1,SLC1A5,SLC8A2,SNRNP70,SNRPD2,SPIB,SULT2B1,SULT2A1,SYT5,TNNI3,TNNT1,TULP2,NR1H2,ZNF28,ZNF137P,ZNF175,SYMPK,TEAD2,PPFIA3,PLA2G4C,NAPA,SIGLEC5,PGLYRP1,CYTH2,NCR1,NAPSA,KLK4,ZNF432,DHX34,SAE1,LILRB2,RUVBL2,LILRB1,KDELR1,LILRB5,LILRB4,KLK11,LILRA1,LILRB3,LILRA2,KPTN,KLK8,PNKP,U2AF2,ATF5,PPP6R1,CARD8,ZC3H4,FBXO46,RPL13A,SYNGR4,LILRA4,NUP62,HSPBP1,PPP1R15A,EML2,KLK5,PRKD2,ZNF473,KLK13,CCDC9,PRPF31,IRF2BP1,FGF21,SNORD35A,SNORD34,SNORD33,SNORD32A,SIGLEC7,BBC3,DKKL1,SIGLEC9,SIGLEC8,C5AR2,DHDH,UBE2S,SLC6A16,TFPT,STRN4,CCDC106,EPN1,GLTSCR2,GLTSCR1,EHD2,KLK14,KLK12,SHANK1,NOSIP,ZNF580,HSD17B14,GP6,VRK3,ZNF581,PTOV1,PPP1R12C,TRPM4,QPCTL,FAM83E,EPS8L1,RASIP1,TMEM160,PIH1D1,C19orf73,PNMAL1,TMEM143,ZNF444,ZNF331,KLK15,NLRP2,ZNF701,ZNF83,ZNF415,CABP5,SPHK2,MEIS3,SLC17A7,NAT14,RCN3,TTYH1,PNMAL2,PRR12,PLEKHA4,SCAF1,CACNG8,CACNG7,CACNG6,ZNF350,TSKS,ELSPBP1,LIN7B,HIF3A,ZNF649,TSEN34,MBOAT7,FKRP,ZSCAN5A,LENG1,LILRP2,LILRA6,TBC1D17,ISOC2,MYH14,ZNF665,ZNF613,ZNF702P,ZNF614,FUZ,OPA3,RSPH6A,ZNF611,MED25,BCL2L12,GRWD1,CCDC8,ZNF541,SYT3,AKT1S1,ZNF528,BRSK1,SNORD35B,ZNF347,ZNF577,KMT5C,C19orf48,FIZ1,GALP,SIGLEC10,SIGLEC12,ZNF628,KIR3DX1,ZNF616,ZNF766,ZNF468,ZNF160,CTU1,ZNF765,NLRP12,MYADM,ZNF845,CCDC114,LOC93429,ACPT,CGB5,CGB7,LRRC4B,LENG9,CGB8,GNG8,BIRC8,FAM71E1,RDH13,PTH2,SIGLEC11,CGB1,CGB2,LMTK3,LENG8,KIR3DL3,CLDND2,ZNF816,COX6B2,OSCAR,ZNF813,JOSD2,IZUMO2,CPT1C,ALDH16A1,NTN5,NLRP13,NLRP8,NLRP5,ZNF787,VSIG10L,C19orf84,SPACA6,ZNF480,ZNF534,ZNF578,ERVV-1,TMEM190,TMC4,TPM3P9,ZNF524,ZNF784,CCDC155,DACT3,SIX5,IGFL2,NLRP4,ZNF542P,ZNF582,CDC42EP5,ZNF836,ZNF610,ZNF600,ZNF320,ZNF579,ZNF114,ZNF525,SPACA4,NLRP7,ADM5,NLRP11,TMEM86B,INAFM1,NAPSB,IL4I1,SSC5D,TPRX1,MAMSTR,IZUMO1,EMC10,MGC45922,KLK9,SIGLEC17P,SIGLECL1,ZNF615,ZNF841,LOC284379,VSTM1,TMEM150B,FAM71E2,VN1R2,VN1R4,NLRP9,MYPOP,NANOS2,C19orf81,ZNF677,RFPL4A,ZSCAN5B,DNAAF3,LILRA5,IGFL1,C19orf68,ZNF582-AS1,BHMG1,IGFL3,ZNF808,ZNF888,ZNF761,TMEM238,ZNF818P,ZNF321P,LOC400706,SIGLEC16,LOC400710,ZNF880,IGLON5,MIRLET7E,MIR125A,MIR150,MIR99B,TARM1,MIR330,MIR371A,MIR372,MIR373,IGFL4,DPRX,ASPDH,MIR512-1,MIR512-2,MIR498,MIR520E,MIR515-1,MIR519E,MIR520F,MIR515-2,MIR519C,MIR520A,MIR526B,MIR519B,MIR525,MIR523,MIR518F,MIR520B,MIR518B,MIR526A1,MIR520C,MIR518C,MIR524,MIR517A,MIR519D,MIR521-2,MIR520D,MIR517B,MIR520G,MIR516B2,MIR526A2,MIR518E,MIR518A1,MIR518D,MIR516B1,MIR518A2,MIR517C,MIR520H,MIR521-1,MIR522,MIR519A1,MIR527,MIR516A1,MIR516A2,MIR519A2,KLKP1,LOC645553,FAM90A27P,SBK2,SEC1P,SNORD23,SNORD88A,SNORD88B,SNORD88C,MIR642A,MIR643,CCDC61,CEACAM18,SHISA7,RFPL4AL1,LINC01530,MIR769,SIGLEC14,MIR935,SNAR-G1,SNAR-F,SNAR-A1,SNAR-A12,LOC100129083,SBK3,BSPH1,KIR2DS2,SNAR-A3,SNAR-A5,SNAR-A7,SNAR-A11,SNAR-A4,SNAR-A6,SNAR-A8,SNAR-A10,SNAR-C2,SNAR-C4,SNAR-E,SNAR-B1,SNAR-C1,SNAR-C3,SNAR-D,SNAR-G2,SNAR-A14,ERVV-2,EML2-AS1,MIR1283-2,MIR1323,MIR1283-1,MIR3191,MIR3190,MIR320E,MIR4324,MIR642B,NAPA-AS1,CARD8-AS1,PPP5D1,PTOV1-AS1,DACT3-AS1,GFY,ZNF865,ZNF816-ZNF321P,MIR4752,MIR371B,MIR4749,MIR4750,MIR4751,MIR5088,NUCB1-AS1,LOC101059948,ZNF350-AS1,LOC101928295,PTOV1-AS2,LOC101928517,ZNF649-AS1,LOC101928804,LOC101928886,SPACA6P-AS,MIR6799,MIR6800,MIR6802,MIR6804,MIR6805,MIR8074,MIR8061,MIR6798,MIR6803,MIR7975,MIR6801,ZNF528-AS1,LENG8-AS1,LOC105372440,LOC105372441,LOC105372444,LOC105447645,PLA2G4C-AS1,GLTSCR2-AS1 |
| 10q23.1 | 0.036878 | chr10:50884518-112289801 | hsa-mir-3158-1,ACTA2,ADD3,ADK,ADRA2A,ANK3,ANXA2P3,ANXA7,ANXA11,FAS,ARL3,BMPR1A,CAMK2G,ENTPD1,CDK1,CHUK,ABCC2,COL13A1,COL17A1,COX15,CPN1,CYP2C19,CYP2C8,CYP2C9,CYP2C18,CYP17A1,CYP26A1,DNA2,DNTT,DUSP5,EGR2,EIF4EBP2,FGF8,GLUD1,GLUD1P3,GOT1,GRID1,HELLS,HHEX,HK1,HNRNPH3,TLX1,HPS1,HTR7,IDE,IFIT2,IFIT1,IFIT3,KCNMA1,KIF11,LIPA,MAT1A,MBL2,MXI1,NDUFB8,NFKB2,NODAL,P4HA1,PAX2,PCBD1,PDE6C,PGAM1,PITX3,PLAU,PPA1,PPP1R3C,PPP3CB,PRF1,SRGN,PRKG1,PSAP,PSD,PTEN,ALDH18A1,RBP4,RGR,RPS24,SCD,SFR |

| | | | |
|---|---|---|---|
| | | | P5,SFTPD,FBXW4,SLIT1,SNCG,SUPV3L1,TACR2,TAF5,TFAM,TLL2,UBE2D1,VCL,VDAC2,WNT8B,XPNPEP1,CCDC6,SLC25A16,SHOC2,NDST2,MBL1P,LIPF,GBF1,LDB1,SGPL1,BTRC,CH25H,PKD2L1,BTAF1,PAPSS2,INA,PDLIM1,SMC3,NEURL1,DDX21,LGI1,NOLC1,DLG5,GSTO1,CHST3,VPS26A,MINPP1,KIF20B,SEC24C,SH3PXD2A,SLK,SPOCK2,ZNF518A,RHOBTB1,FRAT1,PPIF,ACTR1A,SMNDC1,NPM3,MICU1,RPP30,SORBS1,ERLIN1,LBX1,NRG3,MGEA5,ADIRF,POLR3A,ZWINT,LDB3,ECD,CPEB3,ZNF365,DKK1,NT5C2,PDCD11,SORCS3,ZSWIM8,WAPL,PPRC1,RRP12,TBC1D12,DNAJC9,DNMBP,CSTF2T,FRAT2,SIRT1,KAT6B,TSPAN15,IFIT5,DPCD,SEC31B,NUDT13,HERC4,LRIT1,TCTN3,KIF1BP,C10orf12,ANKRD2,CNNM1,MYOF,AP3M1,ANKRD1,GHITM,PALD1,PDCD4,R3HCC1L,POLL,CTNNA3,BLNK,NRBF2,KCNIP2,NEUROG3,ASCC1,EXOSC1,MRPS16,CALHM2,CUTC,PLCE1,DUSP13,SUFU,PANK1,CCSER2,EXOC6,FAM35A,DDIT4,CCNJ,MARCH5,DNAJB12,CNNM2,WBP1L,CRTAC1,CEP55,LRRC20,CWF19L1,SLC29A3,RNLS,PI4K2A,H2AFY2,HIF1AN,RUFY2,SLF2,CCAR1,CISD1,DNAJC12,C10orf2,SAR1A,TM9SF3,ENTPD7,ZMIZ1,AS3MT,STAMBPL1,GPAM,SEMA4G,MYOZ1,AVPI1,HPSE2,CDH23,PBLD,NPFFR1,C10orf54,MMS19,NOC3L,PCDH15,CUEDC2,DDX50,FBXL15,C10orf76,HPS6,MMRN2,MFSD13A,SYNPO2L,C10orf95,PDZD7,OBFC1,UBTD1,BICC1,TMEM254,HKDC1,CFAP43,TET1,TNKS2,TRIM8,TSPAN14,KAZALD1,SFXN3,SLC25A28,ELOVL3,MARVELD1,C10orf11,PCGF6,ARID5B,LOXL4,ZDHHC16,FAM213A,DYDC2,PCGF5,LZTS2,PHYHIPL,LCOR,MRPL43,PLA2G12B,MYPN,PYROXD2,USMG5,ZNF503,AIFM2,ADO,ATAD1,ARHGAP19,JMJD1C-AS1,ITPRIP,OLMALINC,MCU,CDHR1,BBIP1,OPALIN,OPN4,HOGA1,SORCS1,CHCHD1,MSS51,CFAP70,PIK3AP1,MORN4,ZFYVE27,COMTD1,FRA10AC1,ANKRD22,SFXN2,BORCS7,NUDT9P1,AGAP11,GSTO2,SFR1,CALHM3,ANAPC16,ADAMTS14,ACSM6,SAMD8,LIPJ,CFL1P1,DYDC1,EIF5AL1,HECTD2,FGFBP3,USP54,NKX2-3,SLC35G1,CFAP58,FUT11,OIT3,LINC00858,TMEM254-AS1,PLAC9,C10orf107,TMEM26,ZCCHC24,UNC5B,STOX1,C10orf35,TYSND1,RTKN2,TBATA,ATOH7,SLC16A9,FAM13C,REEP3,JMJD1C,ZNF503-AS1,IPMK,CALHM1,PIPSL,BLOC1S2,RBM20,PDCD4-AS1,FLJ37201,LINC01553,LOC283045,ZMIZ1-AS1,FAM149B1,FFAR4,DUPD1,LIPM,CYP26C1,LRIT2,LRRTM3,SH2D4B,C10orf99,SLC16A12,CC2D2B,LBX1-AS1,GOLGA7B,MIR107,C10orf105,C10orf62,C10orf55,LINC00595,DNAJC9-AS1,LINC00857,LINC00863,IFIT1B,MIR346,MIR146B,LOC642361,HOST2,FAM25A,LIPK,LIPN,LINC00865,RPL13AP6,BEND3P3,SFTPA1,SNORA12,SNORD98,MIR605,MIR606,MIR607,MIR608,MIR609,NUTM2A,NUTM2D,NUTM2A-AS1,LINC00864,ENTPD1-AS1,FAM133CP,RPL13AP5,UNC5B-AS1,RPEL1,AGAP5,BMS1P4,SFTPA2,NUTM2B,LOC729815,POU5F1P5,TLX1NB,MIR936,C10orf131,PLCE1-AS1,DLG5-AS1,LOC100130698,ZNF503-AS2,ACTA2-AS1,LINC00856,KLLN,HECTD2-AS1,DNMBP-AS1,BMS1P21,KCNIP2-AS1,MIR1287,MIR1296,MIR1307,MIR548F1,MIR1254-1,FAS-AS1,MIR548E,MIR3157,MIR3158-1,MIR3158-2,MTRNR2L5,MIR3924,SLIT1-AS1,LINC00866,RPARP-AS1,SH3PXD2A-AS1,CFAP58-AS1,SORCS3-AS1,ADD3-AS1,PRKG1-AS1,LINC00844,MRLN,LINC00845,ZSWIM8-AS1,GRID1-AS1,TNKS2-AS1,MARK2P9,BORCS7-ASMT,ARHGAP19-SLIT1,MIR4680,MIR4679-1,MIR4679-2,MIR4676,MIR4678,MIR4482,MIR4685,LINC00502,NUTM2B-AS1,SLC16A12-AS1,LINC01375,LOC101926942,PLCE1-AS2,LOC101927278,LINC01475,LINC01514,LOC101927419,LOC101927472,LOC101927523,LOC101927549,LINC01468,TMEM26-AS1,LOC101928887,LINC01515,LOC101928961,LOC101928994,PPP3CB-AS1,LOC101929165,LOC101929234,KCNMA1-AS3,KCNMA1-AS2,KCNMA1-AS1,LOC101929574,NRG3-AS1,LINC01519,LOC101929646,LOC101929662,LINC01520,MIR6507,MIR7152,MIR7151,LOC102723377,LOC102723439,LOC102723665,LOC102723703,NEURL1-AS1,LOC102724719,LINC01435,LOC105378311,LOC105378330,LOC105378349,LOC105378367,LOC105378385,LOC105378397,XLOC_008559,LOC105378430,LOC105378470,CCEPR |
| 2q37.3 | 0.036878 | chr2:240896041-240998999 | LOC200772 |
