## Supplementary figures and images for "The somatic and germline mutational landscape of HPV-negative oral cancer patients with a history of chewing tobacco and betel nut use"

### Supplementary figure 1

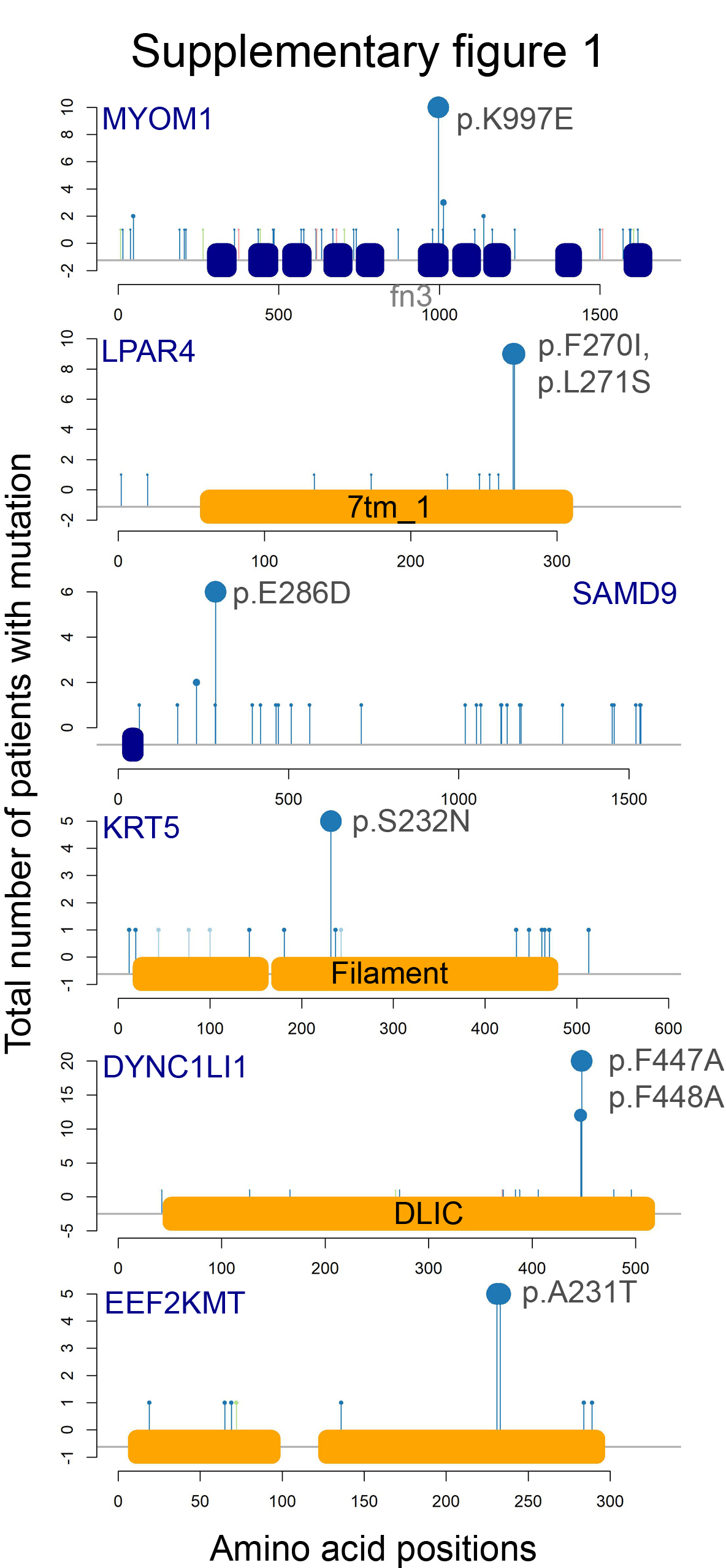

### Supplementary figure 2

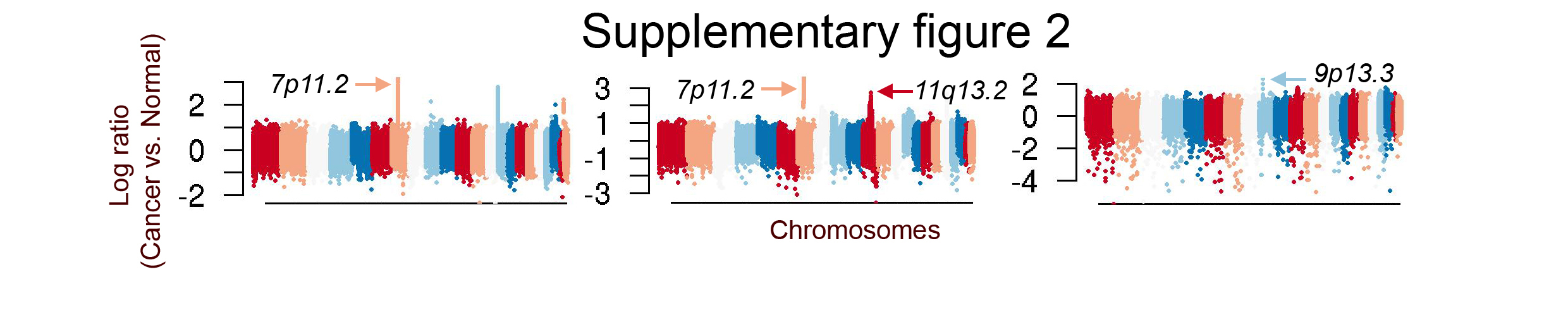

### Supplementary figure 3

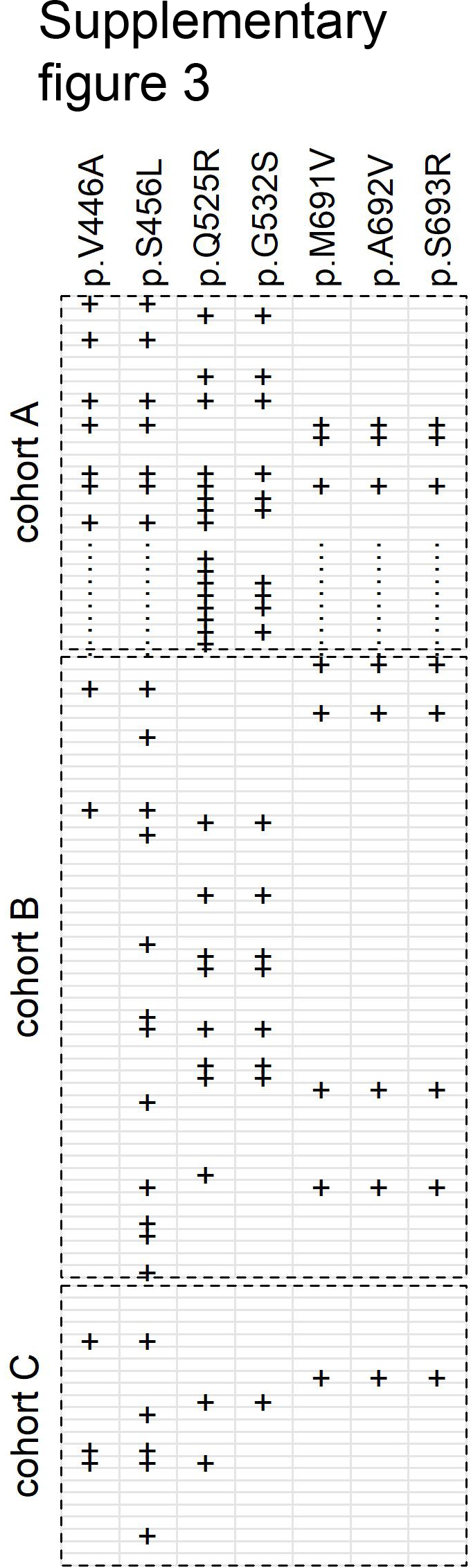
